# Stereoelectroencephalography accuracy in a series of over 3000 trajectories

**DOI:** 10.64898/2026.07.14.26358071

**Authors:** Arun Thurairajah, Greydon Gilmore, Amit RL Persad, Amir Saam Youshani, Alaa Taha, Mohamad Abbass, Brendan Santyr, Khalid Al Orabi, Giovanni Pellegrino, Jorge G. Burneo, Giovanni Pellegrino, Ana Suller-Marti, Western Epilepsy Research Group, Andrew G. Parrent, Keith MacDougall, David A. Steven, Jonathan C. Lau

## Abstract

**Background and Objectives:** Stereoelectroencephalography (SEEG) involves the implantation of intracerebral electrodes to investigate drug-resistant epilepsy. SEEG requires millimetric accuracy to ensure safety and optimal mapping. Although studies have evaluated SEEG accuracy, there is substantial variability in reporting. Here we report on implantation accuracy in a large series using the most common accuracy metrics described in the literature and perform a detailed analysis of contributing factors.

**Methods:** SEEG implantations between 2013 and 2025 were included. Application accuracy was computed for each implanted electrode. Specifically, Euclidean, radial, depth, and angle error were calculated at both target and entry points. Correlative and multivariable analyses were conducted between each variable and error metric. Trajectories were also grouped by atlas-derived lobar target.

**Results:** No metrics met assumptions of normality and thus we report accuracy using median with interquartile range (IQR). In a series of 3176 trajectories, median Euclidean target and entry errors were lower for robot-assisted electrodes (n=2858) at 2.19 (IQR: 1.54-2.98) mm and 1.38 (IQR: 0.89-2.01) mm respectively, compared to frame-based (n=318, p<.001) at 2.76 (IQR:1.79-3.76) mm and 2.21 (IQR: 1.42-3.32) mm. Correlation and multivariable regression analysis showed target error was positively correlated with implantation angle, scalp thickness, skull thickness, and trajectory length. Target error was also higher in obese patients. On lobar analysis, parietal lobe trajectories were the most accurate and frontal lobe trajectories were the least accurate. On temporal lobe trajectory analysis, posterior hippocampus trajectories were the most accurate and temporal pole trajectories were the least accurate. Presence of mesial temporal sclerosis also impacted accuracy.

**Conclusions:** We present a detailed description of SEEG implantation accuracy, demonstrating the superior accuracy and speed of robot-assisted to frame-based methods. Furthermore, we analyzed how accuracy varies with specific factors from a global to trajectory level, which can be accounted for when planning SEEG implantations.

## INTRODUCTION

Introduced by Talairach and Bancaud in 1965, stereoelectroencephalography (SEEG) has become the dominant form of intracranial monitoring for the identification of the epileptogenic zone (EZ) in patients with drug-resistant epilepsy.^1–5^ Technologic advancement has driven a shift from traditional stereotactic frame-based methods to robot-assisted implantation, greatly improving procedural efficiency^6,7^ and overall implantation accuracy.^6–11^

Trajectory errors can impact the Phase II investigation if the seizure onset zone (SOZ) is missed,^12^ or lead to complications such as hemorrhage. Accuracy is also critical for SEEG-guided radiofrequency thermocoagulation (RFTC).^13–17^ Trajectory error is quantified between the planned and actual trajectory with the most reported metrics being the Euclidean error (or vector error), radial error (or lateral error^12^), depth error, and angle error. Prior work has shown that trajectory error is positively correlated with skull thickness, trajectory length, and implantation angle.^6,10,18–23^

There is substantial variability in how implantation accuracy is reported, specifically regarding the choice of evaluation metrics and contributing factors. When this data is aggregated, only a few common metrics and factors can be pooled. In addition, by performing more granular trajectory-level analysis the extent of inaccuracy for specific trajectories and scenarios can be better characterized. To understand these relationships, we investigated how various global and trajectory-level factors impact SEEG accuracy for a series of over 3000 trajectories.

## METHODS

We conducted a retrospective review of patients who underwent SEEG implantation between February 2013 and July 2025. Procedures conducted from 2013 to 2017 employed a traditional frame-based approach, with robotic assistance routinely used from April 2017. Baseline variables included patient demographics, prior cranial surgeries, and radiological findings. Procedural data included total case duration, anaesthesia time (intubation-to-extubation), operative time (skin-to-skin), and perioperative complications.

### SEEG: Surgical Planning and Implantation

Electrode trajectories were planned based on clinically indicated hypotheses about the seizure onset and propagation zones in multidisciplinary epilepsy surgery rounds. A gadolinium-enhanced volumetric T1-weighted (T1w) magnetic resonance imaging (MRI) scan served as the reference for SEEG planning and was acquired on either: a 1.5 T Signa General Electric (Milwaukee, Wisconsin, USA), a 1.5 T Siemens MAGNETOM Sola (Erlangen, Germany), or a 3T Siemens MAGNETOM Vida (parameters in Supplementary Materials). A T1w-MRI without gadolinium was also acquired for cortical surface reconstruction and template registration.^24^

SEEG implantation followed a previously described surgical method.^4,25^ A Leksell G-frame (Elekta AB, Stockholm, Sweden) was fixed to the patient’s head after a bilateral scalp block, followed by a pre-operative contrast-enhanced CT scan (parameters in Supplementary Materials). Frame-based implantations were planned with the Framelink software (v5.4.1; Medtronic, Dublin, Ireland) and electrodes were inserted using a standard arc-frame technique. Planned coordinates were extracted from paper medical records where available. Robot-assisted cases were planned with Neuroinspire™ and electrodes were implanted using the Neuromate® stereotactic robot (Renishaw plc, UK); the Leksell frame was used solely for head fixation and MRI-CT fusion.

All electrodes were implanted under fluoroscopic guidance and secured with bolt anchors. We commonly implanted 10-contact depth electrodes (AdTech Medical, Racine, WI; 3–6 mm center-to-center contact spacing, 0.86 mm diameter). 38 Dixi Medical (Besançon, France) and 3 Behnke-Fried electrodes were also implanted. Bilateral 4-contact reference electrodes (AdTech) were placed into the frontoparietal galea. The distance between scalp and center of dura was measured for each trajectory to determine the anchor bolt length required. A postoperative non-contrast CT scan was acquired for electrode localization.

### Electrode Localization and Accuracy Metrics

We developed an image processing pipeline building from SEEG Assistant.^26^ The postoperative CT was rigidly registered (6 degrees of freedom or DOF) to the reference contrast-T1w using the Advanced Normalization Tools (ANTs; v2.2.0) library.^27^ Entry and target points were then defined using the Markups Module with 3D Slicer (v5.8.1).^28^ The entry point was defined at the dura on the pre-operative contrast T1w scan, while the target was defined at the tip of the electrode on the post-operative CT. Electrode implantation accuracy was retrospectively computed between entry and target points for the planned trajectory and the implanted electrode (Figure 1). Error metrics and variables are summarized in Table 1.

**Figure 1.**
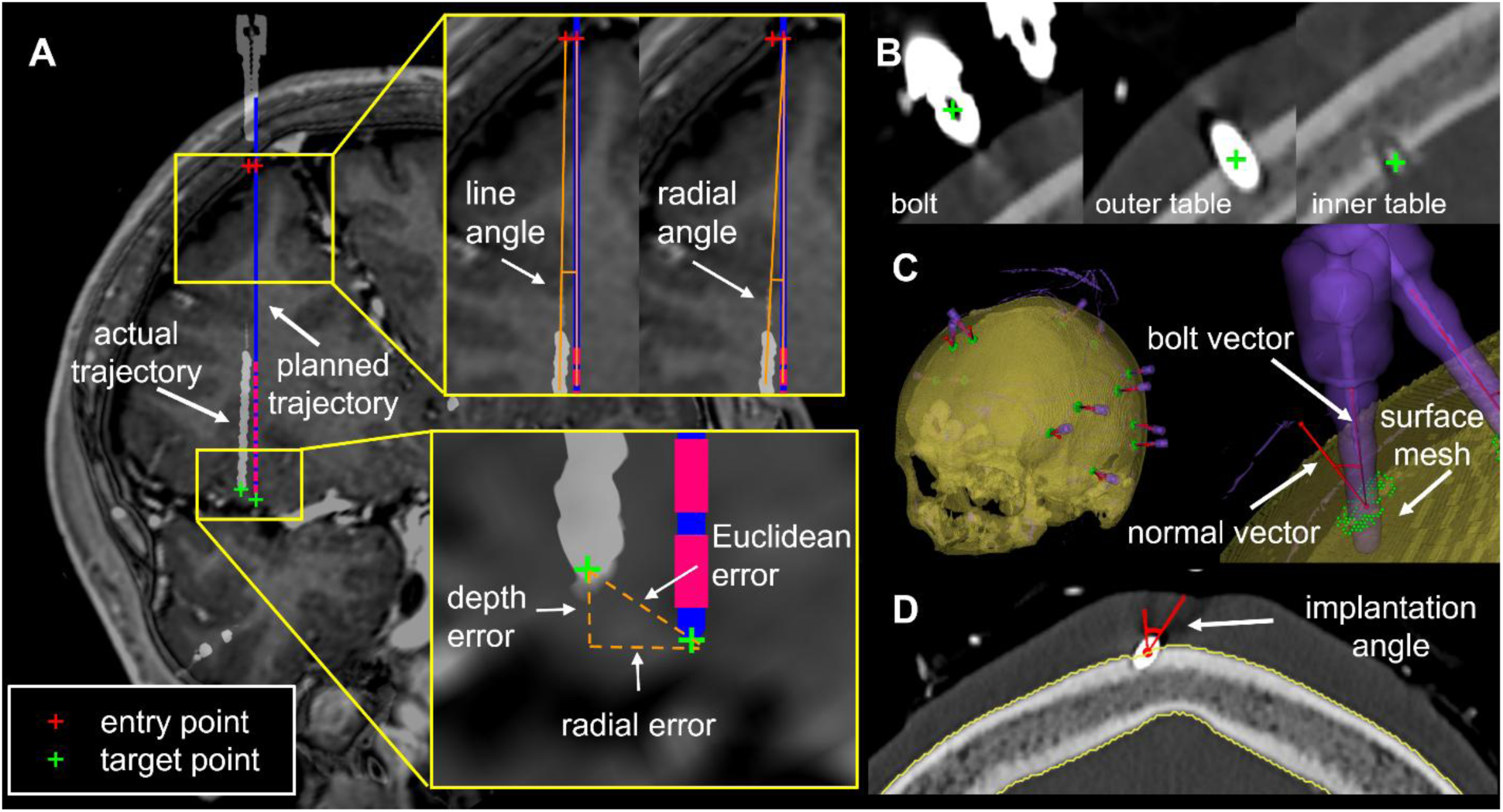
Schematic illustrating fiducial points, error metrics, and skull mesh formation to compute entry angle. (A) Error metrics associated with planned and actual trajectories. (B) three points at the center of the bolt head, outer table and inner table to form the bolt vector. (C) overview of sampled mesh surface point rafts around the bolt impact point (green points). (D) cross-section of anchor bolt point raft used to compute bolt vector, normal vector, and the implantation angle.

**Table 1.** SEEG accuracy metrics and clinical variables.

| Metric | Description |
| --- | --- |
| *error metrics* |  |
| **Euclidean error (mm)** | measured in 3-dimensions (x, y, z) computed as the straight-line distance between planned and actual coordinates |
| **radial error (mm)** | measured in 2-dimensions, represents the distance between the planned point and the actual point orthogonal to the axis of the planned trajectory |
| **depth error (mm)** | computed as the difference in depth between the planned and actual points |
| *angular error* |  |
| **line angle (°)** | defined as the angle between the planned and actual trajectory, independent of starting point, also referred to as the angular error<sup>38,39</sup> |
| **radial angle (°)** | the angle between the planned trajectory and the vector from the planned entry point to the actual target point |
| *clinical variables* |  |
| **scalp-dura distance (mm)** | distance between scalp and center of dura along the planned trajectory path |
| **skull thickness (mm)** | distance between the inner and outer table, determined manually by points placed on the inner and outer table along anchor bolt path |
| **scalp thickness (mm)** | difference between scalp-dura distance and skull thickness |
| **trajectory length (mm)** | distance between planned target and entry points |
| **implantation angle (°)** | inspired by the approach from Janca et al.<sup>21</sup>, defined as the angle between the anchor bolt vector and the *normal vector* (a vector orthogonal to the skull at the bolt impact point). Similar to prior studies that have computed entry/trajectory angle<sup>17,18,20</sup> and approach angle<sup>40</sup> |
| **procedure duration (min)** | time from first skin incision to closure |
| **anesthesia duration (min)** | time from start of anesthesia start to end of anesthesia service |
| **time between MRI and implantation (days)** | the number of days prior to surgery that the preoperative T1w MRI was acquired |
| **body mass index (BMI; kg/m<sup>2</sup>)** | taken on day of surgery prior to admission |
| **cerebrospinal fluid (CSF) volume (mm<sup>3</sup>)** | *atropos3seg* 3-tissue segmentation values from the pre-operative T1w MRI |

To perform atlas fitting, the non-contrast T1w (in the reference T1w space) was non-linearly registered to the *tpl-MNI152NLin2009cSym* T1w template^29,30^ using the symmetric diffeomorphic normalization (*SyN*) deformation model in ANTS.^31–35^ An affine registration^36^ (12 DOF) was performed with the *greedy* toolbox^*^ (parameters in Supplementary Materials) to initialize the non-linear deformation. Registration was done between the brain-masked template and a T1w masked from the ANTs *atropos3seg* 3-tissue segmentation.^37^

### Implantation Angle Calculations

The implantation angle was semi-automatically defined for each trajectory. With 3D Slicer, raters placed three additional points along each trajectory at the center of the bolt head, on the outer table of the skull, and where the anchor bolt laid at the inner table, defining a vector for the bolt anchor (Figure 1B). The pre-operative CT was used to create a skull mesh and Principal Component Analysis was used to define an “impact point normal vector” orthogonal to the skull. The implantation angle was calculated between the bolt and normal vector and manually reviewed. Inter-rater reliability was assessed for 21 patients (Supplementary Figure 1). Previous work by Rollo et al.^18^ compared the accuracy of orthogonal vs. oblique (>30°) trajectories, stratifying angles into four bins at 15-degree increments (<30°, 30-45°, 45-60°, >60°). Due to a lack of extremely oblique trajectories (>60°) in our cohort, we stratified angles into <15°, 15-30°, 30-45°, >45°.

### Lobar and Trajectory-Based Analysis

Trajectories were labelled into six lobes using the *USCLobes* atlas^38^, nonlinearly registered to the *MNI152NLin2009cSym* template space.^29,30^ The non-linear registration from the *USCLobes* template to MNI space was computed using the same methodology as atlas-fitting. Trajectories were also labeled by anatomical target at the time of planning, referred to as the surgeon-labelled trajectory (see Supplementary Materials for naming schema).

### Statistical Analysis

Shapiro-Wilk test was used to evaluate normality of implantation variables and accuracy metrics. Mann-Whitney U test and Spearman’s rank correlation with Bonferroni correction was performed for non-parametric continuous variables. A multivariable linear regression model was fit to predict target radial error from pre-implantation variables. Multicollinearity was assessed using the variance inflation factor (VIF); variables with a VIF>10 were removed.^39^ Finally, logistic regression analysis was used to identify the probability of the target radial error exceeding 2 mm based on implantation angle. This was repeated to determine whether angle cutpoints differ between lobes or surgeon-labels by comparing covariate and interaction effect models.^39^ All statistical testing was done in Python (V3.13.0) with SciPy (v1.2.0), statsmodels (v0.14.6), and scikit-learn (v1.7.2).

## RESULTS

Over a 12-year period, 340 patients underwent SEEG implantation for a total of 3856 electrodes. Implantation accuracy could be calculated for 260 patients and 3176 electrodes (Table 2). All accuracy metrics and variables are in Table S1. Of note, no recorded variables met assumptions of normality (p<0.01), further confirmed on visual inspection of data (Supplementary Figure 2). Thus, we report our accuracy results using median with IQR and use nonparametric statistics. The mean and standard deviation can be found in the Supplementary Materials to facilitate comparisons with previous studies.

**Table 2.** Study Demographics.

|  | Robot-assisted<br>(2858 electrodes) |  | Frame-based<br>(309 electrodes) |  | *p*-value |
| --- | --- | --- | --- | --- | --- |
| Number of Patients, n (%) | 229 |  | 31 |  |  |
| Female | 112 (48.91%) |  | 16 (51.61%) |  | .779 |
| Male | 117 (51.09%) |  | 15 (48.39%) |  |  |
| Mean age (years) | 34.60 ± 12.70 |  | 33.48 ± 10.73 |  | .971 |
| Mean weight (kg) | 79.58 ± 21.15 |  | 79.37 ± 22.64 |  | .545 |
| Mean no. of electrodes | 12.33 ± 3.07 |  | 10.23 ± 2.31 |  | .003* |
| Implantation side, n (%) |  |  |  |  |  |
| Bilateral | 183 (79.91%) |  | 15 (48.39%) |  |  |
| Unilateral left | 28 (12.23%) |  | 11 (35.48%) |  |  |
| Unilateral right | 18 (7.86%) |  | 5 (16.13%) |  |  |
| Anaesthesia Duration (mins) | 167.18 ± 44.90 |  | 220.71 ± 42.60 |  | < .001* |
| Procedure Duration (mins) | 90.58 ± 30.76 |  | 140.42 ± 31.09 |  | < .001* |
| Imaging to Implantation<br>(days) | 121.61 ± 162.43 |  | 57.83 ± 160.05 |  | .005* |
| Monitoring in EMU (days) | 14.92 ± 7.88 |  | 11.97 ± 6.00 |  | .346 |
| Euclidean error (mm) | *median (IQR)* | *mean ± sd* | *median (IQR)* | *mean ± sd* |  |
| Entry | 1.38 (0.89-2.01) | 1.69 ± 1.73 | 2.21 (1.42-3.32) | 2.67 ± 2.23 | < .001* |
| Target | 2.19 (1.54-2.98) | 2.46 ± 2.02 | 2.76 (1.79-3.76) | 3.16 ± 2.17 | < .001* |
| Radial error (mm) |  |  |  |  |  |
| Entry | 1.04 (0.64-1.54) | 1.26 ± 1.53 | 1.87 (1.19-2.68) | 2.16 ± 1.68 | < .001* |
| Target | 1.35 (0.85-2.02) | 1.63 ± 1.59 | 2.17 (1.23-3.14) | 2.48 ± 2.03 | < .001* |
| Depth error (mm) |  |  |  |  |  |
| Entry | 0.62 (0.24-1.23) | 0.91 ± 1.04 | 0.88 (0.34-1.59) | 1.25 ± 1.73 | < .001* |
| Target | 1.32 (0.66-2.11) | 1.55 ± 1.60 | 1.16 (0.55-1.93) | 1.49 ± 1.48 | .025 |

Robot-assisted SEEG implantations accounted for 2858 consecutive electrodes in 229 patients (errors visually summarized in Figure 2). For the frame-based group, planned coordinates were retrieved for 318 electrodes in 31 patients. On average, more electrodes per patient were implanted in the robot-assisted group compared to the frame-based group (p=.003). Despite this both anesthetic time (p<.001) and surgical time (p<.001) were shorter in the robot-assisted group. Robot-assisted electrodes were also more accurate (p<.001) than frame-based at entry (Euclidean, radial, and depth) and target points (Euclidean and radial). For robot-assisted electrodes, target Euclidean error was 2.19 (IQR: 1.54-2.98) mm and entry Euclidean error was 1.38 (0.89-2.01) mm. For frame-based electrodes, target Euclidean error was 2.76 (1.79-3.76) mm and entry Euclidean error 2.21 (1.42-3.32) mm.

**Figure 2.**
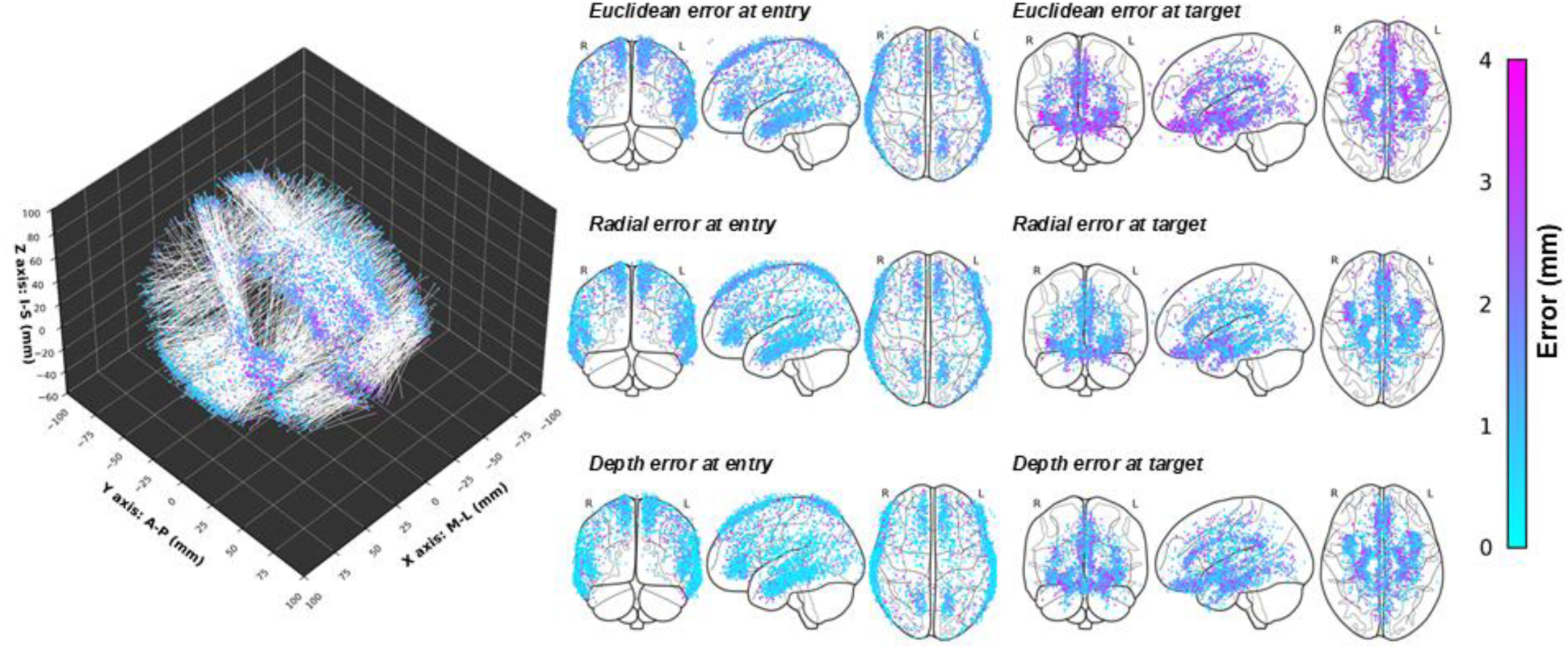
Robot-assisted SEEG implantation accuracy at target and entry points (n = 2858 electrodes, 229 patients). Left panel shows all planned trajectories and Euclidean errors in *MNI152NLin2009cSym* space. Right highlights the Euclidean, radial and depth errors.

In total there were 23 intracranial hemorrhages; only two (0.6%) were symptomatic (headaches/diplopia for 1 patient, GCS 14/headaches for the other) and no patients returned to the OR. Prior to introduction of the robot one patient suffered a fatal intracranial hemorrhage, reported in a previous study^4^; the case was aborted and accuracy was not recorded.

Eight electrodes with target Euclidean error greater than 10 mm were tagged as potential outliers. On review, the high degree of error was due to one of the following factors: electrode pull back (n=2), bending at the electrode tip (n=1), or mislabeling of the planned target coordinates (n=5). Thus, they were removed from subsequent accuracy analysis.

### Correlation analysis of accuracy metrics and variables

Spearman’s rank correlation for the robot-assisted cohort is summarized in Figure 3. Target radial error was most correlated to implantation angle (r=0.28, p<.001), trajectory length (r=0.18, p<.001), scalp thickness (r=0.16, p<.001), and skull thickness (r=0.11, p<.001). Entry radial error was most correlated to implantation angle (r=0.20, p<.001) and scalp thickness (r=0.12, p<.001). BMI was also correlated with target radial error (r=0.10, p<.001), and when stratified, obese patients (BMI≥30, n=67) had a higher target Euclidean (2.46 mm, IQR: 1.94-2.87 vs 2.28 mm, IQR: 1.91-2.55, p<.05) and radial error (1.67 mm, IQR: 1.31-2.00, vs 1.43 mm, IQR: 1.20-1.74, p<.05) than non-obese patients (n=161).

**Figure 3.**
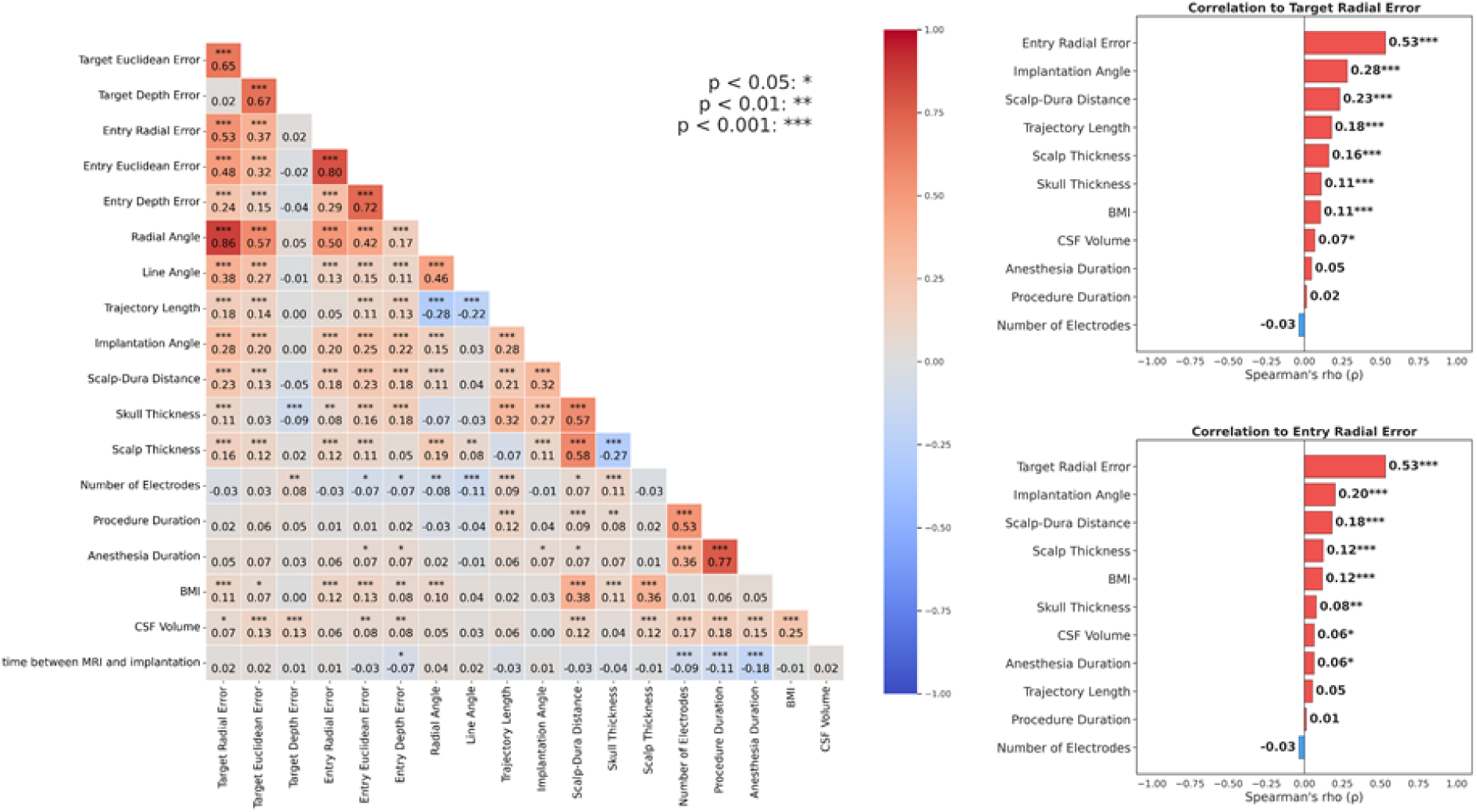
Spearman correlation matrix between SEEG accuracy metrics and operative variables in robot-assisted cohort (n = 2858), corrected for multiple comparisons via Bonferroni method. Tornado plots highlight the strongest correlations to target radial error and entry radial error.

### Multivariable linear analysis for target radial error

After removing variables with VIF>10 and scalp-dura distance (given that scalp thickness was derived from scalp-dura), a multivariable linear model produced an adjusted R^2^ of 0.150 (F-statistic=49.16, see Table 3). Skull thickness (β=0.0283, p<.0005), scalp thickness (β=0.0487, p<.0005), implantation angle (β=0.0274, p<.0005), trajectory length (β=0.0067, p<.0005), the number of electrodes (β=-0.0261, p<.0005) and BMI (β=0.0061, p<.05) reached significance. Individual regression plots are in Supplementary Figure 3.

**Table 3.** Multivariable linear regression of trajectory-related variables to radial error. (r^2^ = 0.150, F-statistic of 49.61).

|  | **$\beta$ Coefficient (95% CI)** | **t** | **P>\|t\|** |
| --- | --- | --- | --- |
| **Intercept** | -0.0155 (-0.284, 0.253) | -0.804 | 0.422 |
| **Trajectory Length** | *0.0067 (0.005, 0.009)* | *5.933* | *p<0.0005* |
| **Implantation Angle** | *0.0274 (0.023, 0.031)* | *13.255* | *p<0.0005* |
| **Scalp Thickness** | *0.0487 (0.035, 0.062)* | *6.940* | *p<0.0005* |
| **Skull Thickness** | *0.0283 (0.014, 0.043)* | *3.814* | *p<0.0005* |
| **Number of Electrodes** | *-0.0261 (-0.039, -0.013)* | *-3.835* | *p<0.0005* |
| **Procedure Duration** | 0.0008 (-0.001, 0.003) | *0.801* | 0.423 |
| **Anesthesia Duration** | 0.0004 (-0.001, 0.002) | *0.683* | 0.495 |
| **BMI** | *0.0061 (0.000, 0.012)* | *2.045* | *p<0.05* |
| **CSF Volume** | $4.31 \times 10^{-7}$ (-1.66$\times 10^{-7}$, 1.03$\times 10^{-6}$) | 1.415 | 0.157 |
| **time between MRI and implantation** | $1.35 \times 10^{-4}$ (-1.08$\times 10^{-4}$, 0.000378) | 1.092 | 0.275 |

### Electrode accuracy in relation to implantation angle

Trajectory errors across four angle categories are summarized in Table 4. The median implantation angle was 20.30° (13.47°-26.73°); oblique trajectories (>30°) were a small proportion (n=446, 18.6%) of our cohort. Kruskal-Wallis test demonstrated that Euclidean and radial errors were statistically different between angle categories for target and entry points (p<.00001). Mann-Whitney U testing showed Euclidean and radial errors to be statistically different (p<.005) for all categories except the 15-30° group; only target radial error reached significance (p=.011). Logistic regression analysis identified a cutpoint of 22.25° in predicting the probability of an electrode surpassing a target radial error of 2 mm (Figure 4).

**Figure 4.**
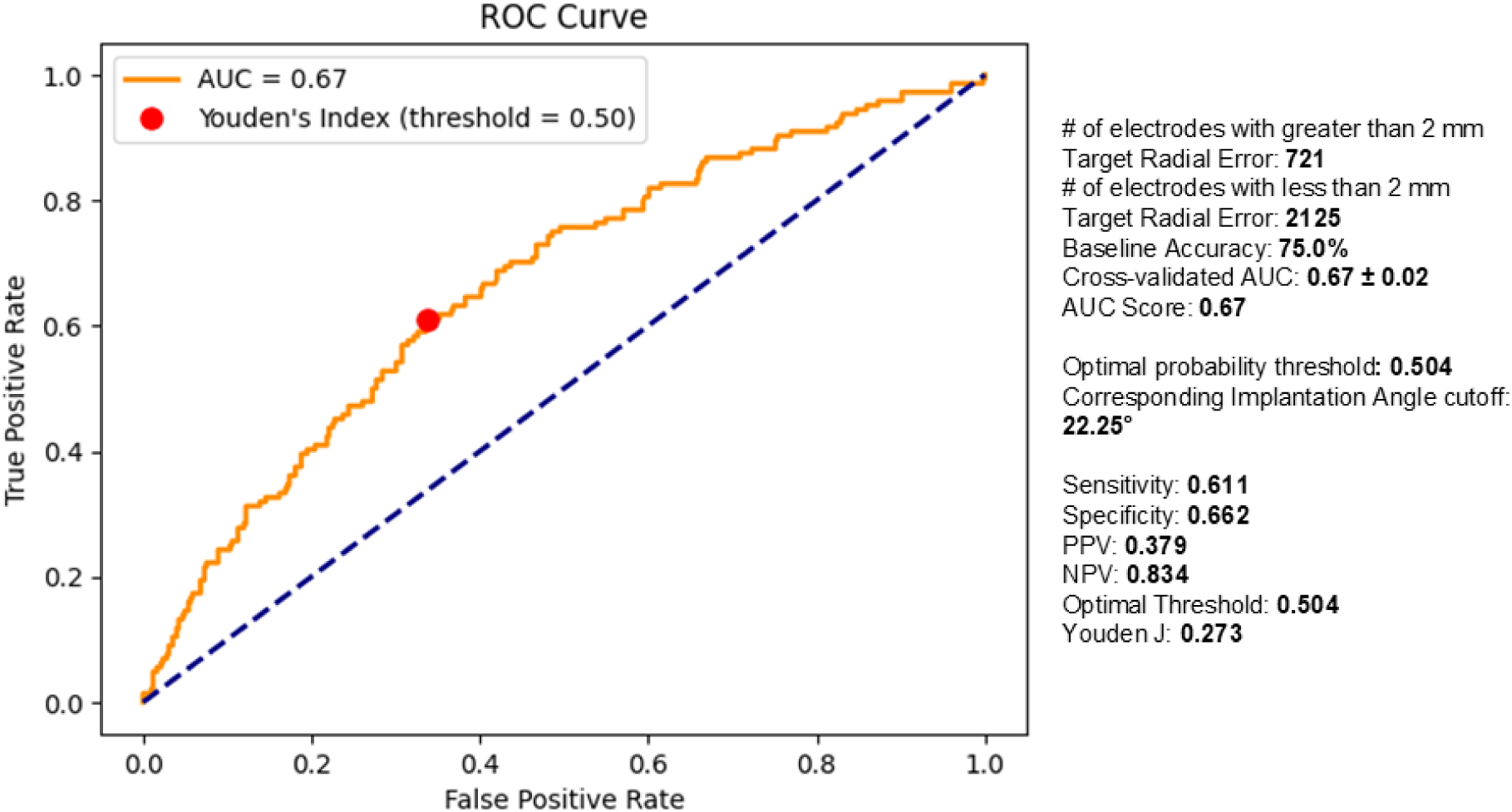
Receiver operating characteristic (ROC) curve for a logistic regression model predicting the probability of a trajectory exceeding 2 mm target radial error from implantation angle. The red point marks the optimal operating threshold by Youden’s J (probability 0.504, corresponding to an implantation-angle cutoff of 22.25°). Cross-validated AUC 0.67 ± 0.02; at this threshold, sensitivity 0.708, specificity 0.660, Youden’s J 0.273.

**Table 4.** SEEG application accuracy based on implantation angle.

|  | *<15°(n=840)* |  |  | *15°-30° (n=1552)* |  |  | *30°-45° (n=410)* |  |  | *>45° (n=32)* |  |  |
| --- | --- | --- | --- | --- | --- | --- | --- | --- | --- | --- | --- | --- |
| **Euclidean error (mm)** | **median (IQR)** | **mean ± sd** | **p-value** | **median (IQR)** | **mean ± sd** | **p-value** | **median (IQR)** | **mean ± sd** | **p-value** | **median (IQR)** | **mean ± sd** | **p-value** |
| Entry | 1.15 (0.72-1.67) | 1.34 ± 0.94 | <.0001 | 1.41 (0.94-1.99) | 1.58 ± 0.98 | .0787 | 1.79 (1.13-2.61) | 2.06 ± 1.34 | <.0001 | 2.47 (1.64-3.73) | 3.04 ± 2.0 | <.0001 |
| Target | 1.94 (1.34-2.65) | 2.07 ± 1.01 | <.0001 | 2.19 (1.56-2.93) | 2.35 ± 1.12 | .3261 | 2.53 (1.79-3.45) | 2.74 ± 1.33 | <.0001 | 3.22 (2.76-4.01) | 3.89 ± 2.12 | <.0001 |
| **Radial error (mm)** |  |  |  |  |  |  |  |  |  |  |  |  |
| Entry | 0.88 (0.52-1.30) | 1.04 ± 0.85 | <.0001 | 1.05 (0.66-1.51) | 1.17 ± 0.77 | .2241 | 1.31 (0.81-1.91) | 1.43 ± 0.9 | <.0001 | 1.46 (1.08-2.23) | 1.78 ± 1.03 | <.0005 |
| Target | 1.1 (0.69-1.63) | 1.24 ± 0.82 | <.0001 | 1.39 (0.90-2.04) | 1.57 ± 0.97 | .0114 | 1.86 (1.15-2.78) | 2.07 ± 1.26 | <.0001 | 2.44 (1.88-3.27) | 3.03 ± 2.11 | <.0001 |
| **Depth error (mm)** |  |  |  |  |  |  |  |  |  |  |  |  |
| Entry | 0.44 (0.17-0.89) | 0.65 ± 0.67 | <.0001 | 0.65 (0.27-1.22) | 0.88 ± 0.85 | <.05 | 0.92 (0.36-1.71) | 1.28 ± 1.26 | <.0001 | 2.56 (1.91- 3.2) | 2.28 ± 1.95 | <.0001 |
| Target | 1.33 (0.66-2.11) | 1.45 ± 0.99 | .9667 | 1.28 (0.63-2.09) | 1.48 ± 1.08 | .6778 | 1.35 (0.68-2.01) | 1.49 ± 1.1 | .7320 | 1.53 (0.75-2.20) | 1.8 ± 1.71 | .5250 |

### Lobar Accuracy Analysis

Kruskal-Wallis comparisons between the six lobes (Figure 5, Table S2, Supplementary Figure 4) reached significance for target Euclidean error (p<.01) and radial error (p<.05). Parietal lobe trajectories were the most accurate with a target Euclidean error of 2.0 mm (1.32-2.68, p<.05). Frontal lobe trajectories were the most inaccurate with a Euclidean error of 2.37 mm (1.66-3.11, p<.005). Insular (p=.80) and temporal lobe (p=.50) Euclidean error did not statistically differ. Repeating our logistic regression analysis, likelihood ratio testing of covariate and interaction effects models reached significance (p<.01) indicating that the relation between implantation angle and target radial error surpassing 2 mm varied between lobes. Cutpoints ranged from 33.9° for the frontal lobe (p<.005) and 60.8° for the parietal lobe (p<.05) (Supplementary Figure 5).

**Figure 5.**
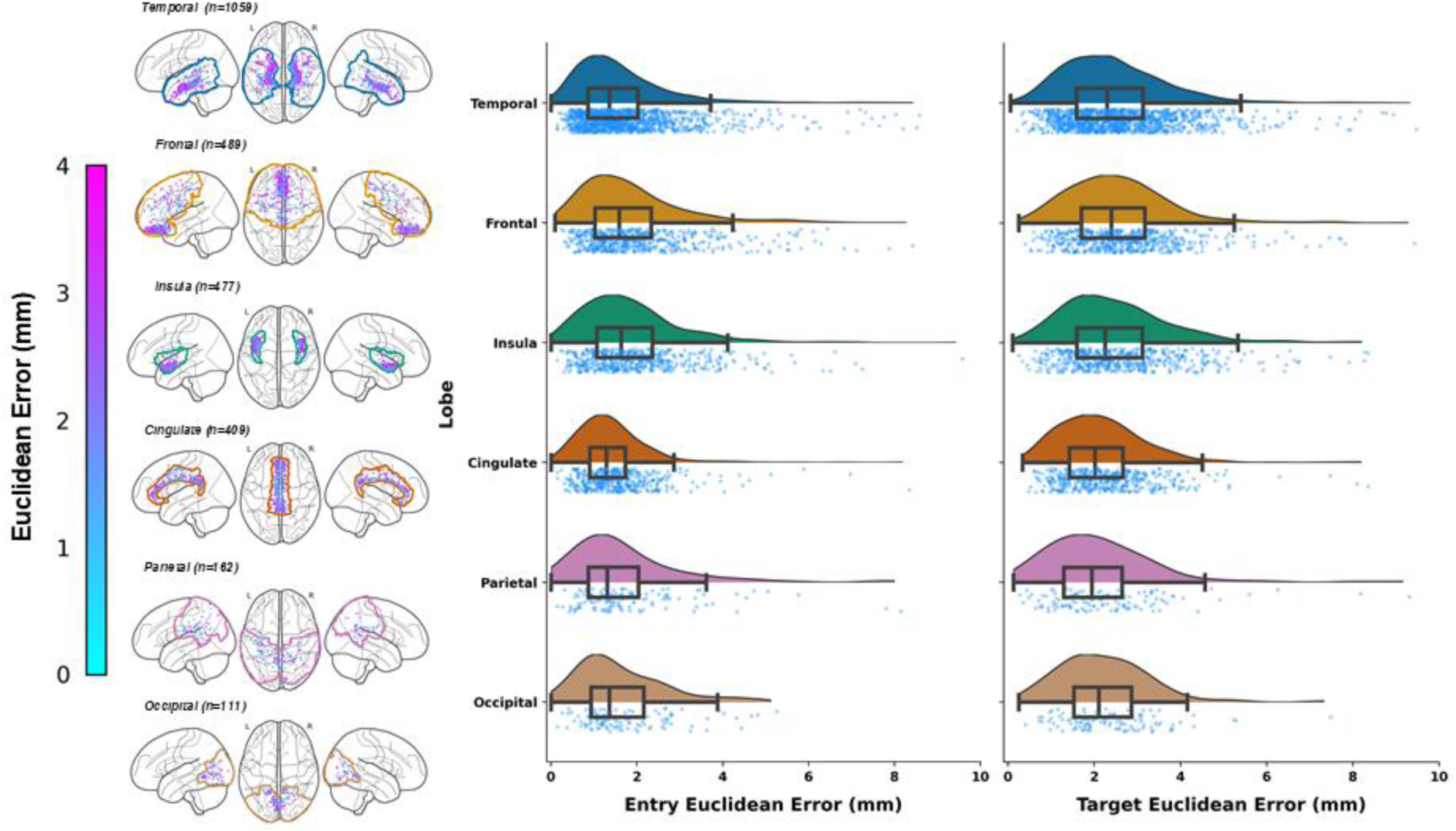
Error metrics by lobe implanted.

### Trajectory-Based Accuracy Analysis

We further compared the accuracy of the most frequent trajectories (n=1904), (Figure 6, Table S3, Supplementary Figure 6). Kruskal-Wallis comparison demonstrated that target radial error was statistically different among trajectories (p<.001). There was no statistical evidence that the relation between implantation angles and radial error exceeding 2 mm varied between trajectories following logistic regression analysis and likelihood testing.

**Figure 6.**
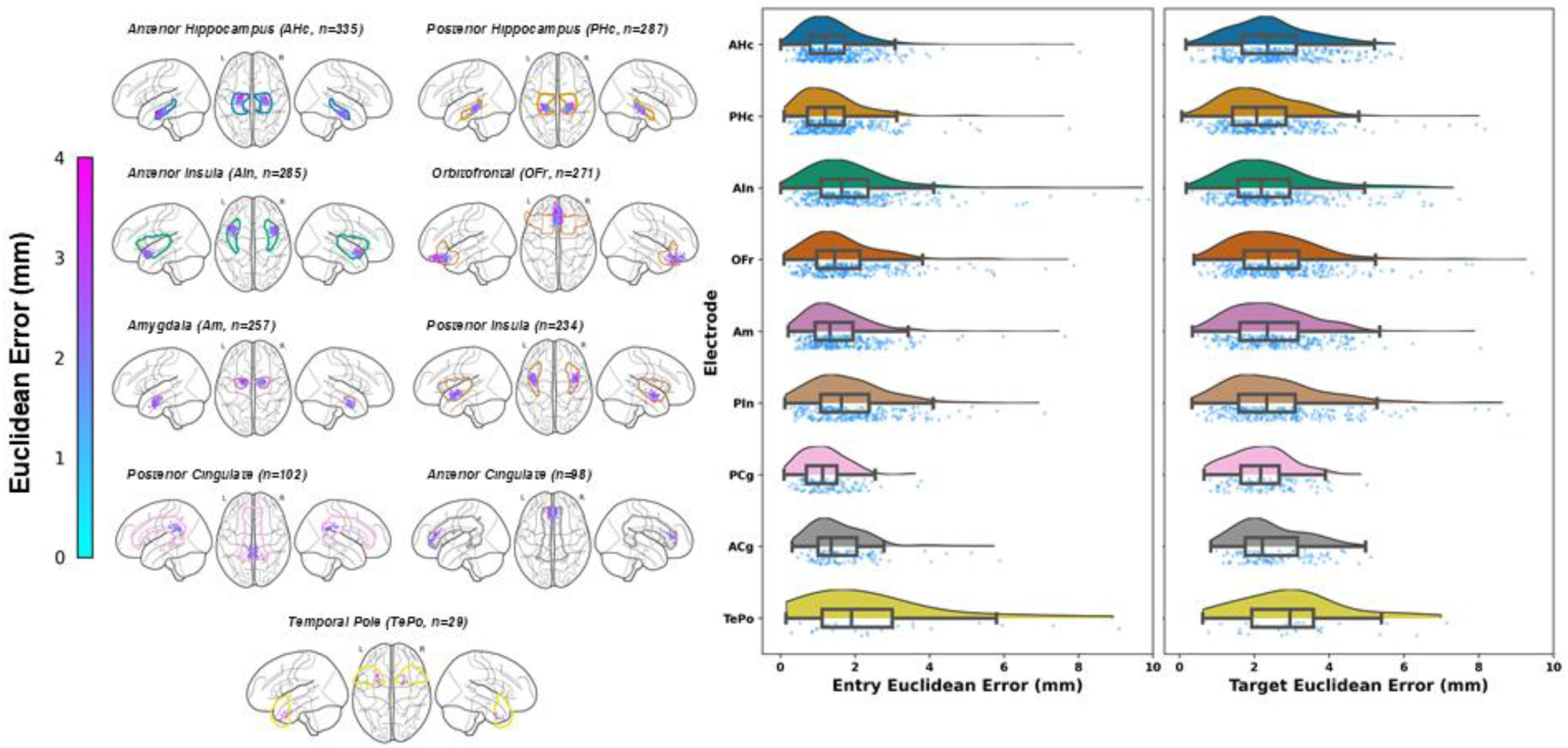
Error metrics by surgeon-labelled trajectory.

Within the temporal lobe (Figure 7), the posterior hippocampus had the lowest target radial error of 1.18 mm (0.73-1.71, p<.0005), followed by the amygdala and anterior hippocampus. The least accurate trajectory was the temporal pole at 3.28 mm (2.11-4.44, p<.05). To understand what factors may influence accuracy, we observed that posterior hippocampus trajectories had the most orthogonal angle at 10.40° (7.67°-14.74°, p<.0001). In contrast, temporal pole trajectories had the steepest angle at 33.92° (27.90°-37.86°, p<.0001). For patients with mesial temporal sclerosis (MTS), electrodes targeting a sclerotic hippocampus (n=88) had a greater target radial error of 1.58 mm (0.91–2.32) than a non-sclerotic hippocampus (n=54) at 1.12 mm (0.85–1.72; p<.05); Euclidean error did not differ (2.39 mm vs. 2.28 mm, p=.591). In the insula (Figure 8), orthogonal trajectories (n=15) had a lower target radial error of 1.07 mm (0.91-1.72) than oblique trajectories (n=525) at 1.64 (1.08-2.42, p<.05).

**Figure 7.**
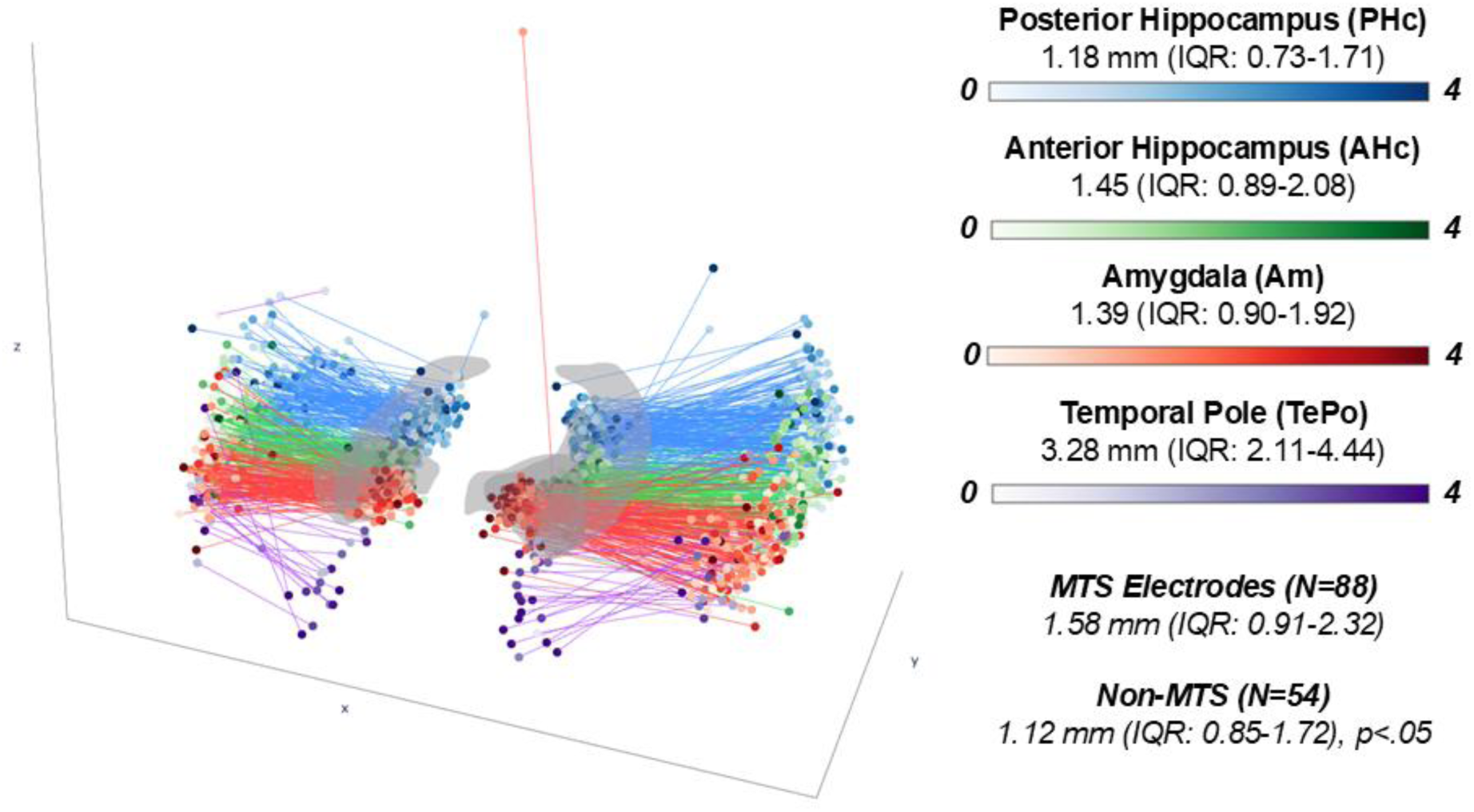
Trajectory-specific analysis of the temporal lobe in *MNI152NLin2009cSym* space highlighting target radial error.

**Figure 8.**
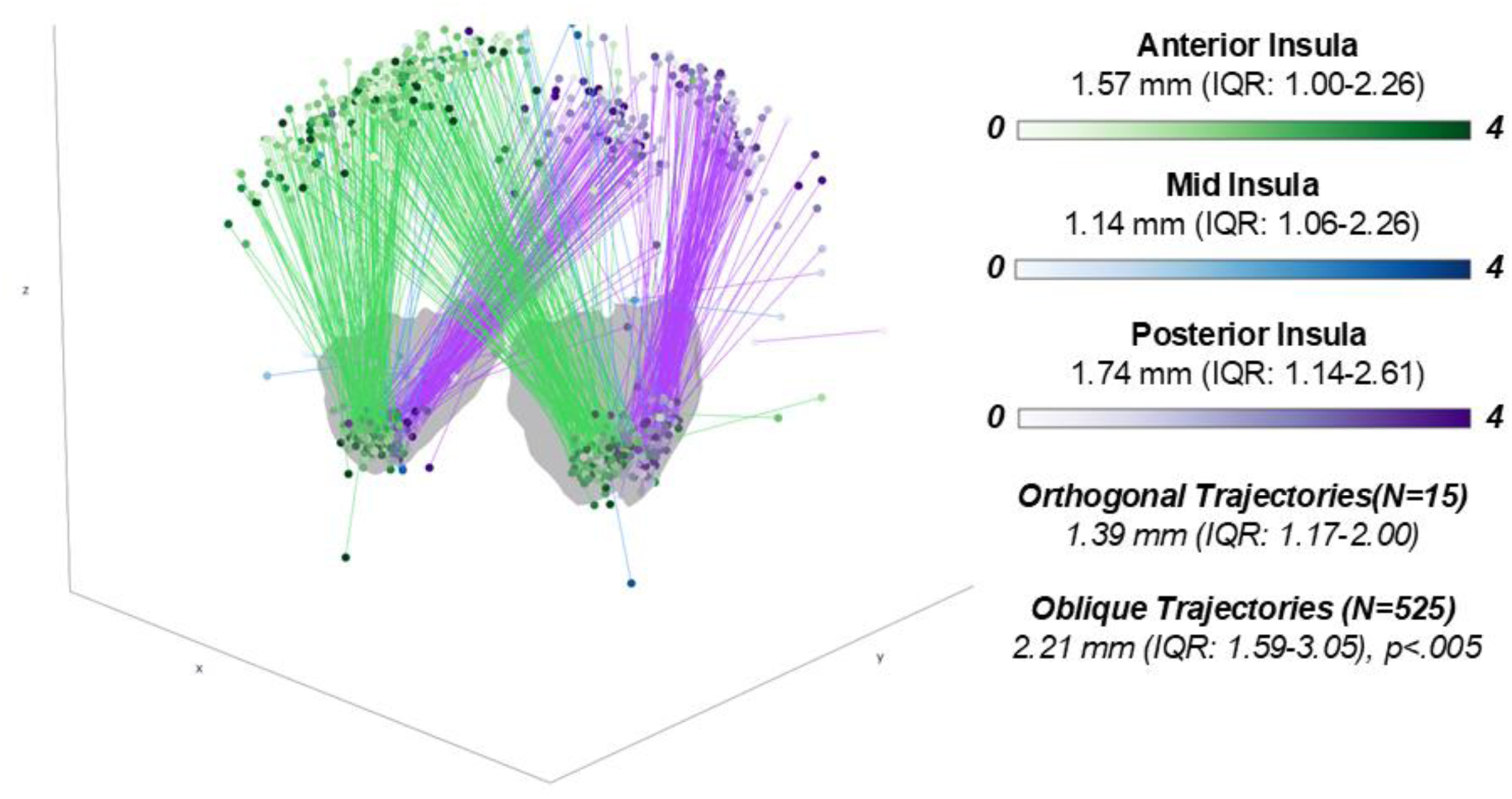
Trajectory-specific analysis of the insula in *MNI152NLin2009cSym space* highlighting target radial error.

## DISCUSSION

We report on the accuracy of a carefully curated series of SEEG implantations (N=3176), which to our knowledge represents the largest reported series to date. Beyond sample size, the detailed investigation of factors ranging from global to trajectory level distinguishes this work from prior studies. At a global level, we demonstrate that robot-assisted implantations are faster and more accurate than frame-based implantations, consistent with prior work^6–11^. Furthermore, implantation angle, scalp thickness, skull thickness, trajectory length, and obesity were positively correlated with target radial error. Entry errors were also correlated with target radial error reinforcing the importance of the entry point in stereotaxy. Trajectory-specific variables including the presence of mesial temporal sclerosis increased targeting error.

### Electrode accuracy in relation to implantation angle

Our statistical analysis showed that a 30° change to implantation angle increased radial error by close to a millimeter and determined cutpoint of 22.25° for predicting radial error greater than 2 mm. This has clinical relevance, as the implantation angle can be adjusted at the time of planning. Previous work has identified the implantation angle as a predictor of target error.^18,20–23^ Rollo et al. investigated the difference in accuracy between orthogonal and oblique (>30°) trajectories for 1987 electrodes and found no significant difference in target point error.^18^ Instead, we observed statistically significant differences in application accuracy at less oblique angles (<15°, 15-30°, and 30-45°).

### Lobar and trajectory-based analysis

The variance in accuracy across anatomical regions indicates that certain targets carry more risk of inaccuracy. Notably, we demonstrate the relation of implantation angle to target radial error varies by lobe, with frontal and parietal trajectories tolerable to steeper angles. Insular target errors did not differ statistically from other lobes supporting the efficacy of the oblique trajectory, which although less accurate than the orthogonal approach, permit more efficient sampling of the insula^44^. Finally, we noted that electrodes targeting sclerotic hippocampi deflected radially by 0.4 mm relative to non-sclerotic hippocampi, presumably due to tissue properties that increase electrode deviation.

### Reporting of SEEG implantation accuracy in the literature

The metrics reported in the literature for SEEG accuracy remain quite inconsistent. While most groups report Euclidean error, a 2017 systematic review identified that few groups (5/15) report radial error.^40^ Recent recommendations suggest using radial error as the primary outcome^41^ and for routine reporting of entry error metrics^42^, since hemorrhagic complications typically occur due to surface vessel incursion. Given the lack of clear guidelines, we opted to report on all commonly used error metrics, although for pragmatic reasons use target radial error as our endpoint in regression analysis.

There is also a lack of standardization regarding the choice of summary statistics for reporting stereotactic accuracy. In their 2013 study, Cardinale et al. observed that localization errors did not meet assumptions of normality and reported their findings using the median and IQR.^6^ Despite this, reporting in subsequent studies remains variable with many only providing the mean with standard deviation.^9,10,23,43^ Our analysis demonstrates that no accuracy metrics meet criteria for normality, compelling us to adopt median and IQR although we include mean and standard deviation to facilitate comparison with other groups.

Finally, although SEEG may not demand the same level of millimetric precision as other stereotactic procedures like deep brain stimulation^19^, optimizing accuracy remains clinically relevant. Knowledge of error-prone trajectories may help with complication avoidance, and radial inaccuracy can change SOZ interpretation. Furthermore, growing interest in subcortical targets and SEEG-guided RFTC necessitates increasingly accurate targeting.^44–46^

## LIMITATIONS

This is a single-center, retrospective analysis, and is subject to selection bias, incomplete charting, and lack of standardized follow-up. Frame-based implantation served as a historical control, introducing potential confounds related to changes in surgical workflow. Atlas-based labelling relied on registration to MNI space, which can be inaccurate but was mitigated by use of validated registration workflows and visual inspection.^33–35^

## CONCLUSIONS

We perform a comprehensive summary of implantation accuracy in over 3000 SEEG trajectories, using the best available recommendations from the literature. We demonstrate improved accuracy and efficiency with robotic over frame-based approaches. Entry error, implantation angle, scalp thickness, skull thickness, trajectory length, and obesity were found to be positively correlated to target error. We also discovered that targeting hippocampi in the presence of sclerosis led to increased error. By characterizing global and trajectory-level inaccuracies in SEEG this study establishes a framework for reporting stereotactic accuracy, which can further improve SEEG planning and implantation and be extended to other procedures.

## Data Availability

All data produced in the present study are available upon reasonable request to the authors

## Footnotes

* v1.3.0-alpha; https://github.com/pyushkevich/greedy

## Notes

### Competing Interest Statement

The authors have declared no competing interest.

### Author Declarations

This research received approval from the Institutional Review Board at the University of Western Ontario (REB# 107590) and London Health Sciences Centre (R-16-065).

