## Supplemental Materials for "Stereoelectroencephalography accuracy in a series of over 3000 trajectories"

### Supplementary Materials

#### MRI and CT Scanning Parameters

***1.5 T Signa General Electric (n=187),***
matrix size = 256 × 256
resolution

- 0.51×0.51×1.40 mm, (n=151)
- 1.02×1.02×1.40 mm, (n=30)
- 0.5×0.5×0.6 mm, (n=6)

***1.5 T Siemens MAGNETOM Sola (n=57)***
matrix size = 464 × 512
resolution

- 0.5x0.5x1mm, (n=55)
- 0.57x.0.57x1mm, (n=1)
- 0.45x0.45x0.9, (n=1)

***3T Siemens MAGNETOM Vida (n=16)***
matrix size = 180 × 230
resolution

- 0.94x0.94x0.9mm, (n=9)
- 0.9x0.9x0.9mm, (n=3)
- 0.8x0.8x0.8mm, (n=1)
- 0.5x.0.5x1, (n=2)
- 1x1x1, (n=1)

***CT Scanning Parameters***
120 kV, 145 mA, 1,331 mm acquisition diameter, 320 mm reconstruction diameter, 512×512 matrix, 0.625×0.625 $mm^2$ pixel spacing, 96 axial slices, 0.625 mm isotropic, 0° gantry tilt

#### Non-deformable registration parameters

***affine registration parameters***

- initial alignment using image centres
- normalized mutual information as the image similarity optimization metric
- registration iterations of 100, 50, and 1

**Supplementary Table 1.** Implantation accuracy and variables for LHSC SEEG cohort (n=3167)

| | **Robot (2858 electrodes)** | | | | **Frame-based (309 electrodes)** | | | |
|---|---|---|---|---|---|---|---|---|
| ***Metric/Variable*** | ***median (IQR)*** | **mean ± sd** | **min** | **max** | ***median (IQR)*** | **mean ± sd** | **min** | **max** |
| **Euclidean error (mm)** | | | | | | | | |
| Entry | 1.38 (0.89-2.01) | 1.69 ± 1.73 | 0.00 | 32.98 | 2.21 (1.42-3.32) | 2.67 ± 2.23 | 0.00 | 22.30 |
| Target | 2.19 (1.54-2.98) | 2.46 ± 2.02 | 0.06 | 54.06 | 2.76 (1.79-3.76) | 3.16 ± 2.17 | 0.25 | 19.55 |
| **Radial error (mm)** | | | | | | | | |
| Entry | 1.04 (0.64-1.54) | 1.26 ± 1.53 | 0.00 | 32.97 | 1.87 (1.19-2.68) | 2.16 ± 1.68 | 0.00 | 19.31 |
| Target | 1.35 (0.85-2.02) | 1.63 ± 1.59 | 0.00 | 51.76 | 2.17 (1.23-3.14) | 2.48 ± 2.03 | 0.08 | 19.43 |
| **Depth error (mm)** | | | | | | | | |
| Entry | 0.62 (0.24-1.23) | 0.91 ± 1.04 | 0.00 | 14.91 | 0.88 (0.34-1.59) | 1.25 ± 1.73 | 0.00 | 20.98 |
| Target | 1.32 (0.66-2.11) | 1.55 ± 1.60 | 0.00 | 44.30 | 1.16 (0.55-1.93) | 1.49 ± 1.48 | 0.01 | 10.48 |
| **Radial Angle (°)** | 1.58 (0.98-2.38) | 1.92 ± 1.93 | 0.00 | 62.51 | 2.80 (1.61-4.32) | 3.42 ± 2.97 | 0.14 | 28.82 |
| **Line Angle (°)** | 1.01 (0.63-1.54) | 1.32 ± 1.80 | 0.00 | 63.64 | 1.68 (1.04-2.85) | 2.40 ± 3.36 | 0.01 | 42.88 |
| **Implantation Angle (°)** | 20.30 (13.46-26.74) | 20.65 ± 9.51 | 0.54 | 62.28 | 21.37 (14.28-28.53) | 21.58 ± 10.44 | 2.03 | 55.75 |
| **Trajectory Length (mm)** | 48.00 (42.10-58.98) | 52.73 ± 18.26 | 4.50 | 122.40 | 44.70 (35.75-51.58) | 46.03 ± 16.55 | 14.60 | 113.20 |
| **Scalp-Dura Distance (mm)** | 14.70 (12.50-17.40) | 15.18 ± 3.70 | 5.90 | 33.00 | 14.56 (11.98-17.33) | 15.07 ± 4.32 | 1.82 | 31.75 |
| **Skull Thickness (mm)** | 4.36 (2.48-6.90) | 4.89 ± 2.94 | 0.46 | 18.08 | 4.06 (2.71-6.28) | 4.85 ± 3.03 | 1.08 | 20.63 |
| **Scalp Thickness (mm)** | 9.77 (8.20-11.87) | 10.29 ± 3.04 | 0.06 | 30.76 | 9.55 (7.87-12.14) | 10.22 ± 3.79 | 26.60 | 0.02 |
| **Weight (kg)** | 76.10 (65.77-90.00) | 80.11 ± 22.00 | 40.70 | 167.40 | 71.60 (61.50-100.00) | 80.08 ± 22.19 | 52.60 | 128.10 |
| **CSF Volume ($mm^3$)** | 65,003 (40,948 – 92,850) | 80053.64 ± 61318.97 | 3760 | 384679 | 80,098 (57,690-116,597) | 95086.1 ± 69,659 | 12165.0 | 330683.0 |
| **Number of Electrodes** | 13.00 (11.00-15.00) | 13.02 ± 3.07 | 6.00 | 21.00 | 11.00 (9.00-12.00) | 10.23 ± 2.31 | 6.00 | 16.00 |
| **Procedure Duration (min)** | 90.00 (73.00-112.00) | 94.43 ± 31.01 | 19.00 | 230.00 | 133.00 (119.00-166.00) | 143.15 ± 31.06 | 87.00 | 202.00 |
| **Anesthesia Duration (min)** | 164.00 (140.00-187.00) | 170.32 ± 44.90 | 84.00 | 362.00 | 226.00 (188.00-250.00) | 225.19 ± 41.57 | 144.00 | 320.00 |

**Supplementary Table 2.** Error metrics by lobe implanted

| Region | Target | | Entry | | N |
|---|---|---|---|---|---|
| | Euclidean (mm) | Radial (mm) | Euclidean (mm) | Radial (mm) | |
| **Temporal** | 2.24 (1.58-3.05) | 1.41 (0.88-2.07) | 1.42 (0.93-2.09) | 1.11 (0.69-1.60) | 1219 |
| **Frontal** | 2.38 (1.66-3.12) | 1.48 (0.92-2.20) | 1.49 (0.91-2.19) | 1.09 (0.66-1.57) | 550 |
| **Insula** | 2.17 (1.60-2.89) | 1.4 (0.89-1.93) | 1.31 (0.87-1.99) | 0.97 (0.54-1.44) | 515 |
| **Cingulate** | 2.07 (1.48-2.92) | 1.27 (0.76-2.03) | 1.36 (0.91-1.95) | 1.04 (0.65-1.51) | 428 |
| **Parietal** | 2.0 (1.32-2.68) | 1.2 (0.80-1.90) | 1.34 (0.86-2.00) | 0.86 (0.50-1.39) | 127 |
| **Occipital** | 2.26 (1.56-2.98) | 1.27 (0.78-2.07) | 1.42 (1.04-2.17) | 1.17 (0.69-1.72) | 110 |

**Supplementary Table 3.** Implantation accuracy for surgeon-labeled trajectory in robot-assisted SEEG cohort (n = 2858)

| electrode | Target | | Entry | | N |
|---|---|---|---|---|---|
| | Euclidean (mm) | Radial (mm) | Euclidean (mm) | Radial (mm) | |
| **Anterior Hippocampus (AHc)** | 2.36 (1.68-3.17) | 1.62 (1.52, 1.71) | 1.21 (0.79-1.72) | 1.03 (0.60-1.43) | 337 |
| **Posterior Hippocampus (PHc)** | 2.08 (1.42-2.90) | 1.18 (0.73-1.71) | 1.19 (0.72-1.72) | 0.87 (0.56-1.39) | 291 |
| **Anterior Insula (AIn)** | 2.19 (1.58-2.98) | 1.58 (1.00-2.27) | 1.65 (1.08-2.36) | 1.19 (0.70-1.85) | 289 |
| **OrbitoFrontal (OFr)** | 2.38 (1.73-3.19) | 1.65 (1.09-2.20) | 1.47 (0.98-2.13) | 1.15 (0.72-1.61) | 273 |
| **Amygdala (Am)** | 2.36 (1.61-3.18) | 1.39 (0.90-1.94) | 1.33 (0.93-1.96) | 1.10 (0.69-1.56) | 259 |
| **Posterior Insula (PIn)** | 2.37 (1.58-3.16) | 1.74 (1.14-2.63) | 1.65 (1.08-2.41) | 1.07 (0.60-1.55) | 238 |
| **Posterior Cingulate (PCg)** | 2.22 (1.64-2.74) | 1.06 (0.66-1.70) | 1.14 (0.69-1.53) | 0.78 (0.51-1.19) | 104 |
| **Anterior Cingulate (ACg)** | 2.21 (1.76-3.15) | 1.35 (0.98-2.08) | 1.35 (1.01-2.04) | 1.18 (0.86-1.61) | 98 |
| **Temporal Pole (TePo)** | 2.96 (2.07-3.62) | 2.49 (1.53-3.14) | 1.96 (1.15-3.31) | 1.44 (0.89-2.27) | 29 |

**Supplementary Figure 1.** Inter-rater reliability of entry angle estimation

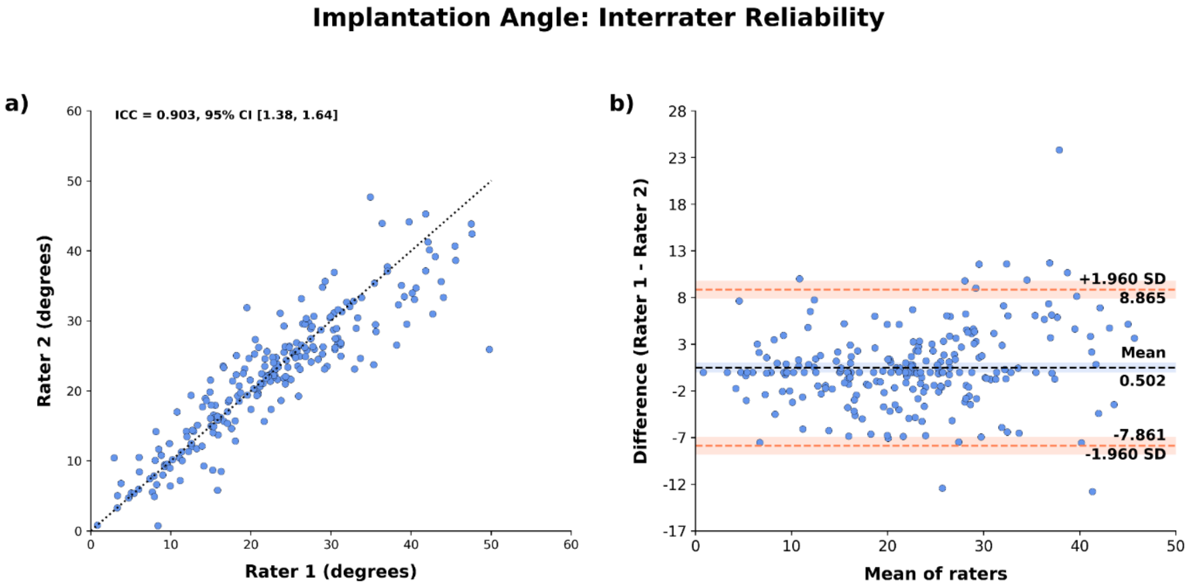

**Supplementary Figure 2.** Q-Q Plots and distributions of implantation variables

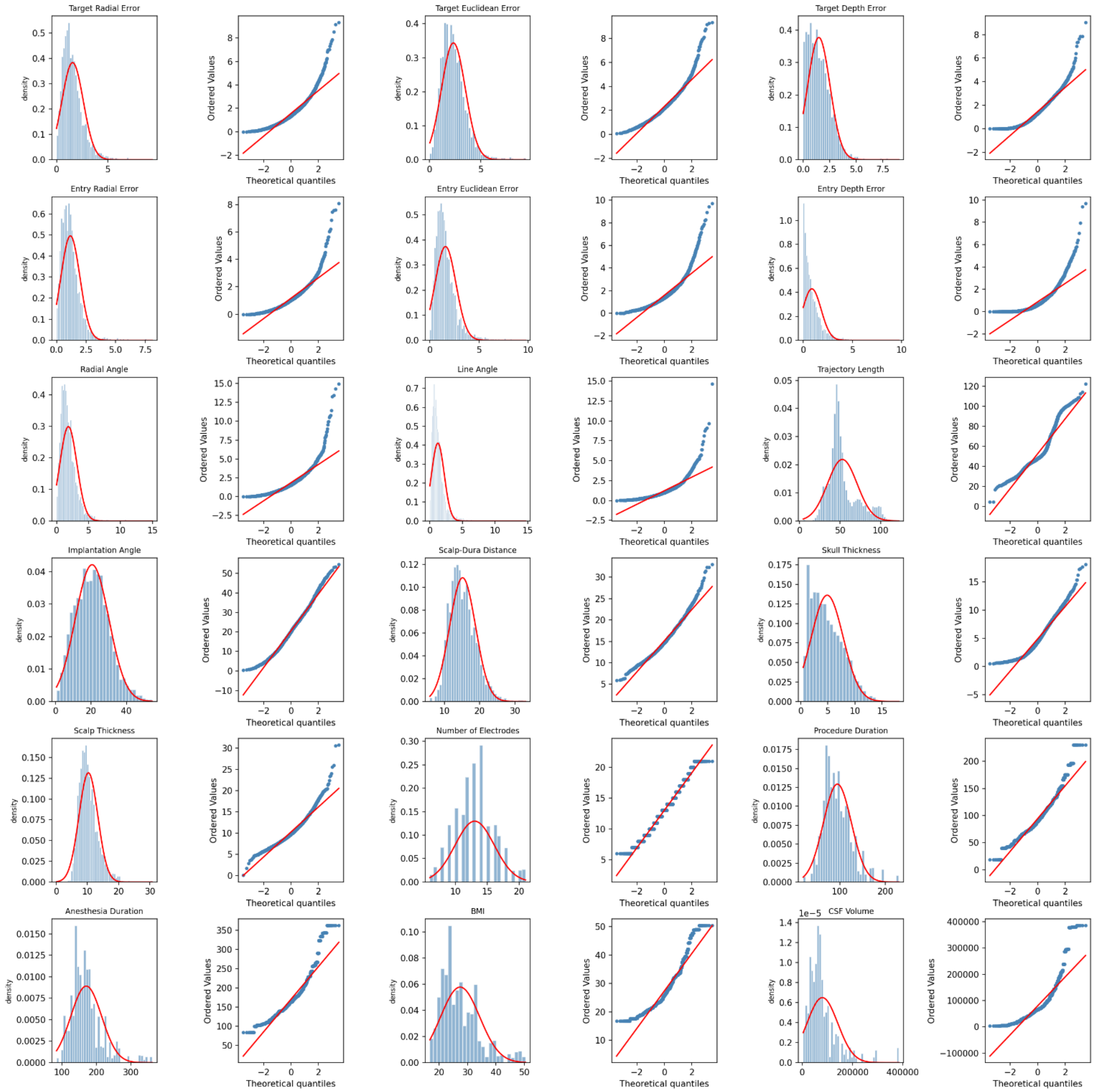

**Supplementary Figure 3.** Individual regression plots of implantation variables to target radial error

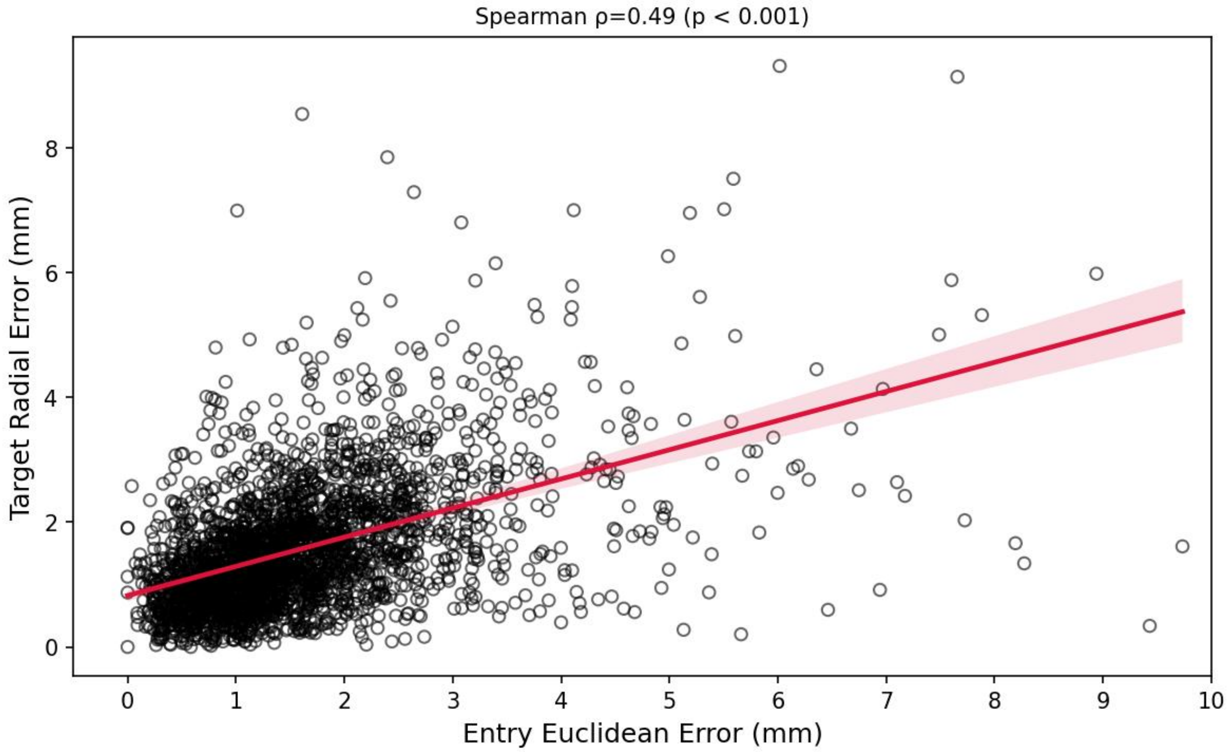

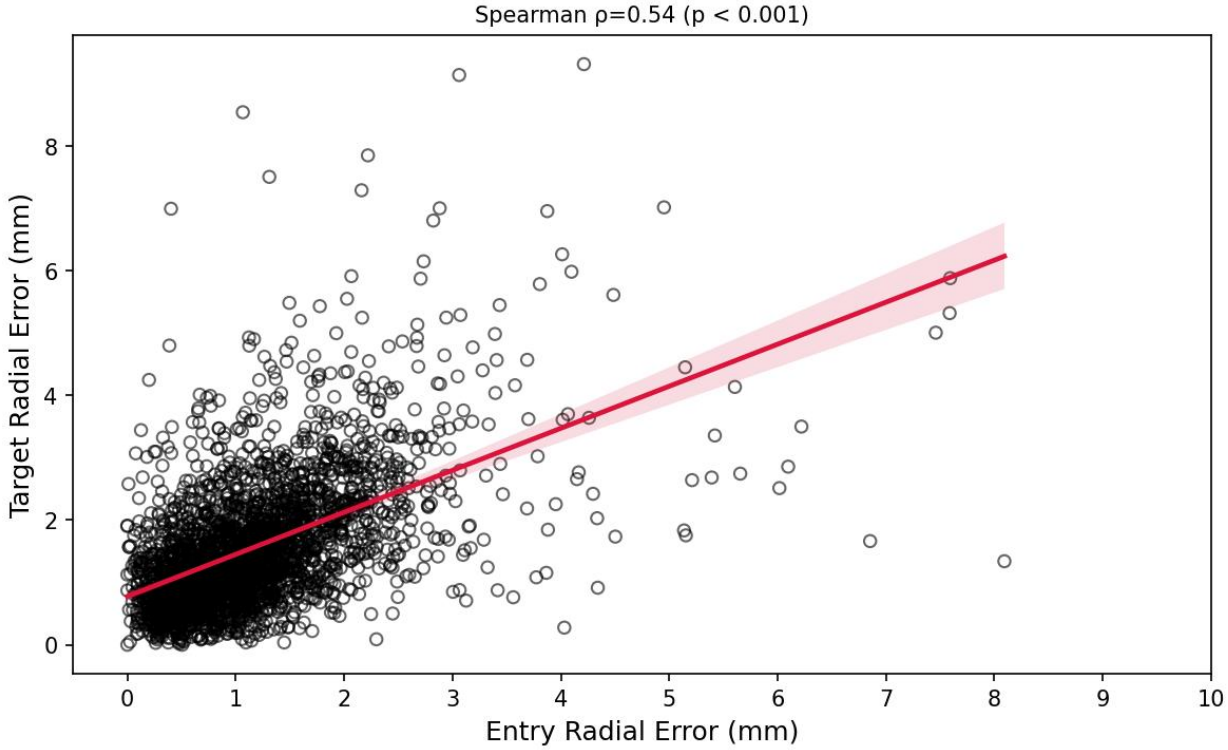

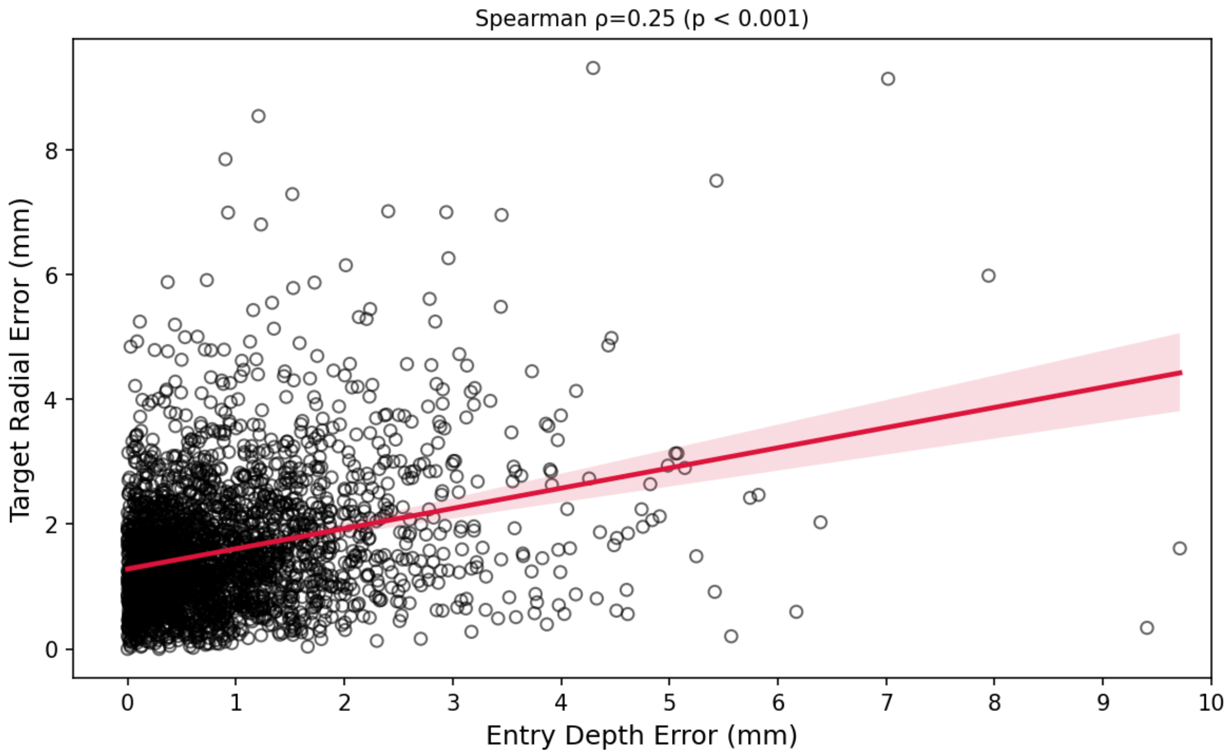
Spearman ρ=0.25 (p < 0.001)
Target Radial Error (mm)
Entry Depth Error (mm)
0
2
4
6
8
0
1
2
3
4
5
6
7
8
9
10

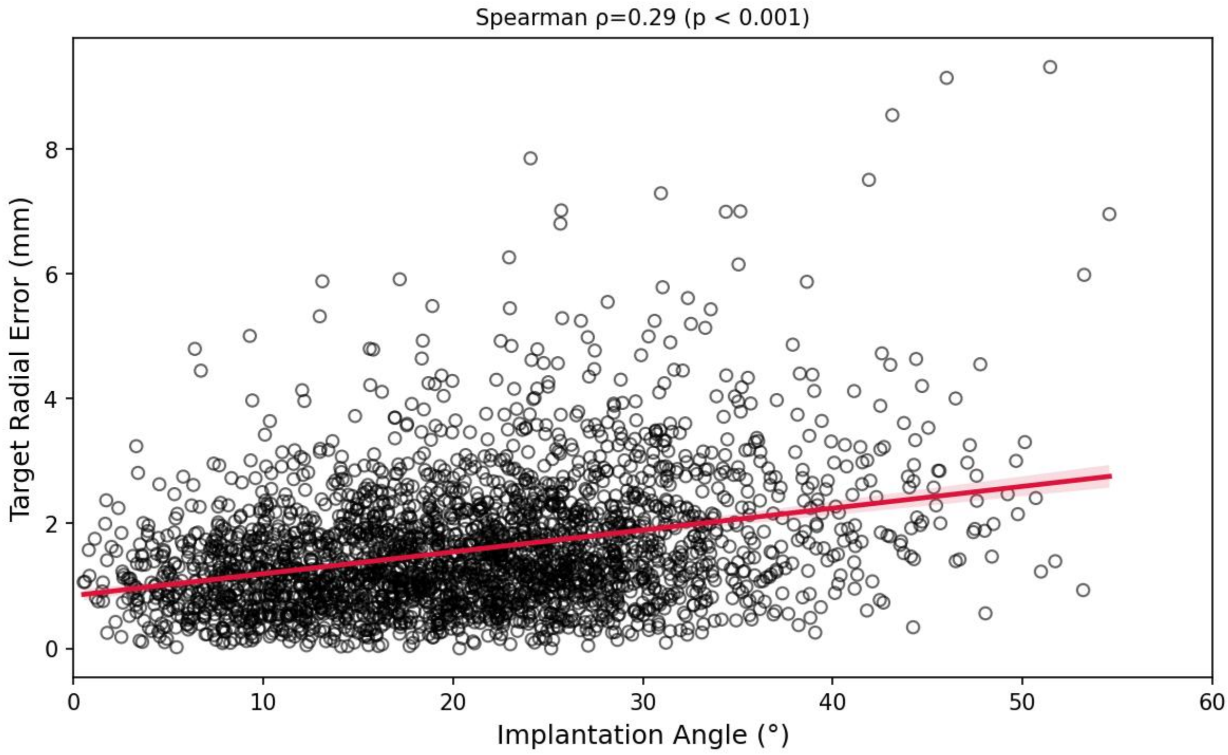
Spearman ρ=0.29 (p < 0.001)
Target Radial Error (mm)
Implantation Angle (°)
0
2
4
6
8
0
10
20
30
40
50
60

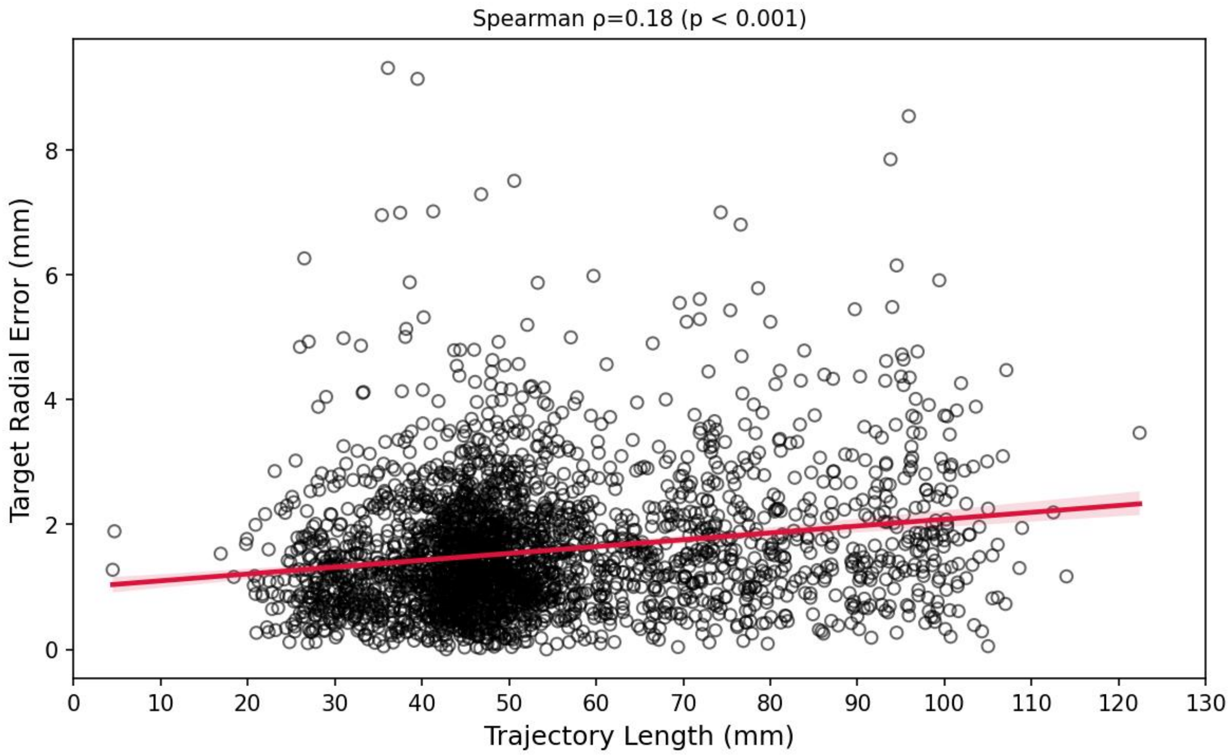
Spearman ρ=0.18 (p < 0.001)
Target Radial Error (mm)
0
2
4
6
8
Trajectory Length (mm)
0
10
20
30
40
50
60
70
80
90
100
110
120
130

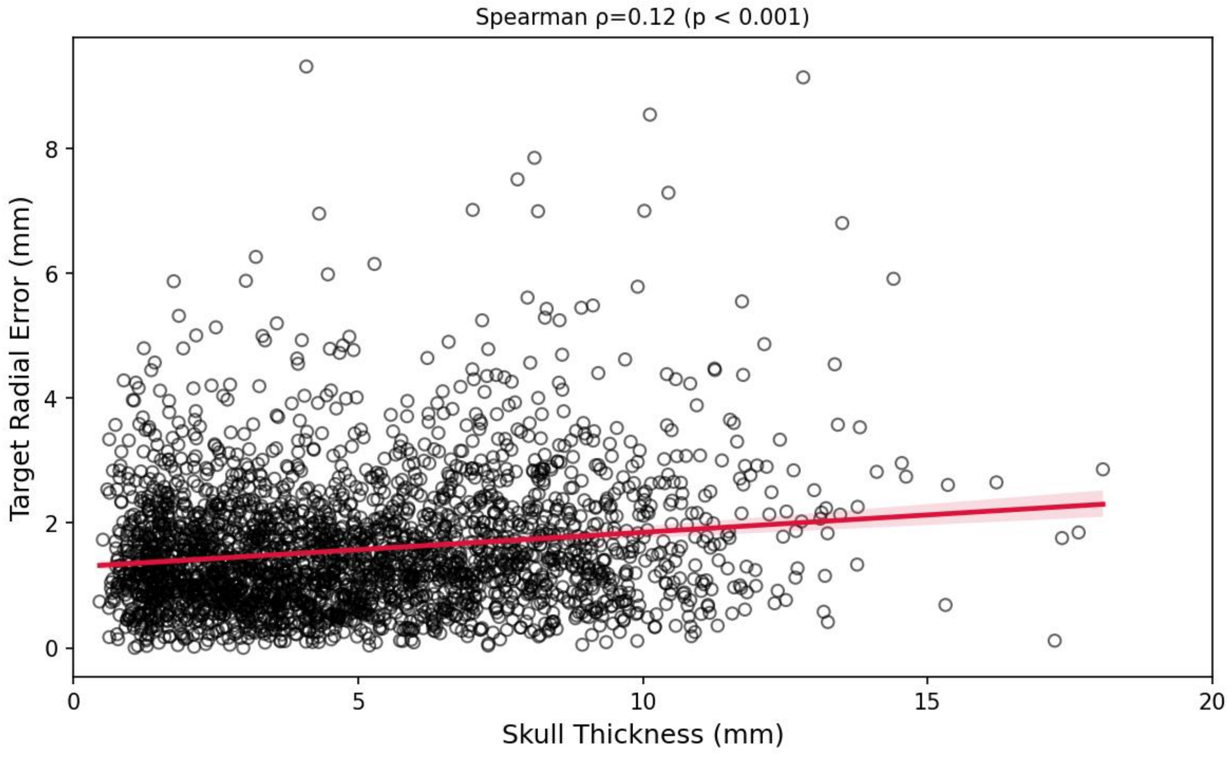
Spearman ρ=0.12 (p < 0.001)
Target Radial Error (mm)
0
2
4
6
8
Skull Thickness (mm)
0
5
10
15
20

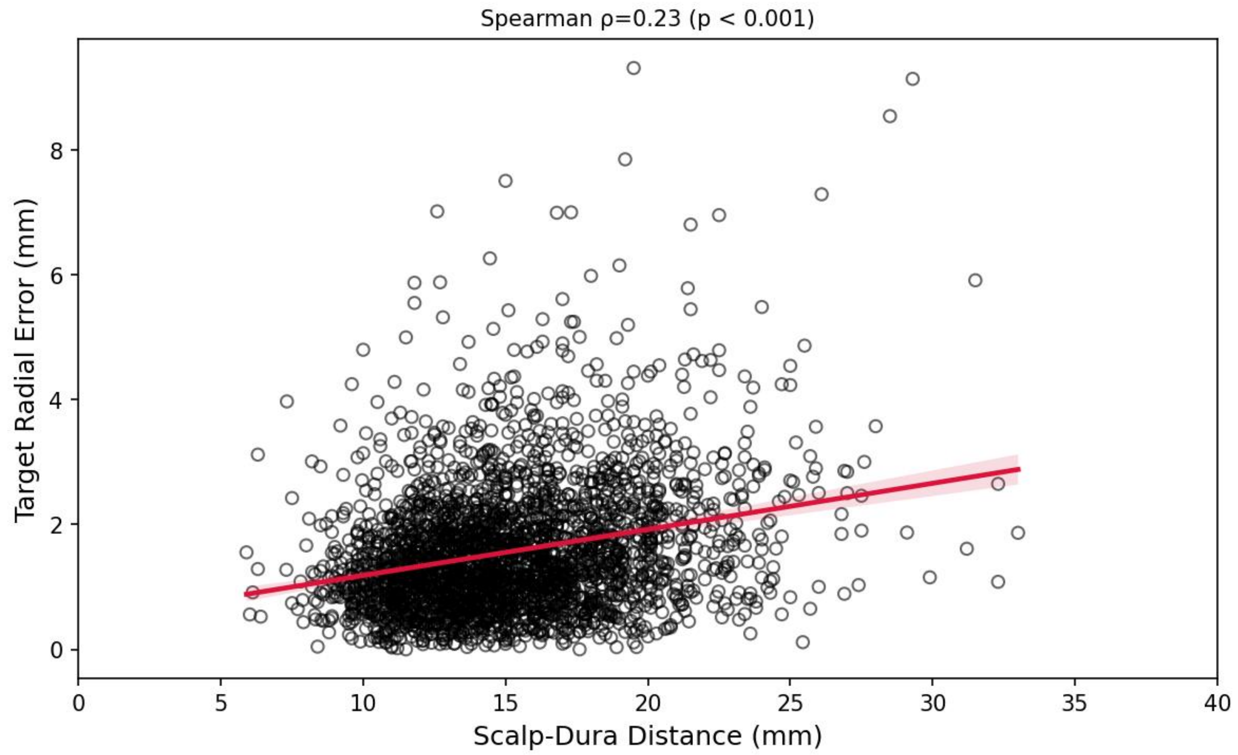

Spearman ρ=0.23 (p < 0.001)
Target Radial Error (mm)
0
2
4
6
8
0
5
10
15
20
25
30
35
40
Scalp-Dura Distance (mm)

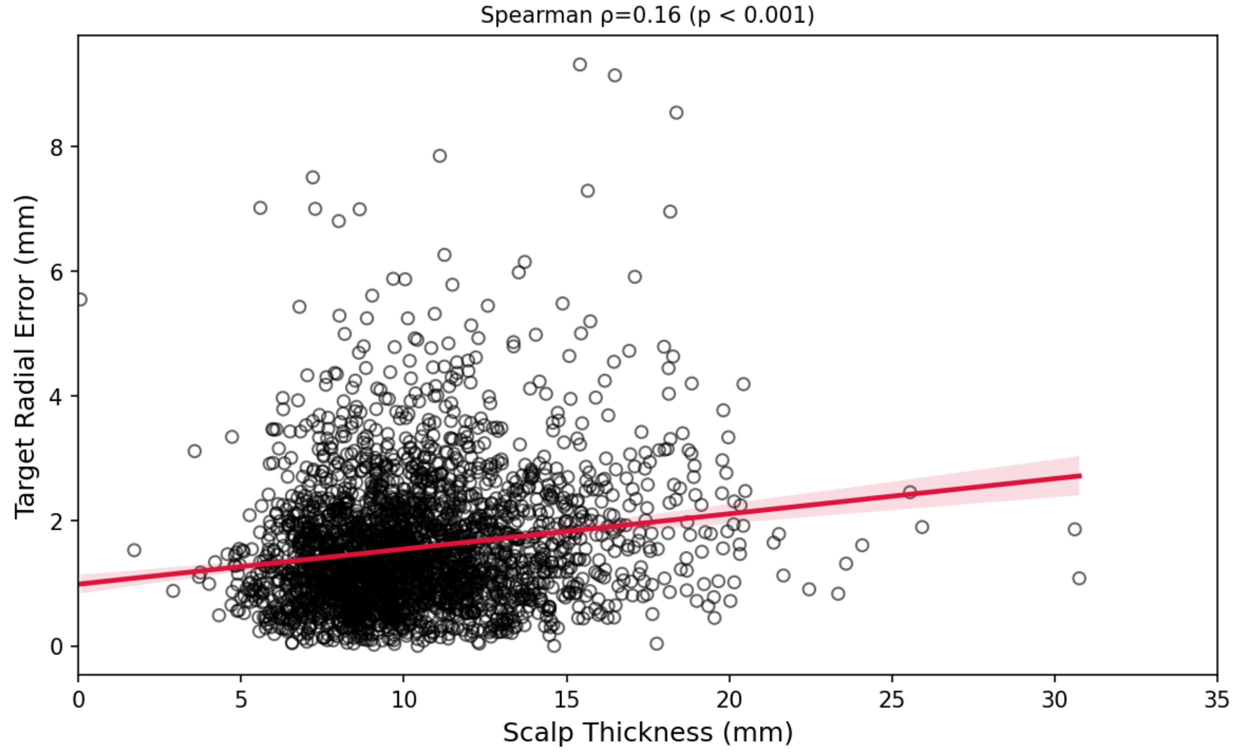

Spearman ρ=0.16 (p < 0.001)
Target Radial Error (mm)
0
2
4
6
8
0
5
10
15
20
25
30
35
Scalp Thickness (mm)

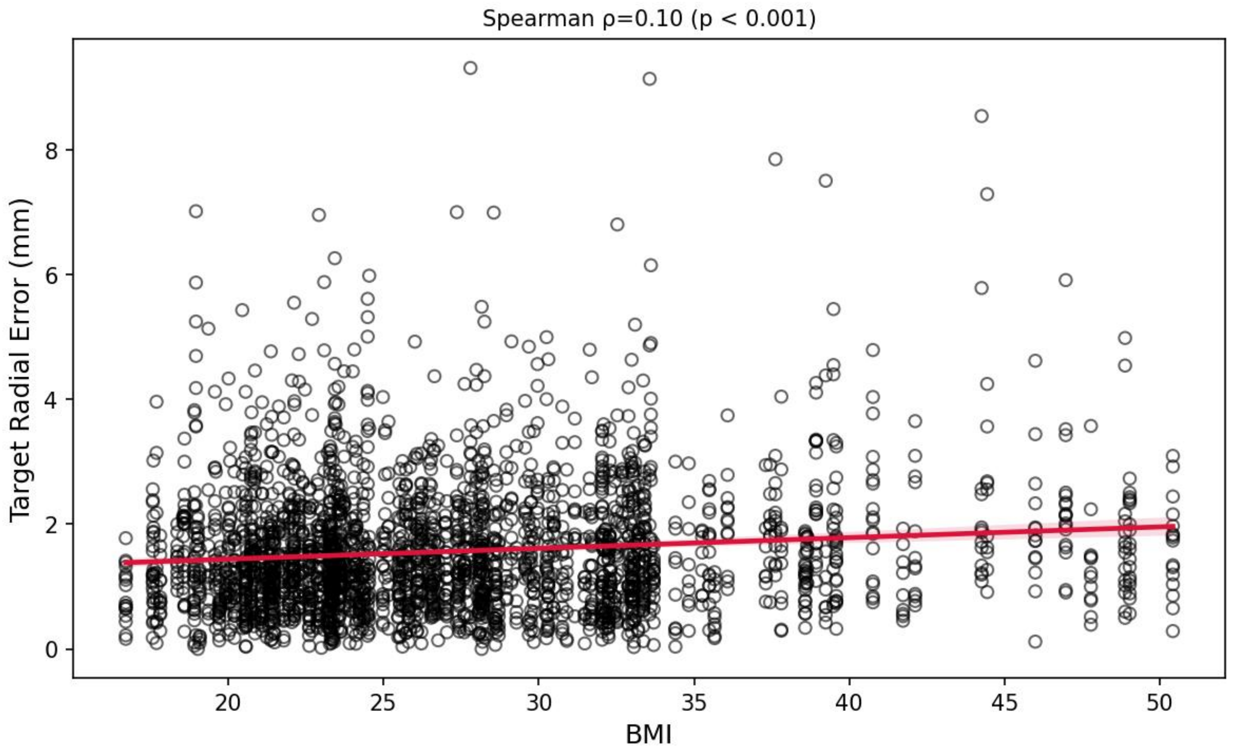

Spearman ρ=0.10 (p < 0.001)
Target Radial Error (mm)
BMI
0
2
4
6
8
20
25
30
35
40
45
50

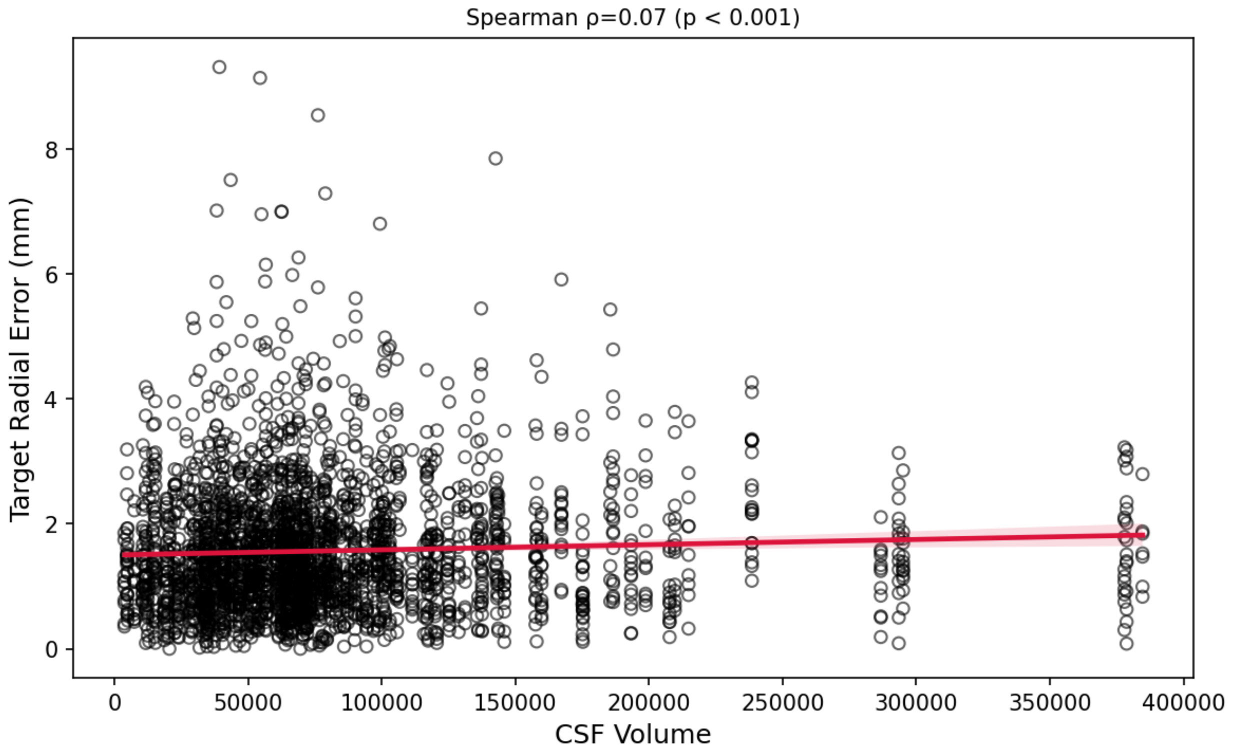

Spearman ρ=0.07 (p < 0.001)
Target Radial Error (mm)
CSF Volume
0
2
4
6
8
0
50000
100000
150000
200000
250000
300000
350000
400000

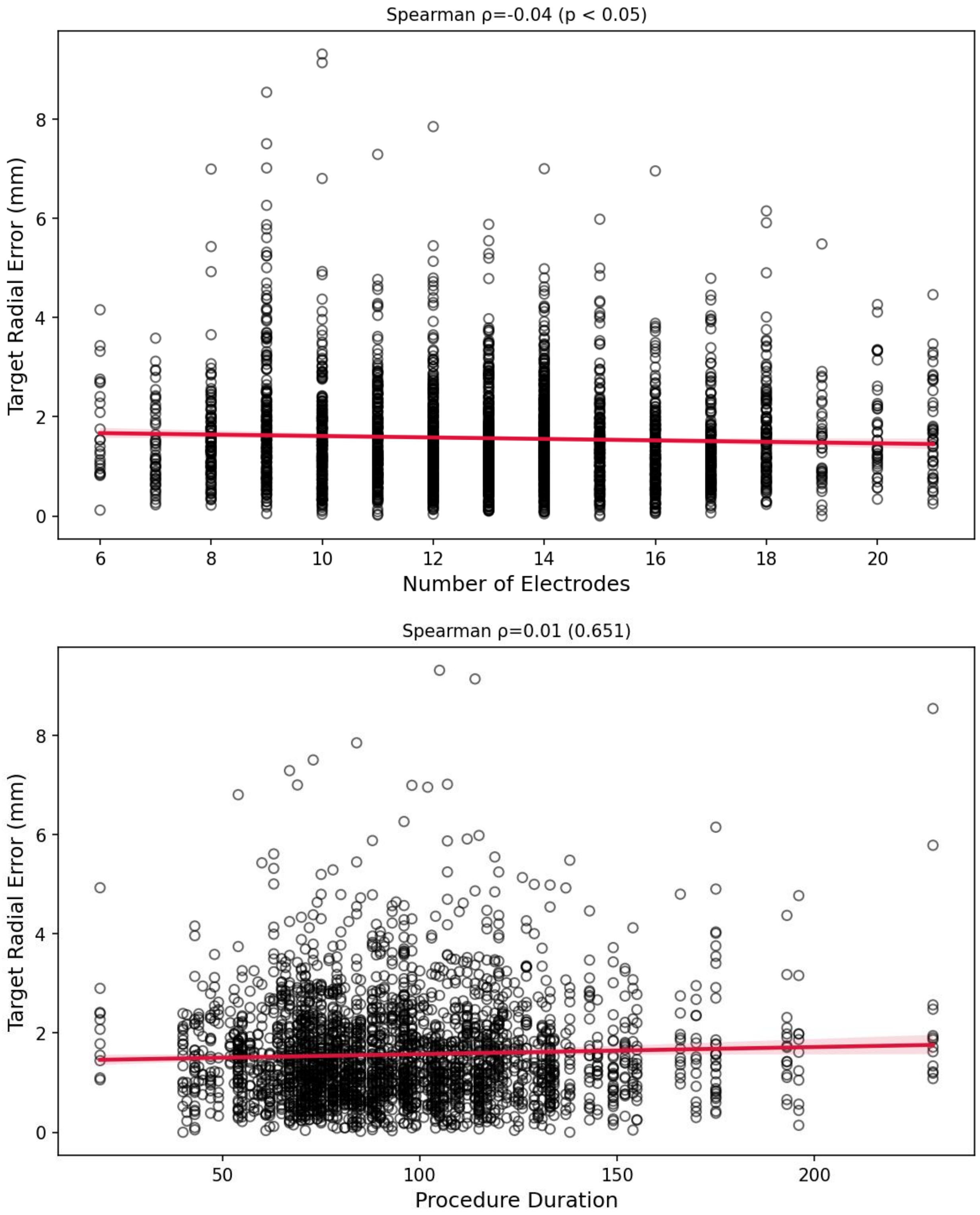
Spearman ρ=-0.04 (p < 0.05)
Target Radial Error (mm)
0
2
4
6
8
6
8
10
12
14
16
18
20
Number of Electrodes
Spearman ρ=0.01 (0.651)
Target Radial Error (mm)
0
2
4
6
8
50
100
150
200
Procedure Duration

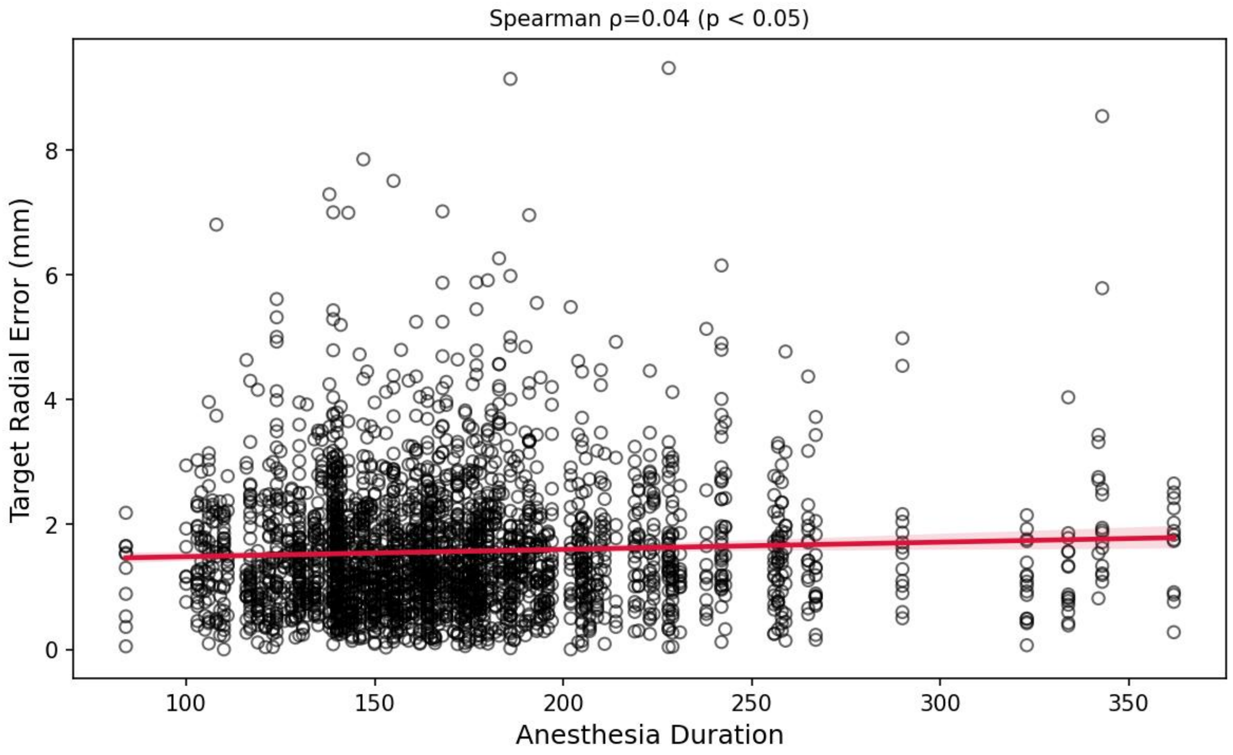
Spearman ρ=0.04 (p < 0.05)
Target Radial Error (mm)
Anesthesia Duration
0
2
4
6
8
100
150
200
250
300
350

**Supplementary Figure 3.** Spearman correlation by implanted lobe.

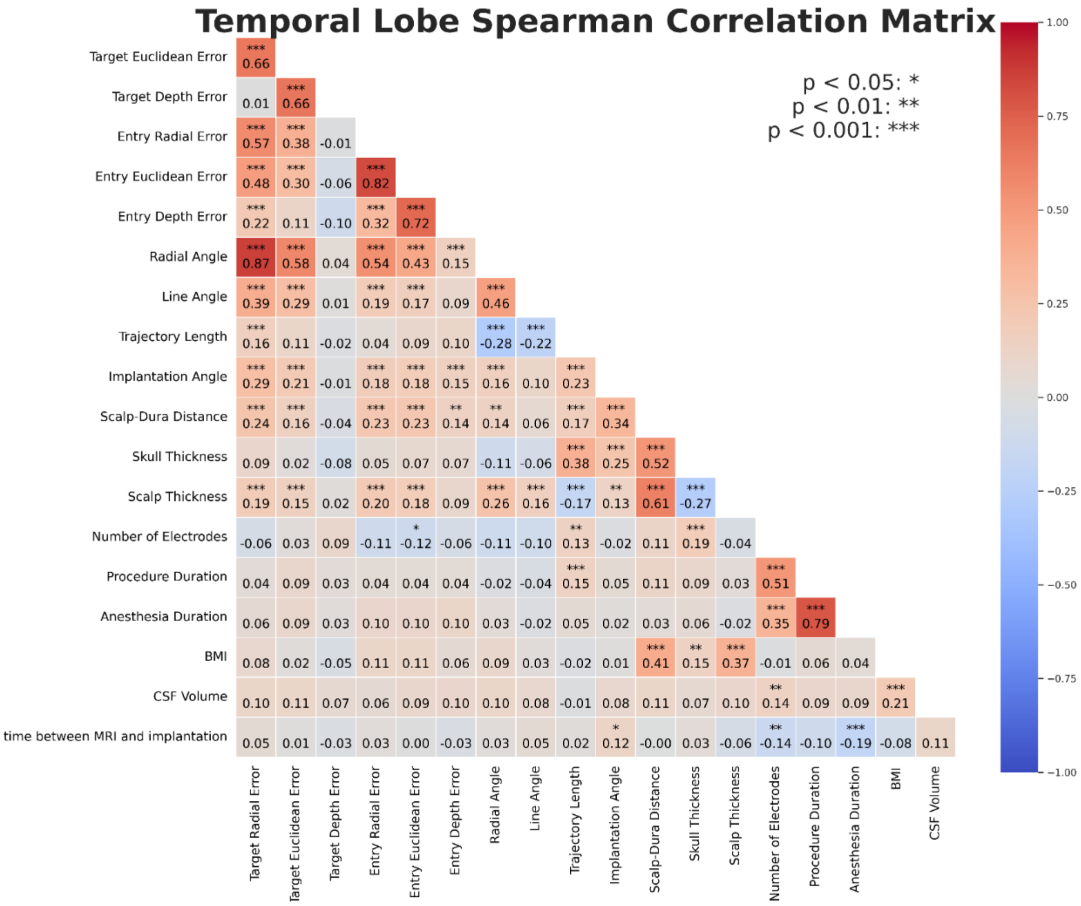

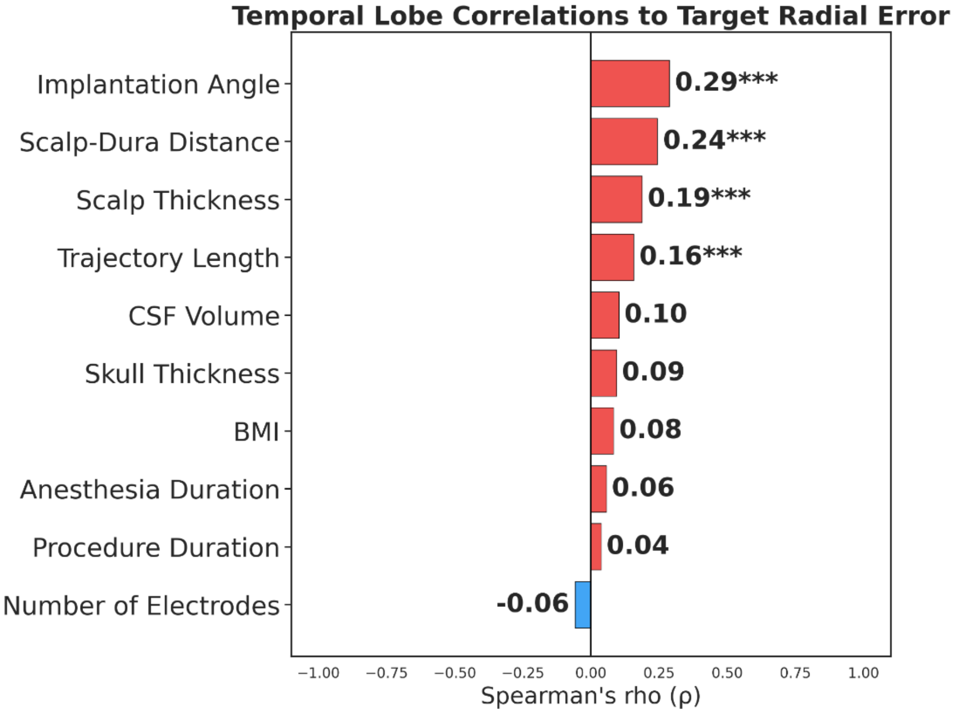

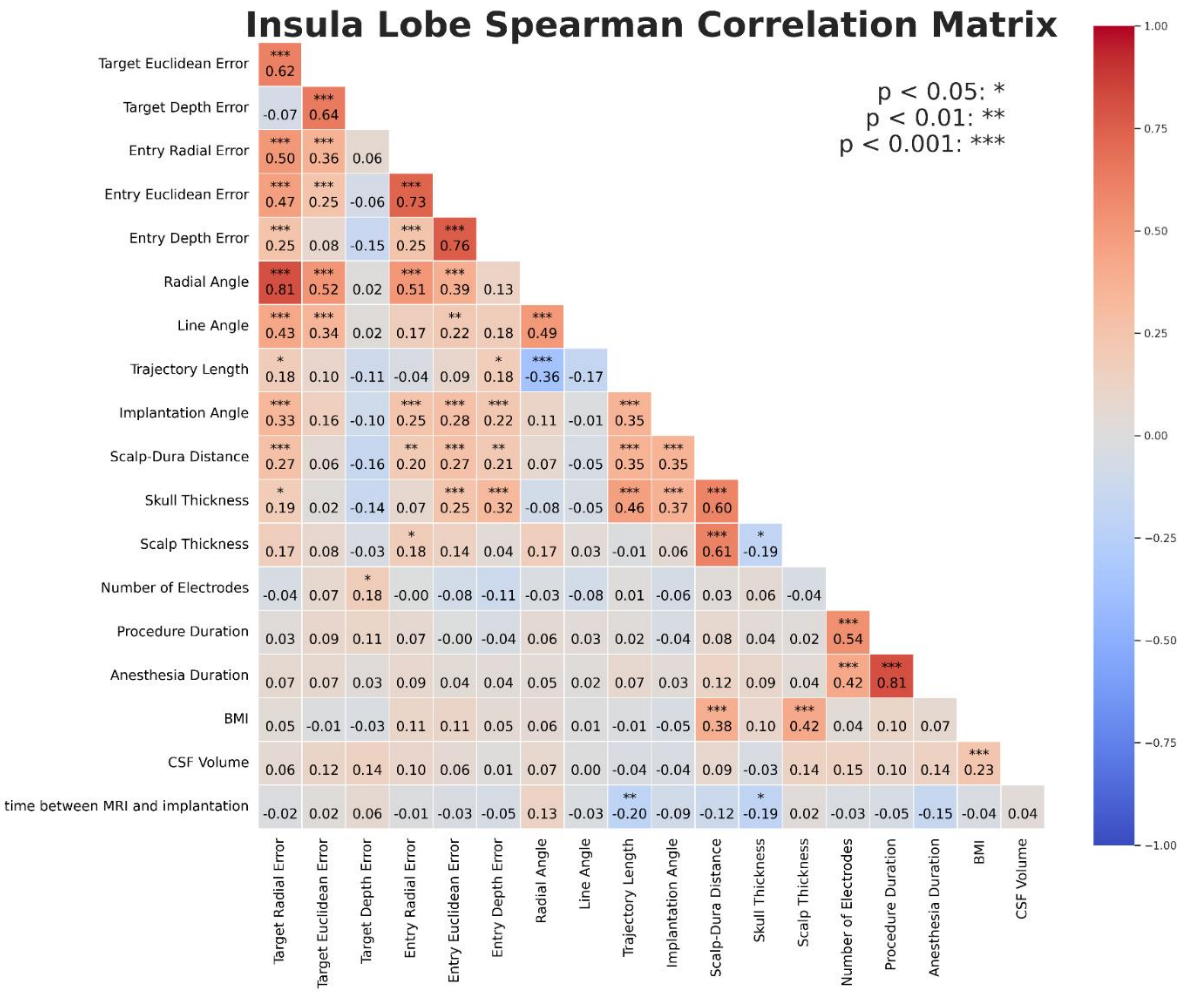
Insula Lobe Spearman Correlation Matrix
p < 0.05: *
p < 0.01: **
p < 0.001: ***
Target Euclidean Error 0.62***
Target Depth Error -0.07 0.64***
Entry Radial Error 0.50*** 0.36*** 0.06
Entry Euclidean Error 0.47*** 0.25*** -0.06 0.73***
Entry Depth Error 0.25*** 0.08 -0.15 0.25*** 0.76***
Radial Angle 0.81*** 0.52*** 0.02 0.51*** 0.39*** 0.13
Line Angle 0.43*** 0.34*** 0.02 0.17 0.22** 0.18 0.49***
Trajectory Length 0.18* 0.10 -0.11 -0.04 0.09 0.18* -0.36*** -0.17
Implantation Angle 0.33*** 0.16 -0.10 0.25*** 0.28*** 0.22*** 0.11 -0.01 0.35***
Scalp-Dura Distance 0.27*** 0.06 -0.16 0.20** 0.27*** 0.21** 0.07 -0.05 0.35*** 0.35***
Skull Thickness 0.19* 0.02 -0.14 0.07 0.25*** 0.32*** -0.08 -0.05 0.46*** 0.37*** 0.60***
Scalp Thickness 0.17 0.08 -0.03 0.18* 0.14 0.04 0.17 0.03 -0.01 0.06 0.61*** -0.19*
Number of Electrodes -0.04 0.07 0.18* -0.00 -0.08 -0.11 -0.03 -0.08 0.01 -0.06 0.03 0.06 -0.04
Procedure Duration 0.03 0.09 0.11 0.07 -0.00 -0.04 0.06 0.03 0.02 -0.04 0.08 0.04 0.02 0.54***
Anesthesia Duration 0.07 0.07 0.03 0.09 0.04 0.04 0.05 0.02 0.07 0.03 0.12 0.09 0.04 0.42*** 0.81***
BMI 0.05 -0.01 -0.03 0.11 0.11 0.05 0.06 0.01 -0.01 -0.05 0.38*** 0.10 0.42*** 0.04 0.10 0.07
CSF Volume 0.06 0.12 0.14 0.10 0.06 0.01 0.07 0.00 -0.04 -0.04 0.09 -0.03 0.14 0.15 0.10 0.14 0.23***
time between MRI and implantation -0.02 0.02 0.06 -0.01 -0.03 -0.05 0.13 -0.03 -0.20** -0.09 -0.12 -0.19* 0.02 -0.03 -0.05 -0.15 -0.04 0.04
Target Radial Error
Target Euclidean Error
Target Depth Error
Entry Radial Error
Entry Euclidean Error
Entry Depth Error
Radial Angle
Line Angle
Trajectory Length
Implantation Angle
Scalp-Dura Distance
Skull Thickness
Scalp Thickness
Number of Electrodes
Procedure Duration
Anesthesia Duration
BMI
CSF Volume
1.00
0.75
0.50
0.25
0.00
−0.25
−0.50
−0.75
−1.00

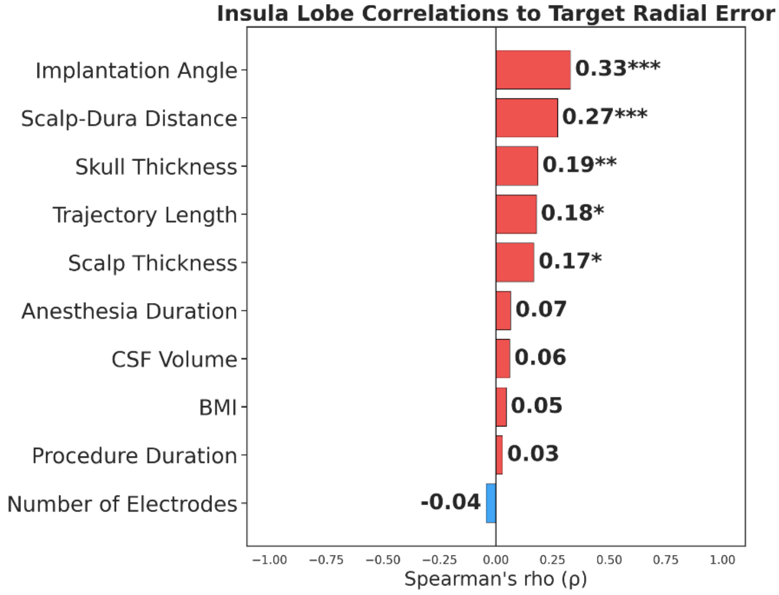
Insula Lobe Correlations to Target Radial Error
Implantation Angle 0.33***
Scalp-Dura Distance 0.27***
Skull Thickness 0.19**
Trajectory Length 0.18*
Scalp Thickness 0.17*
Anesthesia Duration 0.07
CSF Volume 0.06
BMI 0.05
Procedure Duration 0.03
Number of Electrodes -0.04
−1.00 −0.75 −0.50 −0.25 0.00 0.25 0.50 0.75 1.00
Spearman's rho (ρ)

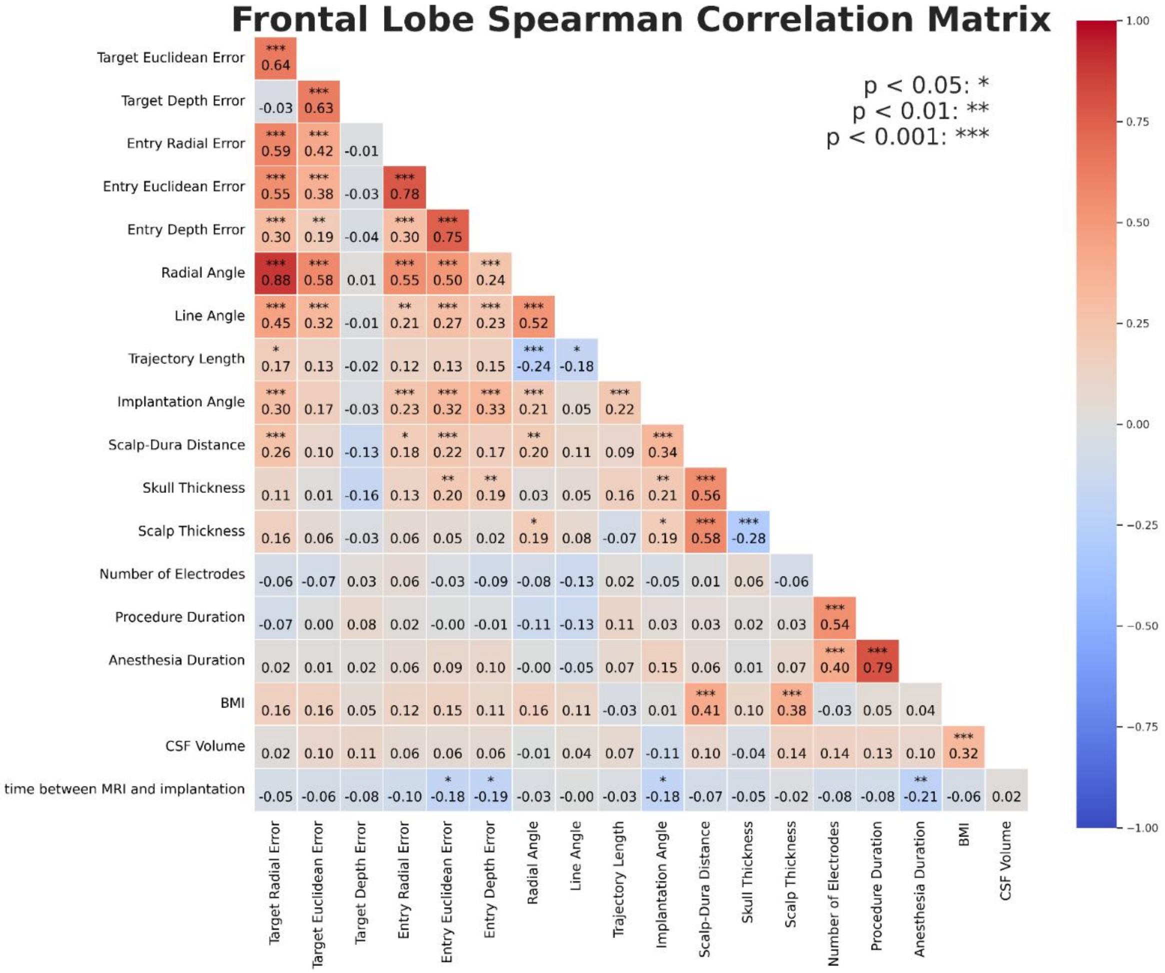
Frontal Lobe Spearman Correlation Matrix
p < 0.05: *
p < 0.01: **
p < 0.001: ***
Target Euclidean Error
Target Depth Error
Entry Radial Error
Entry Euclidean Error
Entry Depth Error
Radial Angle
Line Angle
Trajectory Length
Implantation Angle
Scalp-Dura Distance
Skull Thickness
Scalp Thickness
Number of Electrodes
Procedure Duration
Anesthesia Duration
BMI
CSF Volume
time between MRI and implantation
Target Radial Error
1.00
0.75
0.50
0.25
0.00
−0.25
−0.50
−0.75
−1.00

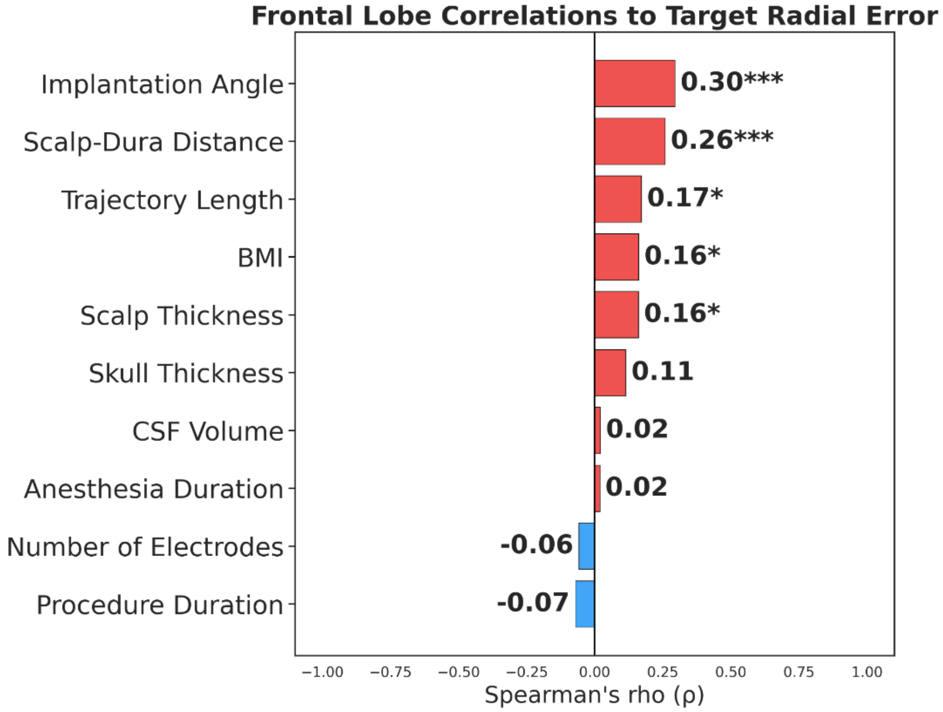
Frontal Lobe Correlations to Target Radial Error
Implantation Angle
Scalp-Dura Distance
Trajectory Length
BMI
Scalp Thickness
Skull Thickness
CSF Volume
Anesthesia Duration
Number of Electrodes
Procedure Duration
0.30***
0.26***
0.17*
0.16*
0.16*
0.11
0.02
0.02
-0.06
-0.07
−1.00 −0.75 −0.50 −0.25 0.00 0.25 0.50 0.75 1.00
Spearman's rho (ρ)

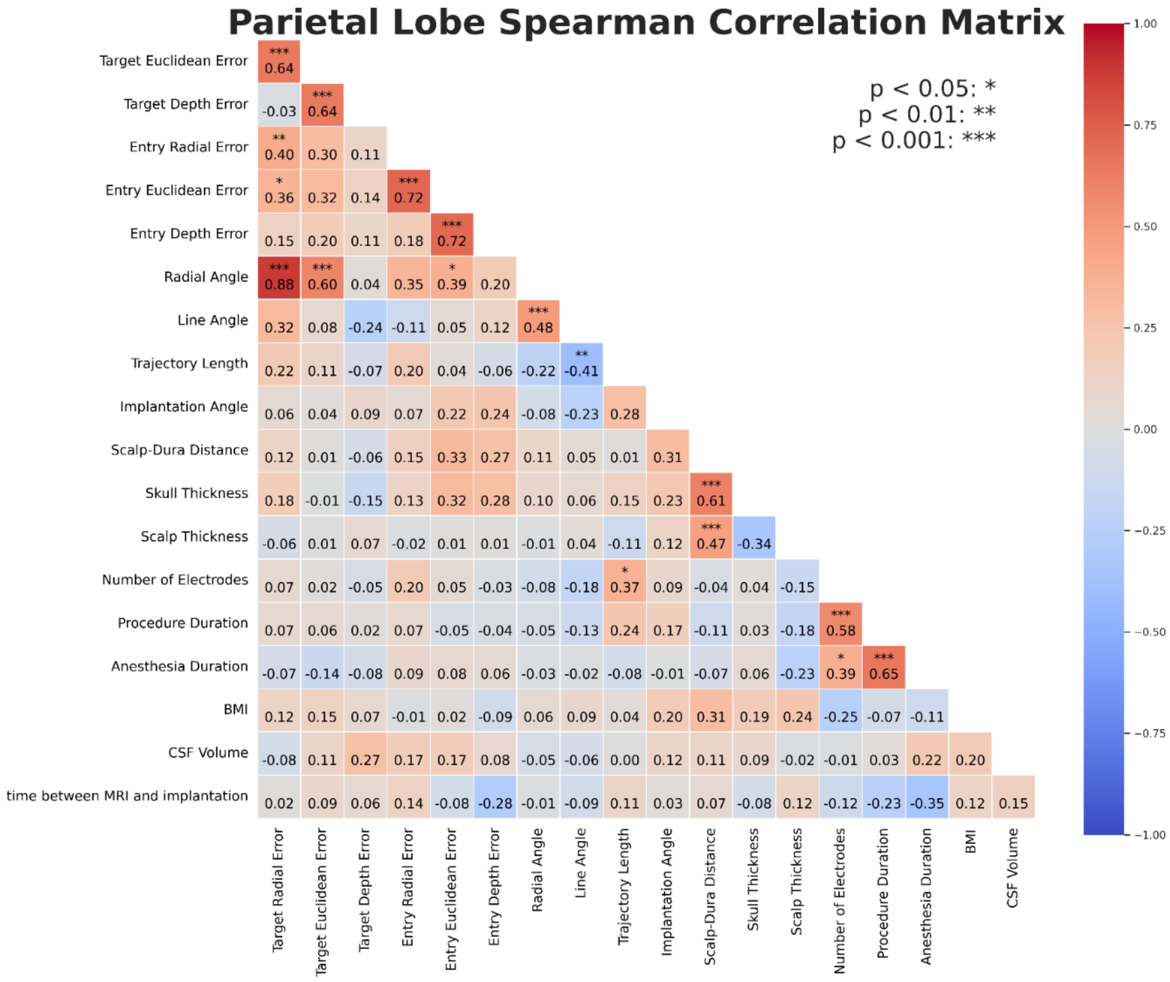
Parietal Lobe Spearman Correlation Matrix
p < 0.05: *
p < 0.01: **
p < 0.001: ***
Target Euclidean Error *** 0.64
Target Depth Error -0.03 *** 0.64
Entry Radial Error ** 0.40 0.30 0.11
Entry Euclidean Error * 0.36 0.32 0.14 *** 0.72
Entry Depth Error 0.15 0.20 0.11 0.18 *** 0.72
Radial Angle *** 0.88 *** 0.60 0.04 0.35 * 0.39 0.20
Line Angle 0.32 0.08 -0.24 -0.11 0.05 0.12 *** 0.48
Trajectory Length 0.22 0.11 -0.07 0.20 0.04 -0.06 -0.22 ** -0.41
Implantation Angle 0.06 0.04 0.09 0.07 0.22 0.24 -0.08 -0.23 0.28
Scalp-Dura Distance 0.12 0.01 -0.06 0.15 0.33 0.27 0.11 0.05 0.01 0.31
Skull Thickness 0.18 -0.01 -0.15 0.13 0.32 0.28 0.10 0.06 0.15 0.23 *** 0.61
Scalp Thickness -0.06 0.01 0.07 -0.02 0.01 0.01 -0.01 0.04 -0.11 0.12 *** 0.47 -0.34
Number of Electrodes 0.07 0.02 -0.05 0.20 0.05 -0.03 -0.08 -0.18 * 0.37 0.09 -0.04 0.04 -0.15
Procedure Duration 0.07 0.06 0.02 0.07 -0.05 -0.04 -0.05 -0.13 0.24 0.17 -0.11 0.03 -0.18 *** 0.58
Anesthesia Duration -0.07 -0.14 -0.08 0.09 0.08 0.06 -0.03 -0.02 -0.08 -0.01 -0.07 0.06 -0.23 * 0.39 *** 0.65
BMI 0.12 0.15 0.07 -0.01 0.02 -0.09 0.06 0.09 0.04 0.20 0.31 0.19 0.24 -0.25 -0.07 -0.11
CSF Volume -0.08 0.11 0.27 0.17 0.17 0.08 -0.05 -0.06 0.00 0.12 0.11 0.09 -0.02 -0.01 0.03 0.22 0.20
time between MRI and implantation 0.02 0.09 0.06 0.14 -0.08 -0.28 -0.01 -0.09 0.11 0.03 0.07 -0.08 0.12 -0.12 -0.23 -0.35 0.12 0.15
Target Radial Error
Target Euclidean Error
Target Depth Error
Entry Radial Error
Entry Euclidean Error
Entry Depth Error
Radial Angle
Line Angle
Trajectory Length
Implantation Angle
Scalp-Dura Distance
Skull Thickness
Scalp Thickness
Number of Electrodes
Procedure Duration
Anesthesia Duration
BMI
CSF Volume
1.00
0.75
0.50
0.25
0.00
−0.25
−0.50
−0.75
−1.00

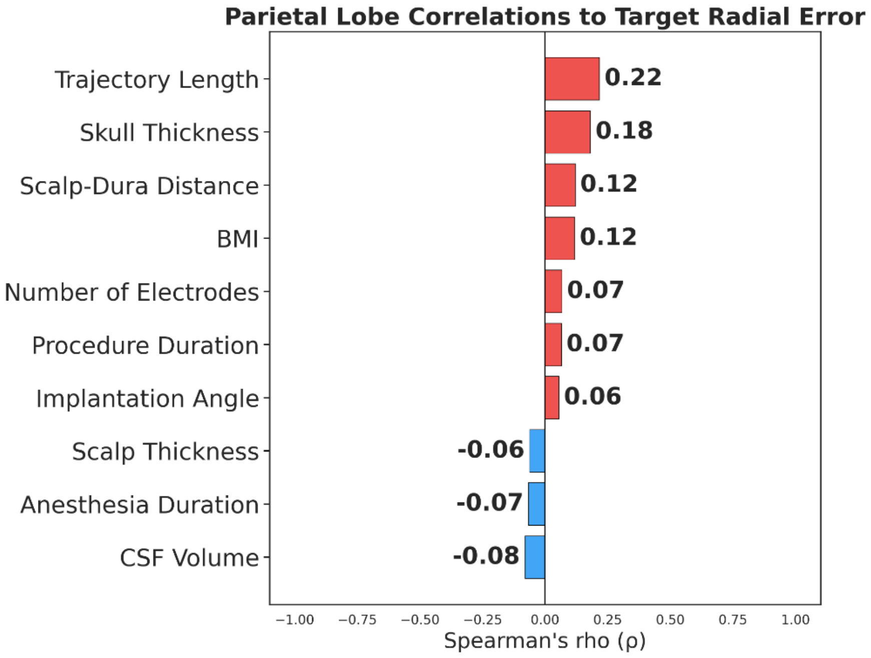
Parietal Lobe Correlations to Target Radial Error
Trajectory Length 0.22
Skull Thickness 0.18
Scalp-Dura Distance 0.12
BMI 0.12
Number of Electrodes 0.07
Procedure Duration 0.07
Implantation Angle 0.06
Scalp Thickness -0.06
Anesthesia Duration -0.07
CSF Volume -0.08
−1.00 −0.75 −0.50 −0.25 0.00 0.25 0.50 0.75 1.00
Spearman's rho (ρ)

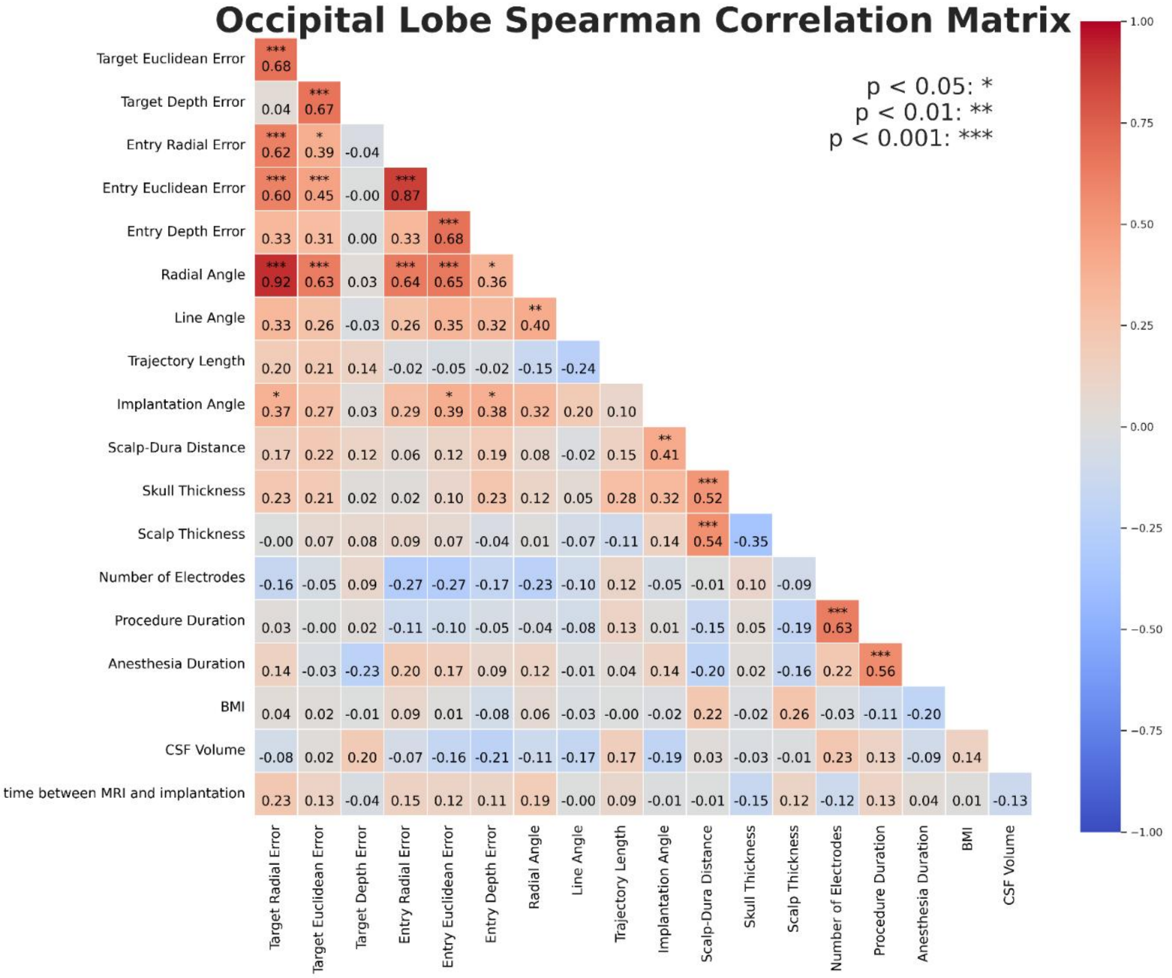

Occipital Lobe Spearman Correlation Matrix
p < 0.05: *
p < 0.01: **
p < 0.001: ***
Target Euclidean Error: *** 0.68
Target Depth Error: 0.04, *** 0.67
Entry Radial Error: *** 0.62, * 0.39, -0.04
Entry Euclidean Error: *** 0.60, *** 0.45, -0.00, *** 0.87
Entry Depth Error: 0.33, 0.31, 0.00, 0.33, *** 0.68
Radial Angle: *** 0.92, *** 0.63, 0.03, *** 0.64, *** 0.65, * 0.36
Line Angle: 0.33, 0.26, -0.03, 0.26, 0.35, 0.32, ** 0.40
Trajectory Length: 0.20, 0.21, 0.14, -0.02, -0.05, -0.02, -0.15, -0.24
Implantation Angle: * 0.37, 0.27, 0.03, 0.29, * 0.39, * 0.38, 0.32, 0.20, 0.10
Scalp-Dura Distance: 0.17, 0.22, 0.12, 0.06, 0.12, 0.19, 0.08, -0.02, 0.15, ** 0.41
Skull Thickness: 0.23, 0.21, 0.02, 0.02, 0.10, 0.23, 0.12, 0.05, 0.28, 0.32, *** 0.52
Scalp Thickness: -0.00, 0.07, 0.08, 0.09, 0.07, -0.04, 0.01, -0.07, -0.11, 0.14, *** 0.54, -0.35
Number of Electrodes: -0.16, -0.05, 0.09, -0.27, -0.27, -0.17, -0.23, -0.10, 0.12, -0.05, -0.01, 0.10, -0.09
Procedure Duration: 0.03, -0.00, 0.02, -0.11, -0.10, -0.05, -0.04, -0.08, 0.13, 0.01, -0.15, 0.05, -0.19, *** 0.63
Anesthesia Duration: 0.14, -0.03, -0.23, 0.20, 0.17, 0.09, 0.12, -0.01, 0.04, 0.14, -0.20, 0.02, -0.16, 0.22, *** 0.56
BMI: 0.04, 0.02, -0.01, 0.09, 0.01, -0.08, 0.06, -0.03, -0.00, -0.02, 0.22, -0.02, 0.26, -0.03, -0.11, -0.20
CSF Volume: -0.08, 0.02, 0.20, -0.07, -0.16, -0.21, -0.11, -0.17, 0.17, -0.19, 0.03, -0.03, -0.01, 0.23, 0.13, -0.09, 0.14
time between MRI and implantation: 0.23, 0.13, -0.04, 0.15, 0.12, 0.11, 0.19, -0.00, 0.09, -0.01, -0.01, -0.15, 0.12, -0.12, 0.13, 0.04, 0.01, -0.13
Target Radial Error
Target Euclidean Error
Target Depth Error
Entry Radial Error
Entry Euclidean Error
Entry Depth Error
Radial Angle
Line Angle
Trajectory Length
Implantation Angle
Scalp-Dura Distance
Skull Thickness
Scalp Thickness
Number of Electrodes
Procedure Duration
Anesthesia Duration
BMI
CSF Volume
1.00
0.75
0.50
0.25
0.00
−0.25
−0.50
−0.75
−1.00

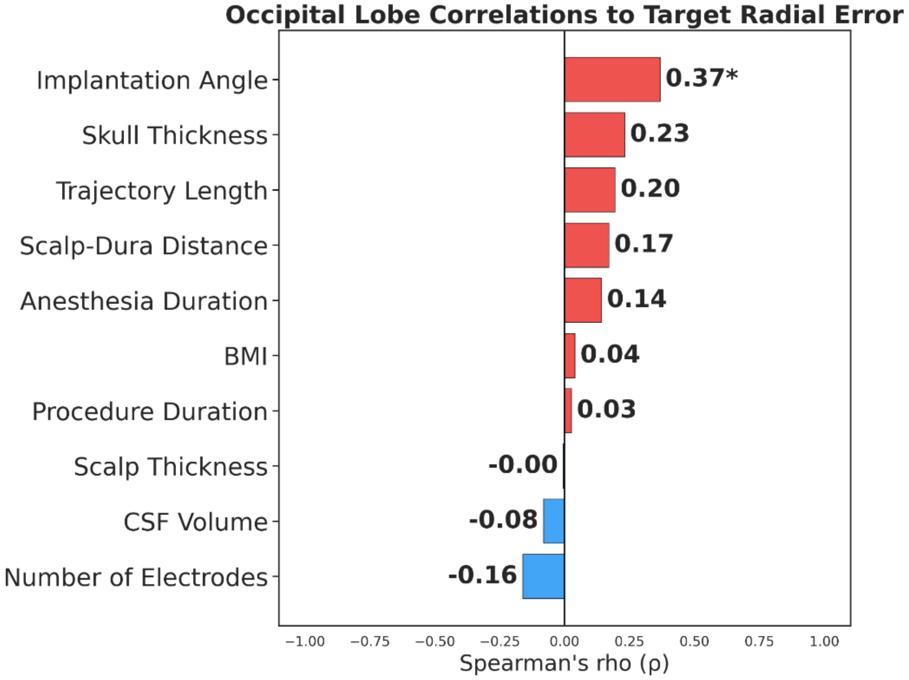

Occipital Lobe Correlations to Target Radial Error
Implantation Angle 0.37*
Skull Thickness 0.23
Trajectory Length 0.20
Scalp-Dura Distance 0.17
Anesthesia Duration 0.14
BMI 0.04
Procedure Duration 0.03
Scalp Thickness -0.00
CSF Volume -0.08
Number of Electrodes -0.16
−1.00 −0.75 −0.50 −0.25 0.00 0.25 0.50 0.75 1.00
Spearman's rho (ρ)

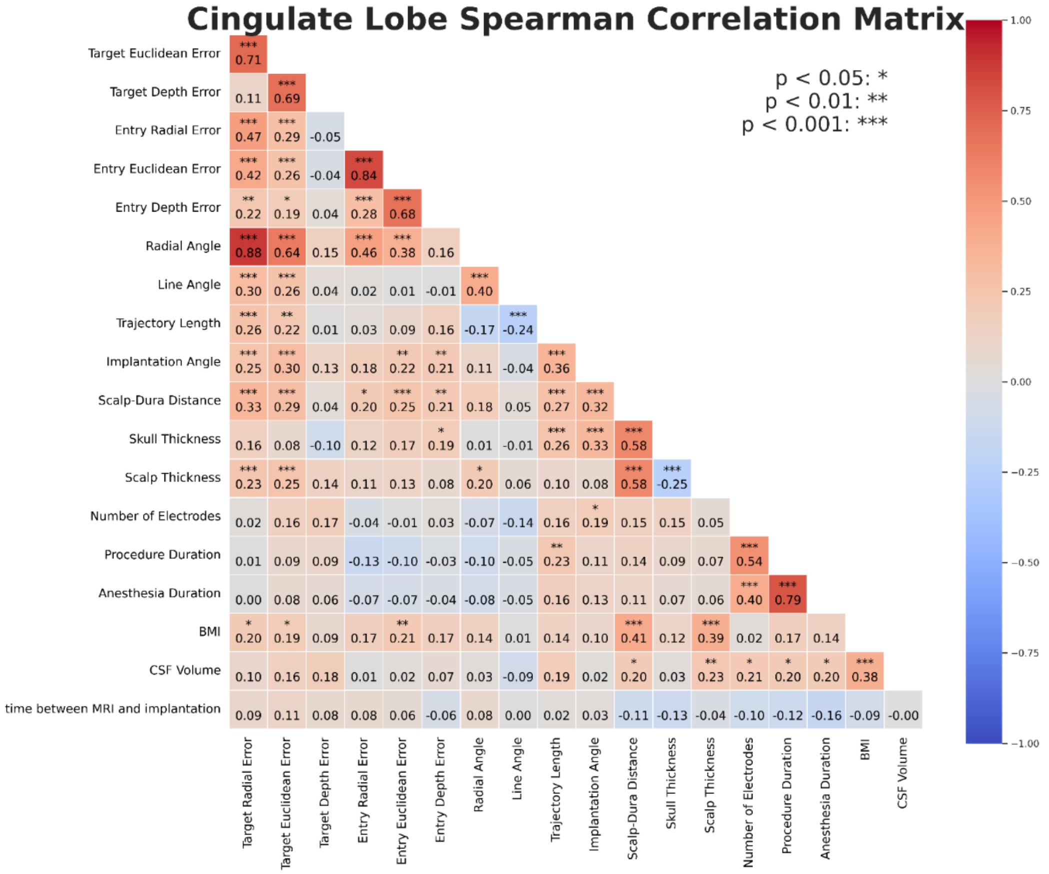

Cingulate Lobe Spearman Correlation Matrix
p < 0.05: *
p < 0.01: **
p < 0.001: ***
Target Euclidean Error
Target Depth Error
Entry Radial Error
Entry Euclidean Error
Entry Depth Error
Radial Angle
Line Angle
Trajectory Length
Implantation Angle
Scalp-Dura Distance
Skull Thickness
Scalp Thickness
Number of Electrodes
Procedure Duration
Anesthesia Duration
BMI
CSF Volume
time between MRI and implantation
Target Radial Error

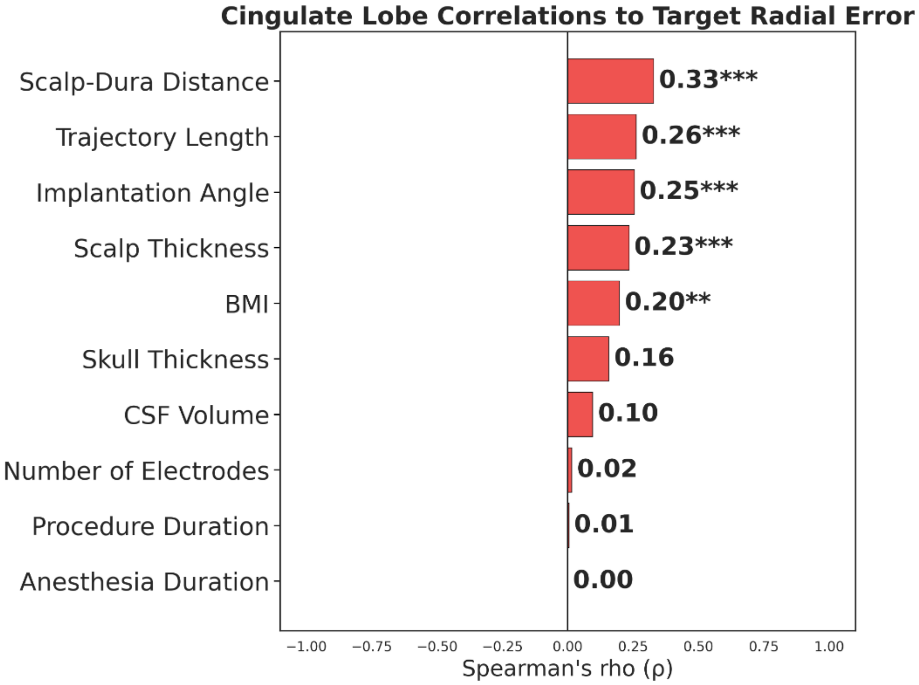

Cingulate Lobe Correlations to Target Radial Error
Scalp-Dura Distance 0.33***
Trajectory Length 0.26***
Implantation Angle 0.25***
Scalp Thickness 0.23***
BMI 0.20**
Skull Thickness 0.16
CSF Volume 0.10
Number of Electrodes 0.02
Procedure Duration 0.01
Anesthesia Duration 0.00
Spearman's rho (ρ)

**Supplementary Figure 4.** Forest plots and ROC Curve for implantation angle and target radial error, considering implanted lobe.

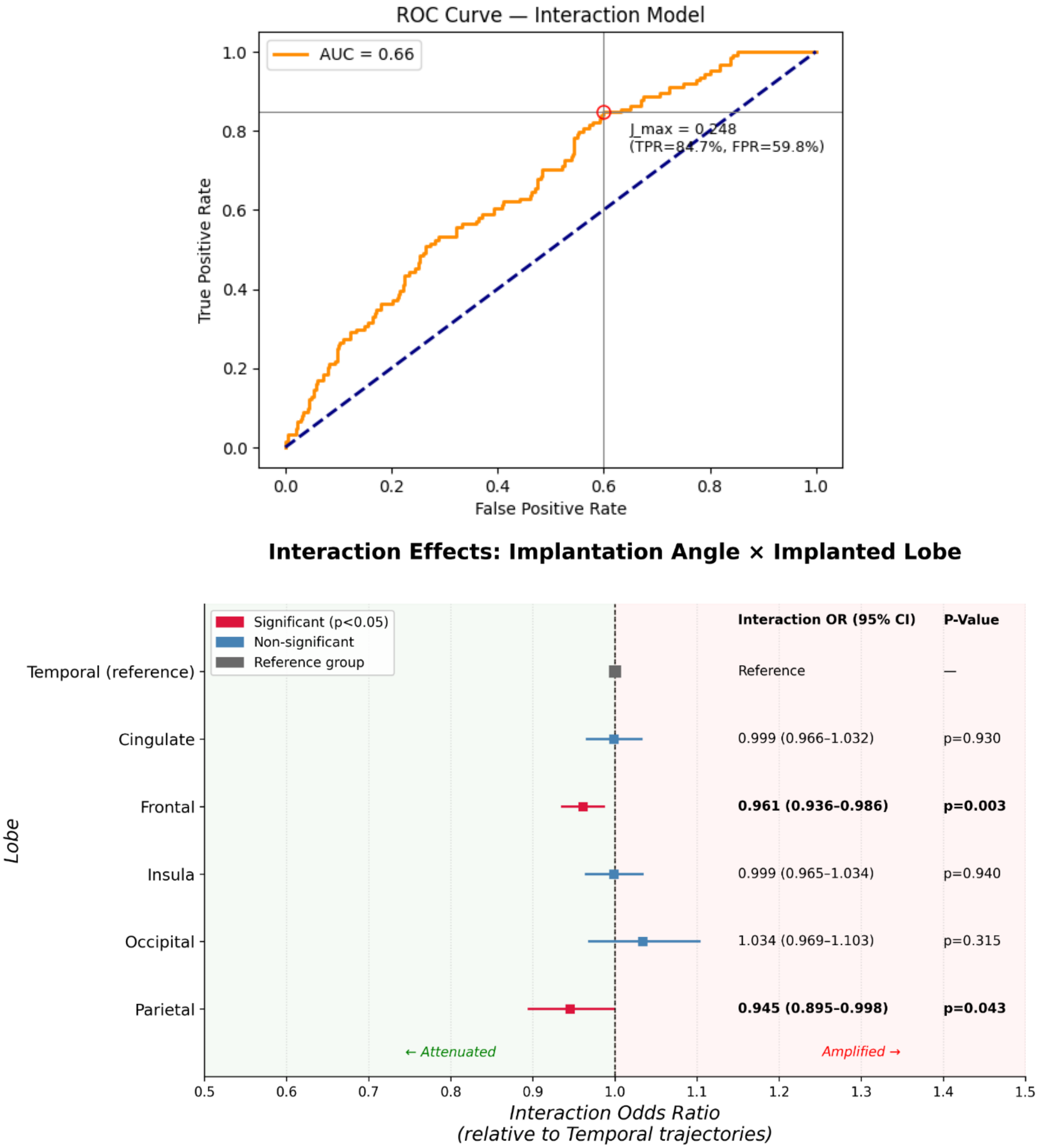

Cross-validated AUC: **0.67 ± 0.01** | AUC Score: **0.66**
Sensitivity: **0.847** | Specificity: **0.402** | PPV: **0.333** | NPV: **0.881** | Optimal threshold: **0.391** | Youden J: **0.248**
Per-group angle cutpoints (interaction model):
Temporal: 28.09°, Cingulate: 29.18°, Frontal: 33.86°, Insula: 32.24°, Occipital: 27.65°, Parietal: 60.75°

**Supplementary Figure 6.** Spearman correlation for surgeon-labeled targets.

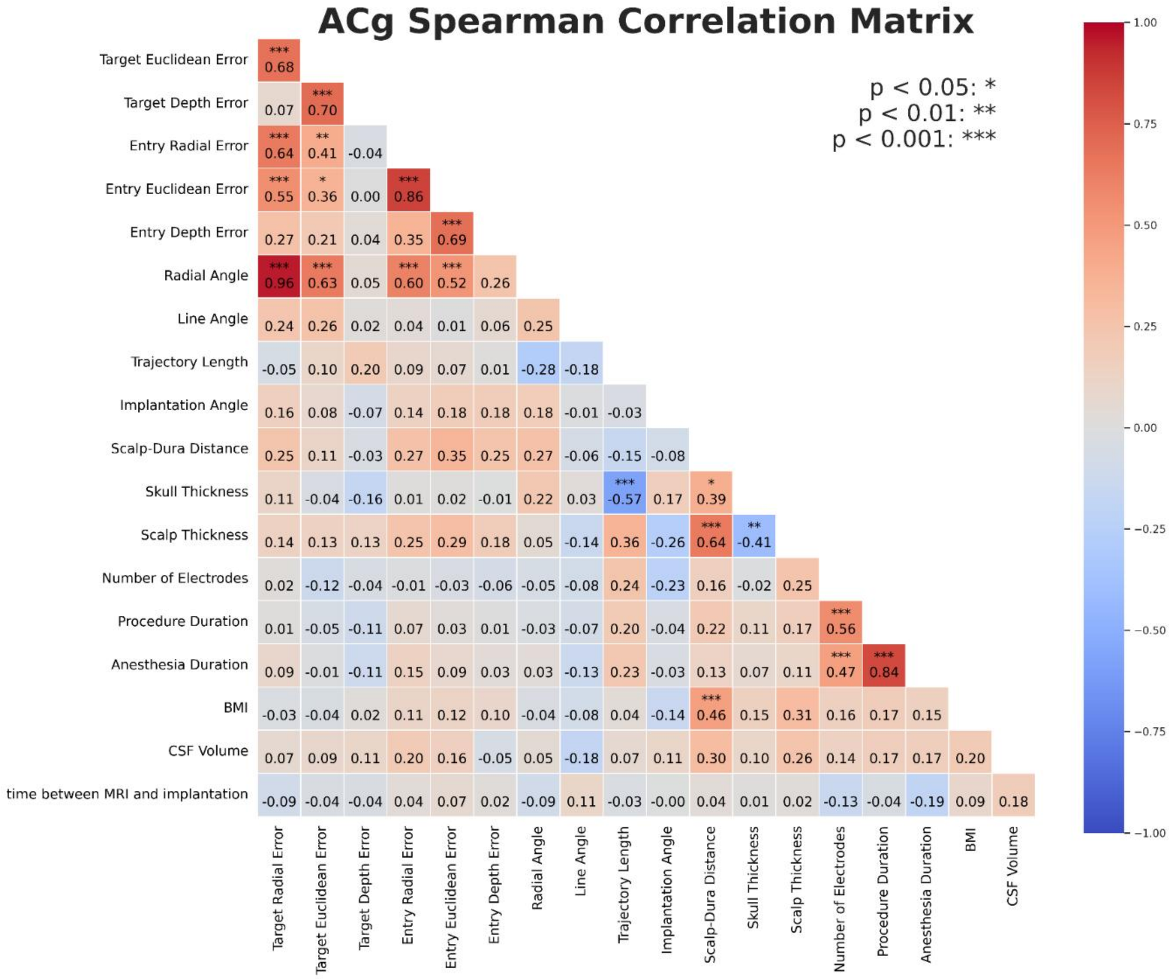

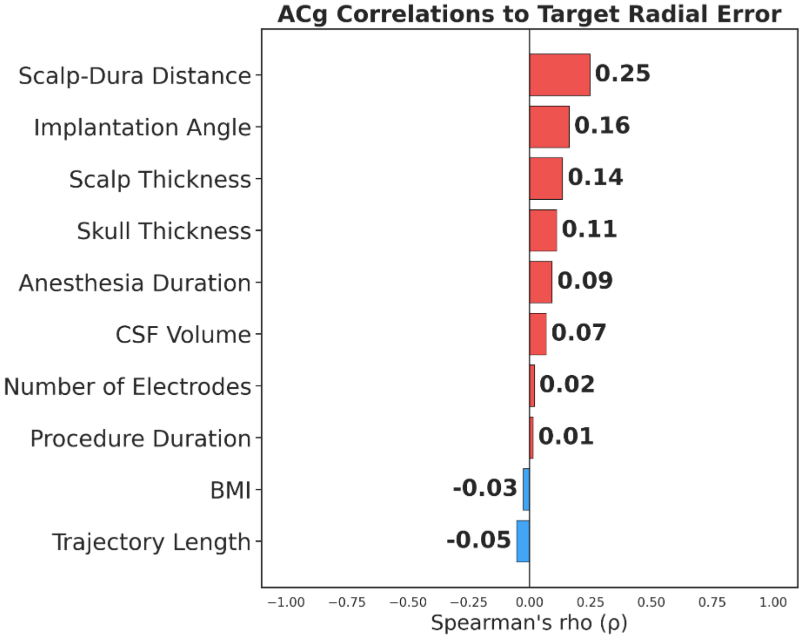

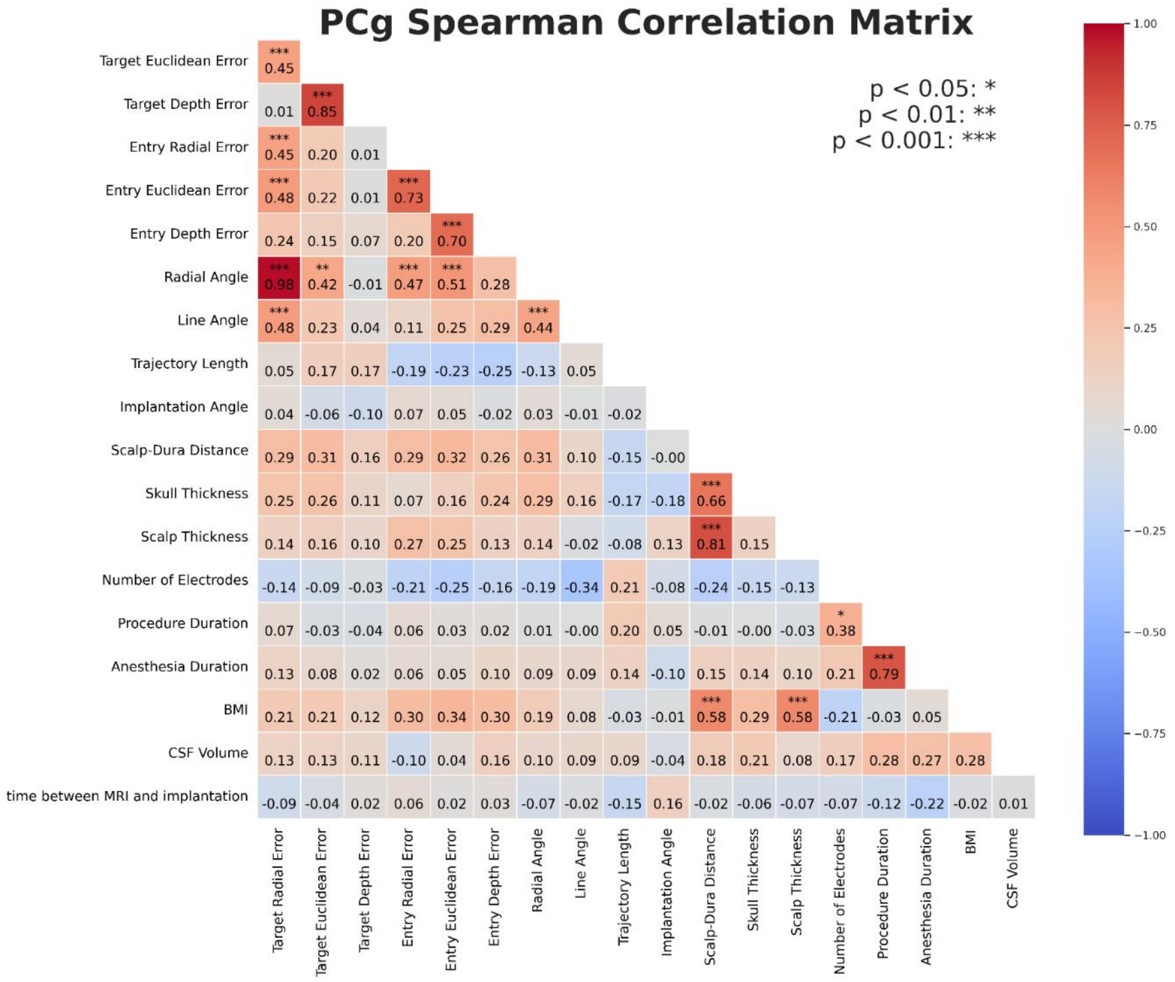
PCg Spearman Correlation Matrix
p < 0.05: *
p < 0.01: **
p < 0.001: ***
Target Euclidean Error *** 0.45
Target Depth Error 0.01 *** 0.85
Entry Radial Error *** 0.45 0.20 0.01
Entry Euclidean Error *** 0.48 0.22 0.01 *** 0.73
Entry Depth Error 0.24 0.15 0.07 0.20 *** 0.70
Radial Angle *** 0.98 ** 0.42 -0.01 *** 0.47 *** 0.51 0.28
Line Angle *** 0.48 0.23 0.04 0.11 0.25 0.29 *** 0.44
Trajectory Length 0.05 0.17 0.17 -0.19 -0.23 -0.25 -0.13 0.05
Implantation Angle 0.04 -0.06 -0.10 0.07 0.05 -0.02 0.03 -0.01 -0.02
Scalp-Dura Distance 0.29 0.31 0.16 0.29 0.32 0.26 0.31 0.10 -0.15 -0.00
Skull Thickness 0.25 0.26 0.11 0.07 0.16 0.24 0.29 0.16 -0.17 -0.18 *** 0.66
Scalp Thickness 0.14 0.16 0.10 0.27 0.25 0.13 0.14 -0.02 -0.08 0.13 *** 0.81 0.15
Number of Electrodes -0.14 -0.09 -0.03 -0.21 -0.25 -0.16 -0.19 -0.34 0.21 -0.08 -0.24 -0.15 -0.13
Procedure Duration 0.07 -0.03 -0.04 0.06 0.03 0.02 0.01 -0.00 0.20 0.05 -0.01 -0.00 -0.03 * 0.38
Anesthesia Duration 0.13 0.08 0.02 0.06 0.05 0.10 0.09 0.09 0.14 -0.10 0.15 0.14 0.10 0.21 *** 0.79
BMI 0.21 0.21 0.12 0.30 0.34 0.30 0.19 0.08 -0.03 -0.01 *** 0.58 0.29 *** 0.58 -0.21 -0.03 0.05
CSF Volume 0.13 0.13 0.11 -0.10 0.04 0.16 0.10 0.09 0.09 -0.04 0.18 0.21 0.08 0.17 0.28 0.27 0.28
time between MRI and implantation -0.09 -0.04 0.02 0.06 0.02 0.03 -0.07 -0.02 -0.15 0.16 -0.02 -0.06 -0.07 -0.07 -0.12 -0.22 -0.02 0.01
Target Radial Error
Target Euclidean Error
Target Depth Error
Entry Radial Error
Entry Euclidean Error
Entry Depth Error
Radial Angle
Line Angle
Trajectory Length
Implantation Angle
Scalp-Dura Distance
Skull Thickness
Scalp Thickness
Number of Electrodes
Procedure Duration
Anesthesia Duration
BMI
CSF Volume
1.00
0.75
0.50
0.25
0.00
−0.25
−0.50
−0.75
−1.00

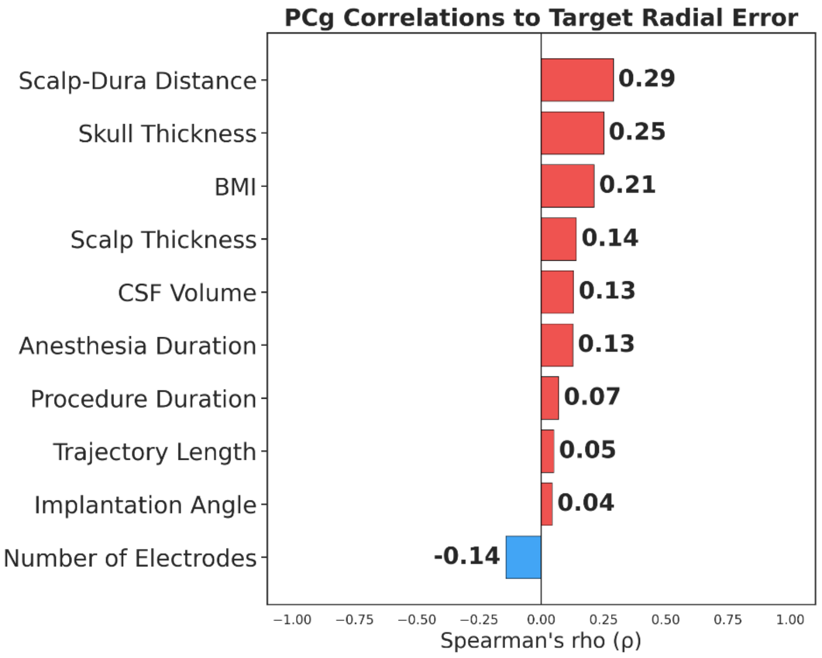
PCg Correlations to Target Radial Error
Scalp-Dura Distance 0.29
Skull Thickness 0.25
BMI 0.21
Scalp Thickness 0.14
CSF Volume 0.13
Anesthesia Duration 0.13
Procedure Duration 0.07
Trajectory Length 0.05
Implantation Angle 0.04
Number of Electrodes -0.14
−1.00 −0.75 −0.50 −0.25 0.00 0.25 0.50 0.75 1.00
Spearman's rho (ρ)

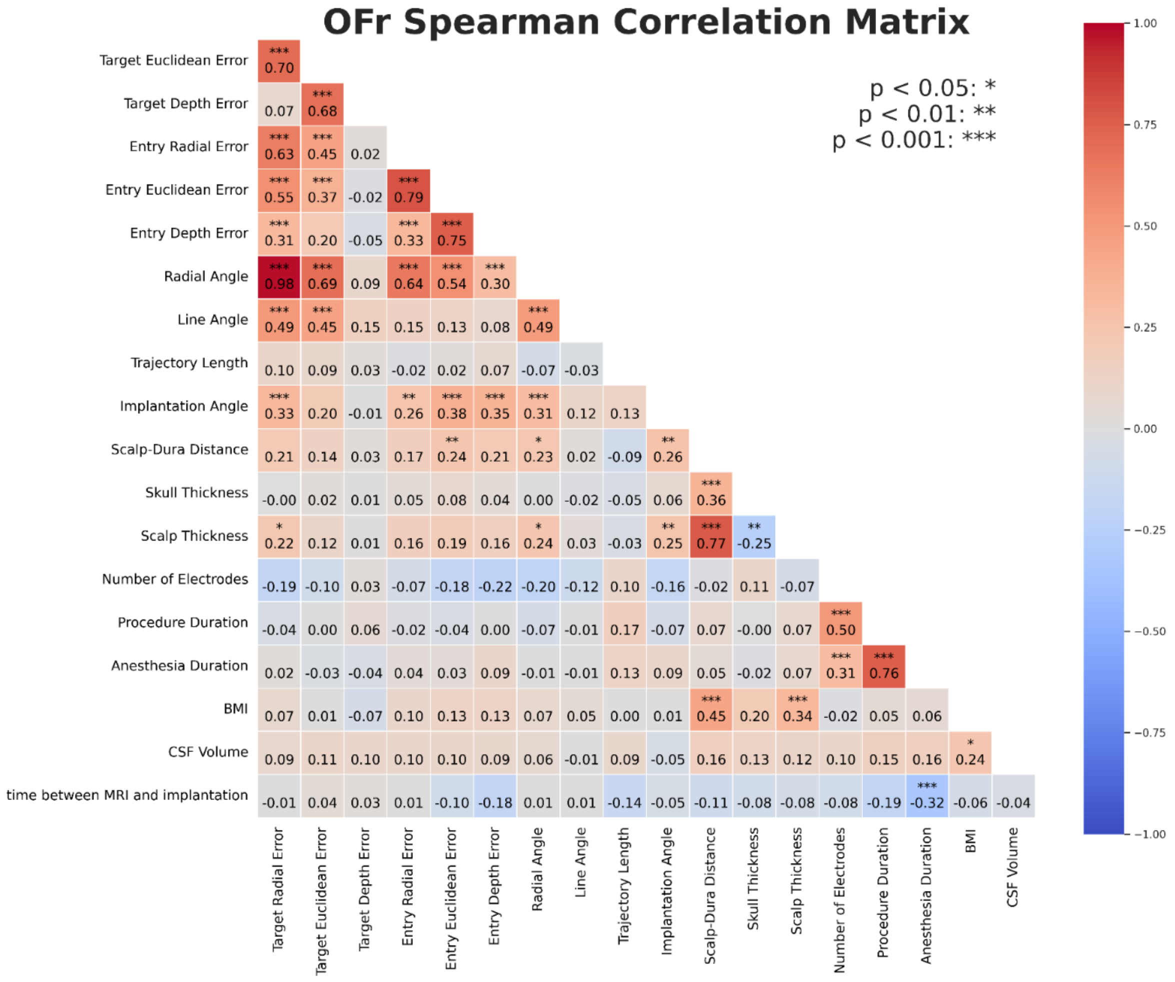
OFr Spearman Correlation Matrix
p < 0.05: *
p < 0.01: **
p < 0.001: ***
Target Euclidean Error 0.70
Target Depth Error 0.07 0.68
Entry Radial Error 0.63 0.45 0.02
Entry Euclidean Error 0.55 0.37 -0.02 0.79
Entry Depth Error 0.31 0.20 -0.05 0.33 0.75
Radial Angle 0.98 0.69 0.09 0.64 0.54 0.30
Line Angle 0.49 0.45 0.15 0.15 0.13 0.08 0.49
Trajectory Length 0.10 0.09 0.03 -0.02 0.02 0.07 -0.07 -0.03
Implantation Angle 0.33 0.20 -0.01 0.26 0.38 0.35 0.31 0.12 0.13
Scalp-Dura Distance 0.21 0.14 0.03 0.17 0.24 0.21 0.23 0.02 -0.09 0.26
Skull Thickness -0.00 0.02 0.01 0.05 0.08 0.04 0.00 -0.02 -0.05 0.06 0.36
Scalp Thickness 0.22 0.12 0.01 0.16 0.19 0.16 0.24 0.03 -0.03 0.25 0.77 -0.25
Number of Electrodes -0.19 -0.10 0.03 -0.07 -0.18 -0.22 -0.20 -0.12 0.10 -0.16 -0.02 0.11 -0.07
Procedure Duration -0.04 0.00 0.06 -0.02 -0.04 0.00 -0.07 -0.01 0.17 -0.07 0.07 -0.00 0.07 0.50
Anesthesia Duration 0.02 -0.03 -0.04 0.04 0.03 0.09 -0.01 -0.01 0.13 0.09 0.05 -0.02 0.07 0.31 0.76
BMI 0.07 0.01 -0.07 0.10 0.13 0.13 0.07 0.05 0.00 0.01 0.45 0.20 0.34 -0.02 0.05 0.06
CSF Volume 0.09 0.11 0.10 0.10 0.10 0.09 0.06 -0.01 0.09 -0.05 0.16 0.13 0.12 0.10 0.15 0.16 0.24
time between MRI and implantation -0.01 0.04 0.03 0.01 -0.10 -0.18 0.01 0.01 -0.14 -0.05 -0.11 -0.08 -0.08 -0.08 -0.19 -0.32 -0.06 -0.04
Target Radial Error
Target Euclidean Error
Target Depth Error
Entry Radial Error
Entry Euclidean Error
Entry Depth Error
Radial Angle
Line Angle
Trajectory Length
Implantation Angle
Scalp-Dura Distance
Skull Thickness
Scalp Thickness
Number of Electrodes
Procedure Duration
Anesthesia Duration
BMI
CSF Volume
1.00
0.75
0.50
0.25
0.00
−0.25
−0.50
−0.75
−1.00

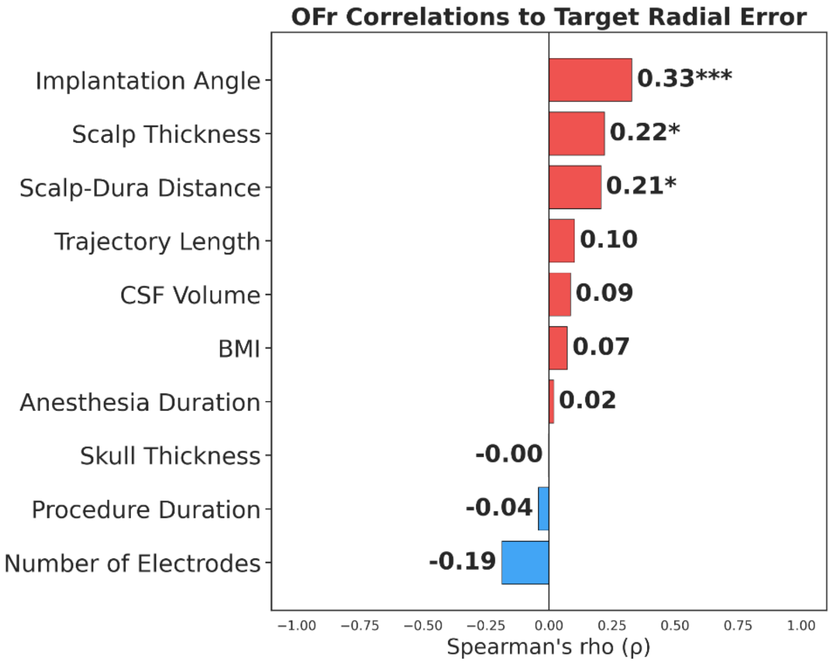
OFr Correlations to Target Radial Error
Implantation Angle 0.33***
Scalp Thickness 0.22*
Scalp-Dura Distance 0.21*
Trajectory Length 0.10
CSF Volume 0.09
BMI 0.07
Anesthesia Duration 0.02
Skull Thickness -0.00
Procedure Duration -0.04
Number of Electrodes -0.19
−1.00 −0.75 −0.50 −0.25 0.00 0.25 0.50 0.75 1.00
Spearman's rho (ρ)

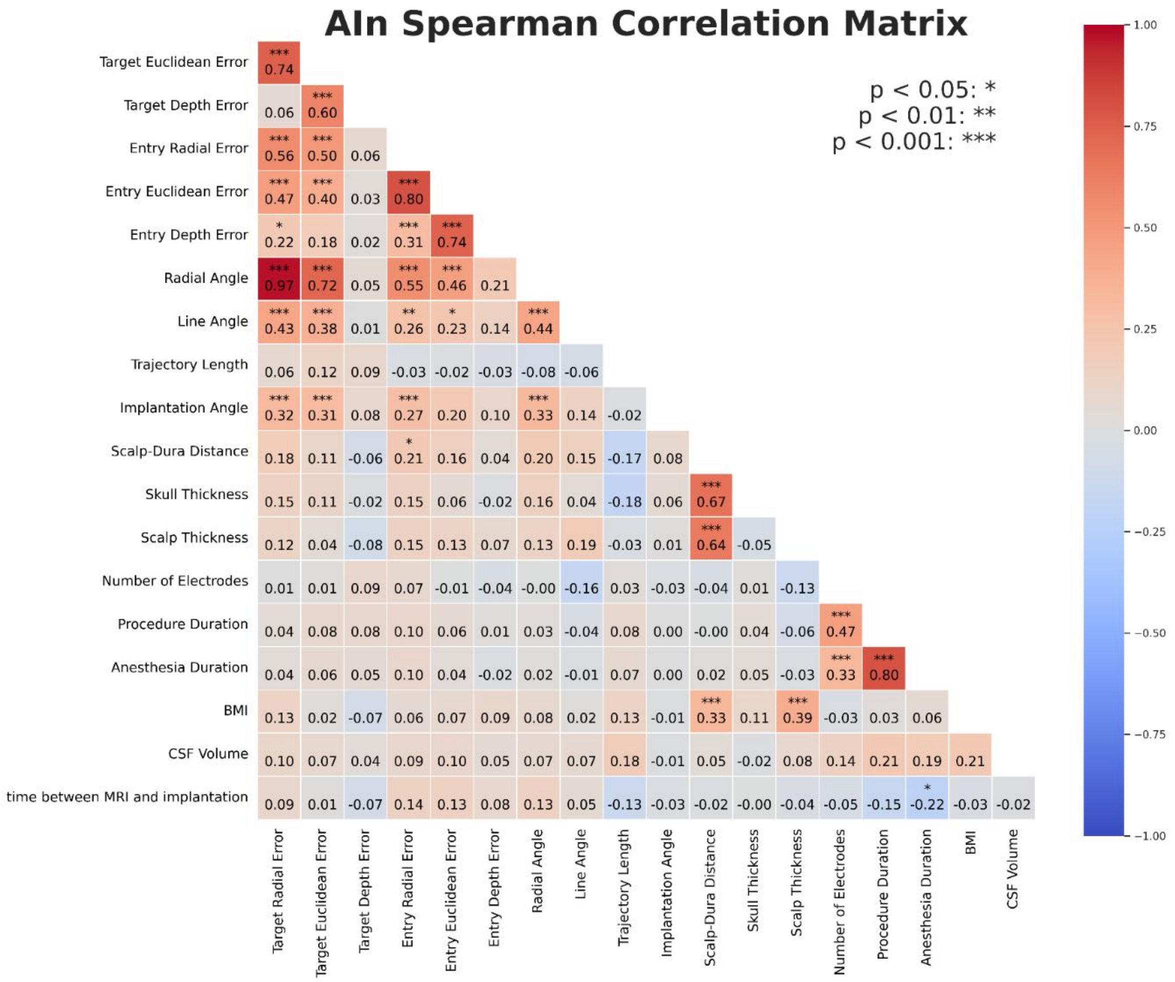
AIn Spearman Correlation Matrix
p < 0.05: *
p < 0.01: **
p < 0.001: ***
Target Euclidean Error 0.74
Target Depth Error 0.06 0.60
Entry Radial Error 0.56 0.50 0.06
Entry Euclidean Error 0.47 0.40 0.03 0.80
Entry Depth Error 0.22 0.18 0.02 0.31 0.74
Radial Angle 0.97 0.72 0.05 0.55 0.46 0.21
Line Angle 0.43 0.38 0.01 0.26 0.23 0.14 0.44
Trajectory Length 0.06 0.12 0.09 -0.03 -0.02 -0.03 -0.08 -0.06
Implantation Angle 0.32 0.31 0.08 0.27 0.20 0.10 0.33 0.14 -0.02
Scalp-Dura Distance 0.18 0.11 -0.06 0.21 0.16 0.04 0.20 0.15 -0.17 0.08
Skull Thickness 0.15 0.11 -0.02 0.15 0.06 -0.02 0.16 0.04 -0.18 0.06 0.67
Scalp Thickness 0.12 0.04 -0.08 0.15 0.13 0.07 0.13 0.19 -0.03 0.01 0.64 -0.05
Number of Electrodes 0.01 0.01 0.09 0.07 -0.01 -0.04 -0.00 -0.16 0.03 -0.03 -0.04 0.01 -0.13
Procedure Duration 0.04 0.08 0.08 0.10 0.06 0.01 0.03 -0.04 0.08 0.00 -0.00 0.04 -0.06 0.47
Anesthesia Duration 0.04 0.06 0.05 0.10 0.04 -0.02 0.02 -0.01 0.07 0.00 0.02 0.05 -0.03 0.33 0.80
BMI 0.13 0.02 -0.07 0.06 0.07 0.09 0.08 0.02 0.13 -0.01 0.33 0.11 0.39 -0.03 0.03 0.06
CSF Volume 0.10 0.07 0.04 0.09 0.10 0.05 0.07 0.07 0.18 -0.01 0.05 -0.02 0.08 0.14 0.21 0.19 0.21
time between MRI and implantation 0.09 0.01 -0.07 0.14 0.13 0.08 0.13 0.05 -0.13 -0.03 -0.02 -0.00 -0.04 -0.05 -0.15 -0.22 -0.03 -0.02
Target Radial Error
Target Euclidean Error
Target Depth Error
Entry Radial Error
Entry Euclidean Error
Entry Depth Error
Radial Angle
Line Angle
Trajectory Length
Implantation Angle
Scalp-Dura Distance
Skull Thickness
Scalp Thickness
Number of Electrodes
Procedure Duration
Anesthesia Duration
BMI
CSF Volume
1.00
0.75
0.50
0.25
0.00
−0.25
−0.50
−0.75
−1.00

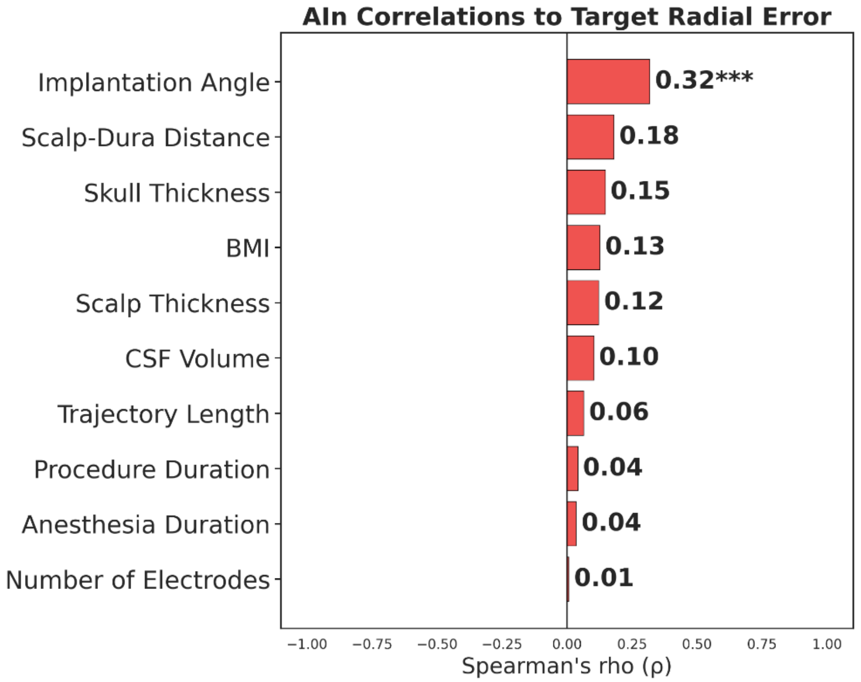
AIn Correlations to Target Radial Error
Implantation Angle 0.32***
Scalp-Dura Distance 0.18
Skull Thickness 0.15
BMI 0.13
Scalp Thickness 0.12
CSF Volume 0.10
Trajectory Length 0.06
Procedure Duration 0.04
Anesthesia Duration 0.04
Number of Electrodes 0.01
−1.00 −0.75 −0.50 −0.25 0.00 0.25 0.50 0.75 1.00
Spearman's rho (ρ)

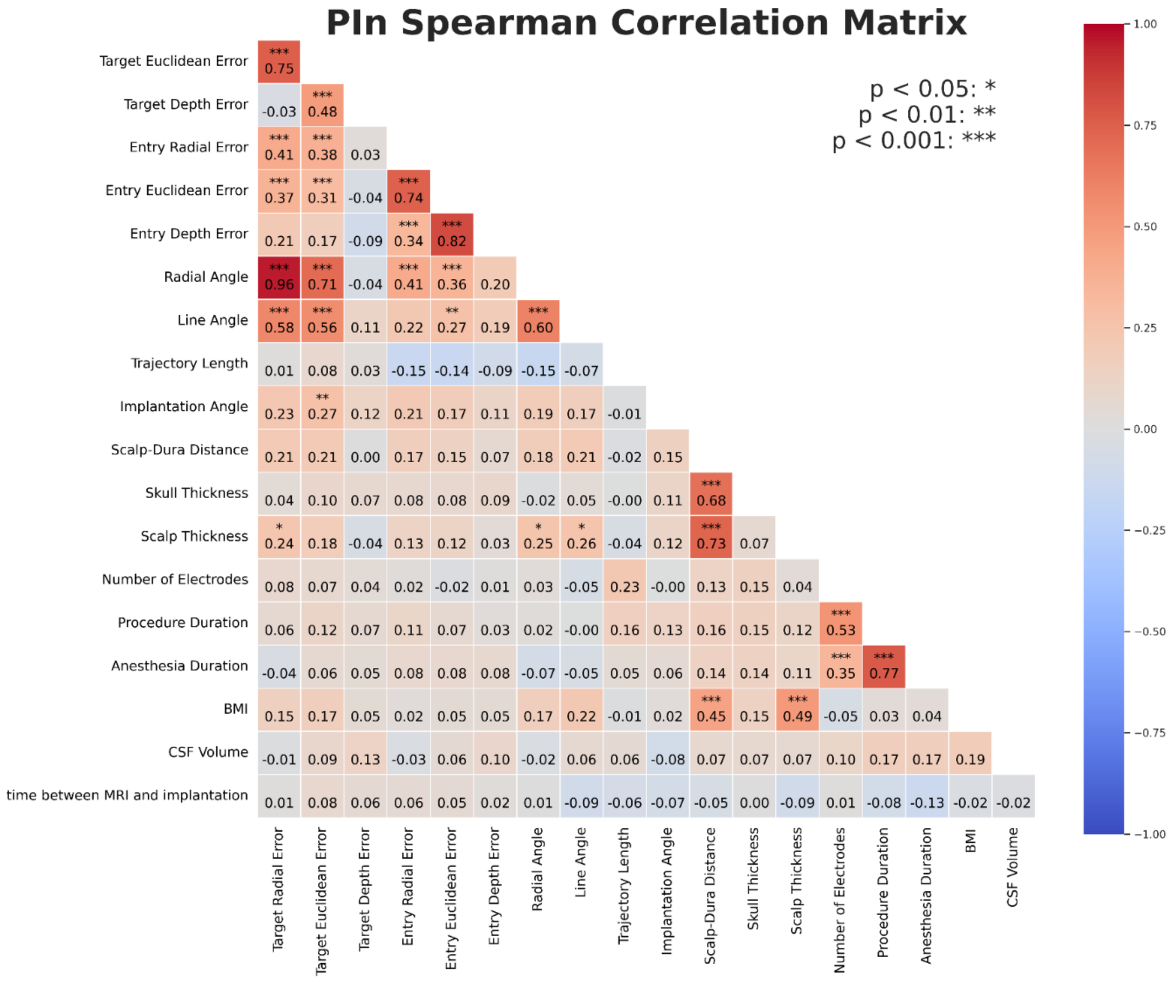
PIn Spearman Correlation Matrix
p < 0.05: *
p < 0.01: **
p < 0.001: ***
Target Euclidean Error: *** 0.75
Target Depth Error: -0.03, *** 0.48
Entry Radial Error: *** 0.41, *** 0.38, 0.03
Entry Euclidean Error: *** 0.37, *** 0.31, -0.04, *** 0.74
Entry Depth Error: 0.21, 0.17, -0.09, *** 0.34, *** 0.82
Radial Angle: *** 0.96, *** 0.71, -0.04, *** 0.41, *** 0.36, 0.20
Line Angle: *** 0.58, *** 0.56, 0.11, 0.22, ** 0.27, 0.19, *** 0.60
Trajectory Length: 0.01, 0.08, 0.03, -0.15, -0.14, -0.09, -0.15, -0.07
Implantation Angle: 0.23, ** 0.27, 0.12, 0.21, 0.17, 0.11, 0.19, 0.17, -0.01
Scalp-Dura Distance: 0.21, 0.21, 0.00, 0.17, 0.15, 0.07, 0.18, 0.21, -0.02, 0.15
Skull Thickness: 0.04, 0.10, 0.07, 0.08, 0.08, 0.09, -0.02, 0.05, -0.00, 0.11, *** 0.68
Scalp Thickness: * 0.24, 0.18, -0.04, 0.13, 0.12, 0.03, * 0.25, * 0.26, -0.04, 0.12, *** 0.73, 0.07
Number of Electrodes: 0.08, 0.07, 0.04, 0.02, -0.02, 0.01, 0.03, -0.05, 0.23, -0.00, 0.13, 0.15, 0.04
Procedure Duration: 0.06, 0.12, 0.07, 0.11, 0.07, 0.03, 0.02, -0.00, 0.16, 0.13, 0.16, 0.15, 0.12, *** 0.53
Anesthesia Duration: -0.04, 0.06, 0.05, 0.08, 0.08, 0.08, -0.07, -0.05, 0.05, 0.06, 0.14, 0.14, 0.11, *** 0.35, *** 0.77
BMI: 0.15, 0.17, 0.05, 0.02, 0.05, 0.05, 0.17, 0.22, -0.01, 0.02, *** 0.45, 0.15, *** 0.49, -0.05, 0.03, 0.04
CSF Volume: -0.01, 0.09, 0.13, -0.03, 0.06, 0.10, -0.02, 0.06, 0.06, -0.08, 0.07, 0.07, 0.07, 0.10, 0.17, 0.17, 0.19
time between MRI and implantation: 0.01, 0.08, 0.06, 0.06, 0.05, 0.02, 0.01, -0.09, -0.06, -0.07, -0.05, 0.00, -0.09, 0.01, -0.08, -0.13, -0.02, -0.02
Target Radial Error
Target Euclidean Error
Target Depth Error
Entry Radial Error
Entry Euclidean Error
Entry Depth Error
Radial Angle
Line Angle
Trajectory Length
Implantation Angle
Scalp-Dura Distance
Skull Thickness
Scalp Thickness
Number of Electrodes
Procedure Duration
Anesthesia Duration
BMI
CSF Volume
1.00
0.75
0.50
0.25
0.00
−0.25
−0.50
−0.75
−1.00

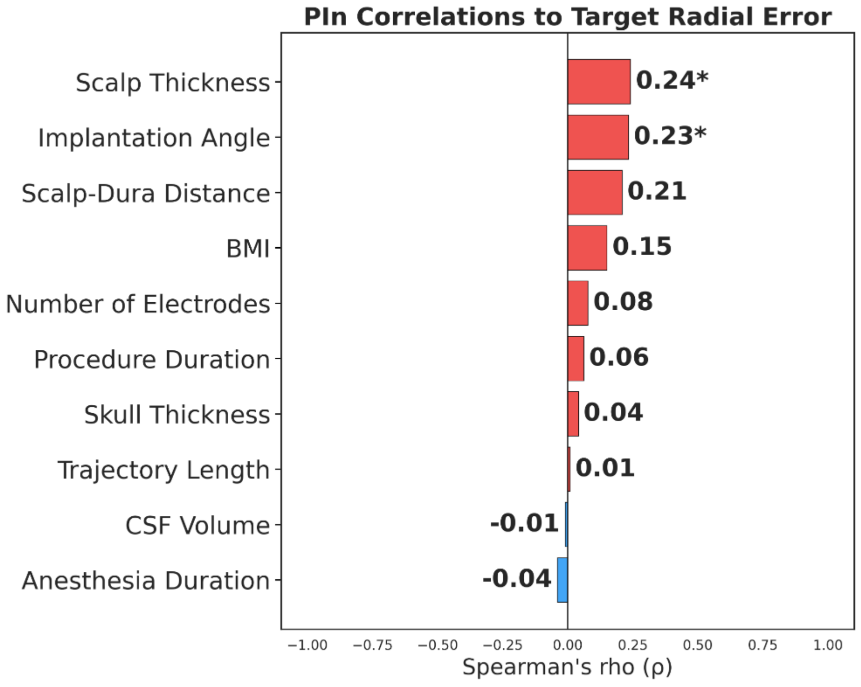
PIn Correlations to Target Radial Error
Scalp Thickness 0.24*
Implantation Angle 0.23*
Scalp-Dura Distance 0.21
BMI 0.15
Number of Electrodes 0.08
Procedure Duration 0.06
Skull Thickness 0.04
Trajectory Length 0.01
CSF Volume -0.01
Anesthesia Duration -0.04
−1.00 −0.75 −0.50 −0.25 0.00 0.25 0.50 0.75 1.00
Spearman's rho (ρ)

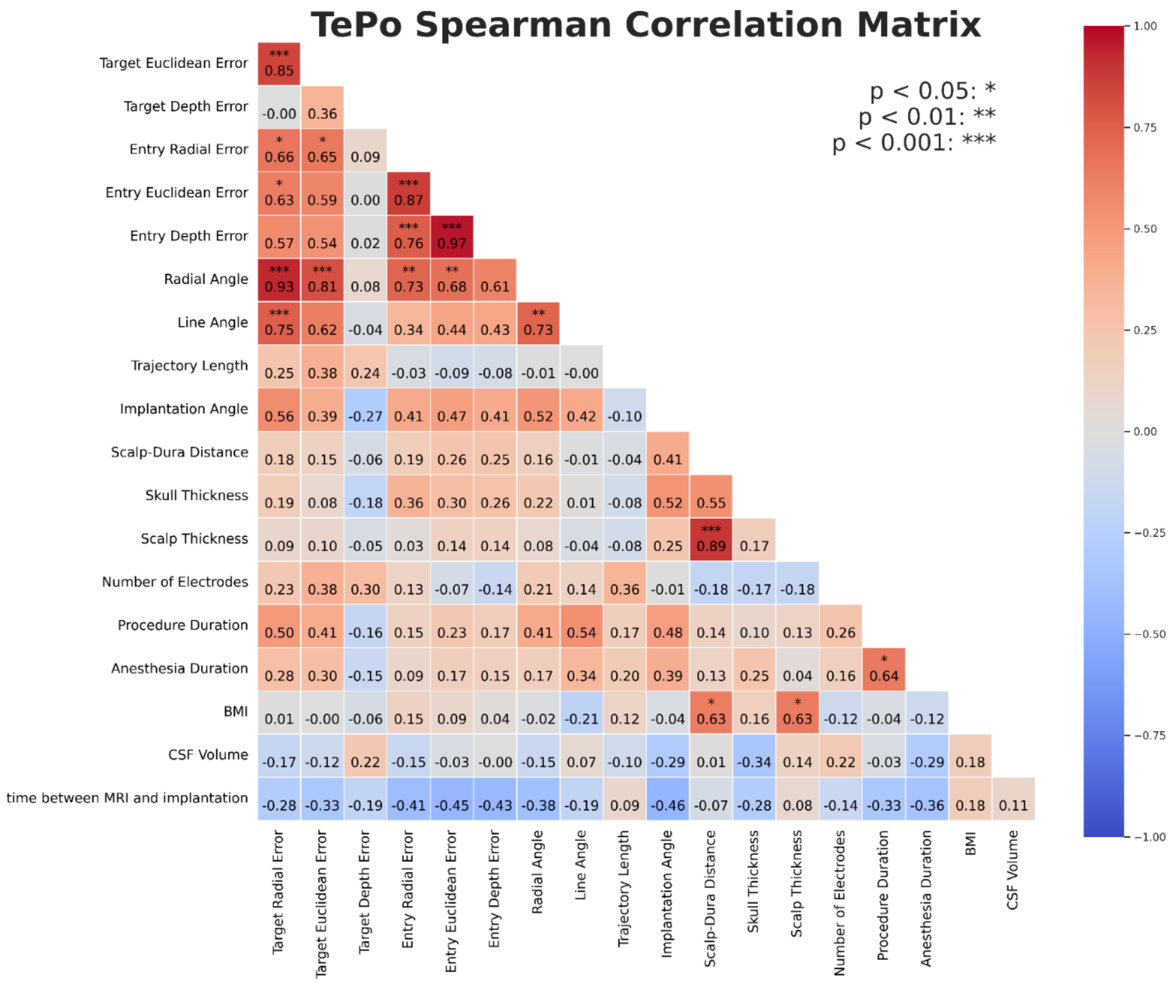
TePo Spearman Correlation Matrix
p < 0.05: *
p < 0.01: **
p < 0.001: ***
Target Euclidean Error *** 0.85
Target Depth Error -0.00 0.36
Entry Radial Error * 0.66 * 0.65 0.09
Entry Euclidean Error * 0.63 0.59 0.00 *** 0.87
Entry Depth Error 0.57 0.54 0.02 *** 0.76 *** 0.97
Radial Angle *** 0.93 *** 0.81 0.08 ** 0.73 ** 0.68 0.61
Line Angle *** 0.75 0.62 -0.04 0.34 0.44 0.43 ** 0.73
Trajectory Length 0.25 0.38 0.24 -0.03 -0.09 -0.08 -0.01 -0.00
Implantation Angle 0.56 0.39 -0.27 0.41 0.47 0.41 0.52 0.42 -0.10
Scalp-Dura Distance 0.18 0.15 -0.06 0.19 0.26 0.25 0.16 -0.01 -0.04 0.41
Skull Thickness 0.19 0.08 -0.18 0.36 0.30 0.26 0.22 0.01 -0.08 0.52 0.55
Scalp Thickness 0.09 0.10 -0.05 0.03 0.14 0.14 0.08 -0.04 -0.08 0.25 *** 0.89 0.17
Number of Electrodes 0.23 0.38 0.30 0.13 -0.07 -0.14 0.21 0.14 0.36 -0.01 -0.18 -0.17 -0.18
Procedure Duration 0.50 0.41 -0.16 0.15 0.23 0.17 0.41 0.54 0.17 0.48 0.14 0.10 0.13 0.26
Anesthesia Duration 0.28 0.30 -0.15 0.09 0.17 0.15 0.17 0.34 0.20 0.39 0.13 0.25 0.04 0.16 * 0.64
BMI 0.01 -0.00 -0.06 0.15 0.09 0.04 -0.02 -0.21 0.12 -0.04 * 0.63 0.16 * 0.63 -0.12 -0.04 -0.12
CSF Volume -0.17 -0.12 0.22 -0.15 -0.03 -0.00 -0.15 0.07 -0.10 -0.29 0.01 -0.34 0.14 0.22 -0.03 -0.29 0.18
time between MRI and implantation -0.28 -0.33 -0.19 -0.41 -0.45 -0.43 -0.38 -0.19 0.09 -0.46 -0.07 -0.28 0.08 -0.14 -0.33 -0.36 0.18 0.11
Target Radial Error
Target Euclidean Error
Target Depth Error
Entry Radial Error
Entry Euclidean Error
Entry Depth Error
Radial Angle
Line Angle
Trajectory Length
Implantation Angle
Scalp-Dura Distance
Skull Thickness
Scalp Thickness
Number of Electrodes
Procedure Duration
Anesthesia Duration
BMI
CSF Volume
1.00
0.75
0.50
0.25
0.00
−0.25
−0.50
−0.75
−1.00

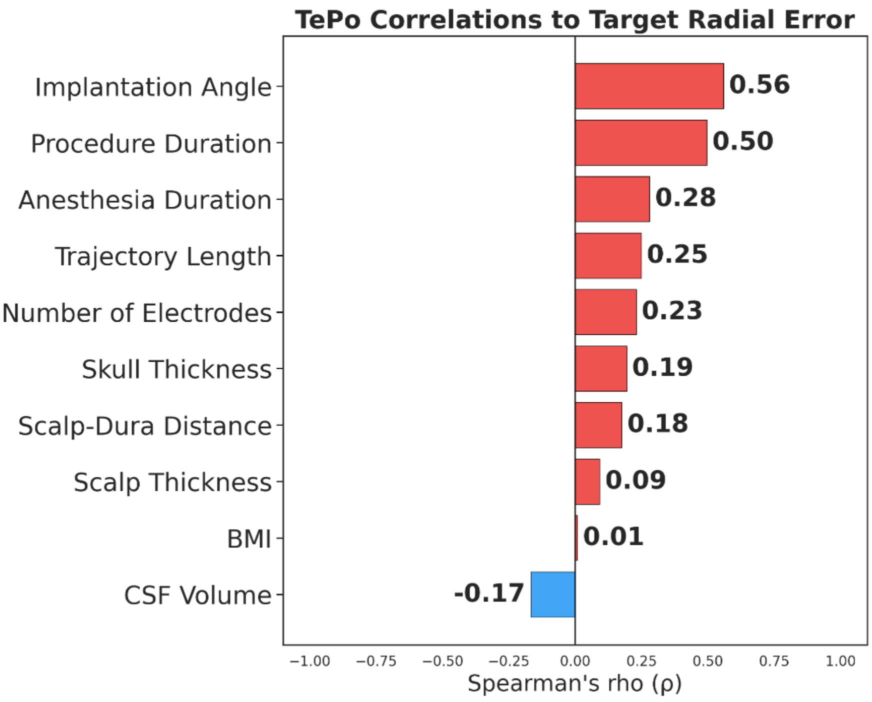
TePo Correlations to Target Radial Error
Implantation Angle 0.56
Procedure Duration 0.50
Anesthesia Duration 0.28
Trajectory Length 0.25
Number of Electrodes 0.23
Skull Thickness 0.19
Scalp-Dura Distance 0.18
Scalp Thickness 0.09
BMI 0.01
CSF Volume -0.17
−1.00 −0.75 −0.50 −0.25 0.00 0.25 0.50 0.75 1.00
Spearman's rho (ρ)

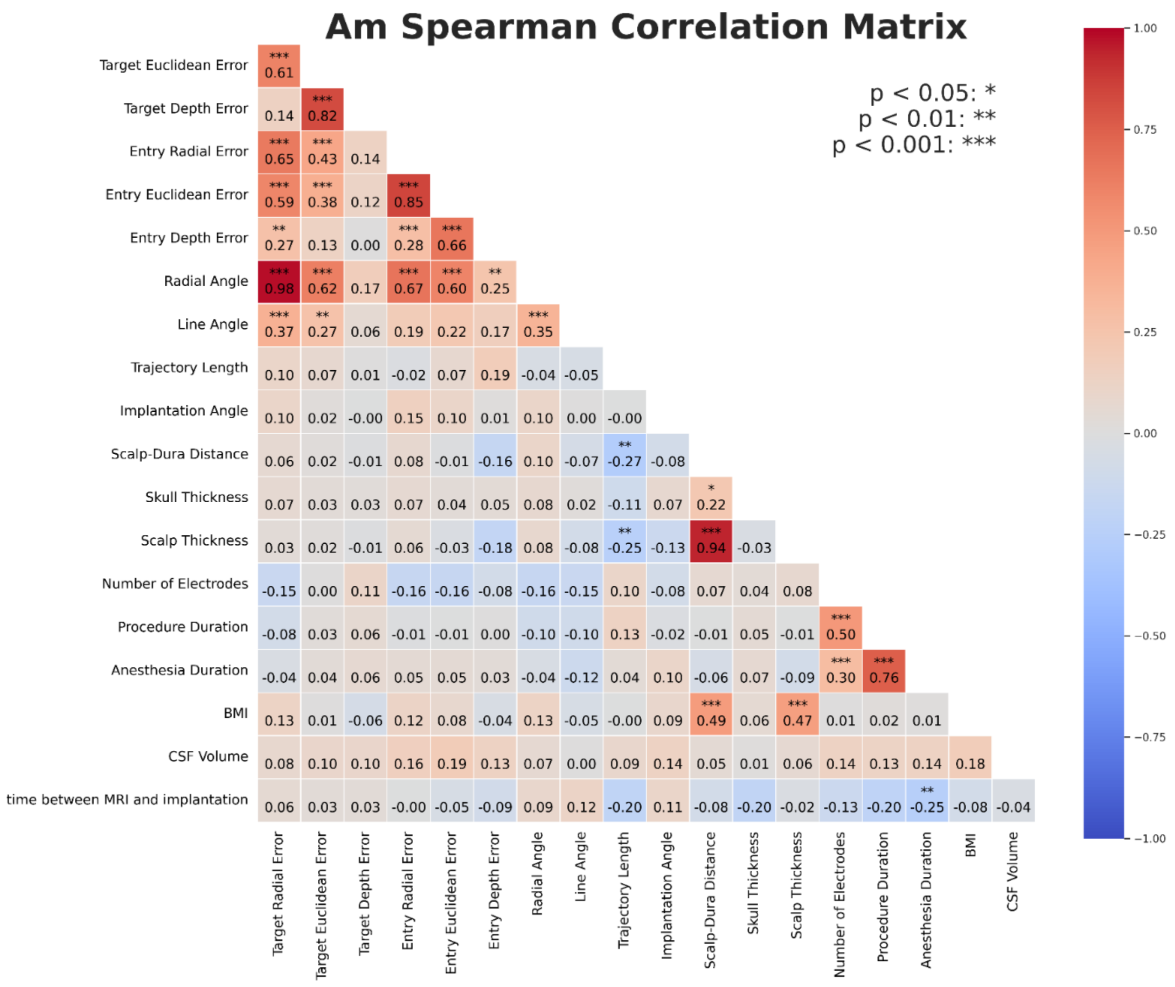
Am Spearman Correlation Matrix
p < 0.05: *
p < 0.01: **
p < 0.001: ***
Target Euclidean Error *** 0.61
Target Depth Error 0.14 *** 0.82
Entry Radial Error *** 0.65 *** 0.43 0.14
Entry Euclidean Error *** 0.59 *** 0.38 0.12 *** 0.85
Entry Depth Error ** 0.27 0.13 0.00 *** 0.28 *** 0.66
Radial Angle *** 0.98 *** 0.62 0.17 *** 0.67 *** 0.60 ** 0.25
Line Angle *** 0.37 ** 0.27 0.06 0.19 0.22 0.17 *** 0.35
Trajectory Length 0.10 0.07 0.01 -0.02 0.07 0.19 -0.04 -0.05
Implantation Angle 0.10 0.02 -0.00 0.15 0.10 0.01 0.10 0.00 -0.00
Scalp-Dura Distance 0.06 0.02 -0.01 0.08 -0.01 -0.16 0.10 -0.07 ** -0.27 -0.08
Skull Thickness 0.07 0.03 0.03 0.07 0.04 0.05 0.08 0.02 -0.11 0.07 * 0.22
Scalp Thickness 0.03 0.02 -0.01 0.06 -0.03 -0.18 0.08 -0.08 ** -0.25 -0.13 *** 0.94 -0.03
Number of Electrodes -0.15 0.00 0.11 -0.16 -0.16 -0.08 -0.16 -0.15 0.10 -0.08 0.07 0.04 0.08
Procedure Duration -0.08 0.03 0.06 -0.01 -0.01 0.00 -0.10 -0.10 0.13 -0.02 -0.01 0.05 -0.01 *** 0.50
Anesthesia Duration -0.04 0.04 0.06 0.05 0.05 0.03 -0.04 -0.12 0.04 0.10 -0.06 0.07 -0.09 *** 0.30 *** 0.76
BMI 0.13 0.01 -0.06 0.12 0.08 -0.04 0.13 -0.05 -0.00 0.09 *** 0.49 0.06 *** 0.47 0.01 0.02 0.01
CSF Volume 0.08 0.10 0.10 0.16 0.19 0.13 0.07 0.00 0.09 0.14 0.05 0.01 0.06 0.14 0.13 0.14 0.18
time between MRI and implantation 0.06 0.03 0.03 -0.00 -0.05 -0.09 0.09 0.12 -0.20 0.11 -0.08 -0.20 -0.02 -0.13 -0.20 ** -0.25 -0.08 -0.04
Target Radial Error
Target Euclidean Error
Target Depth Error
Entry Radial Error
Entry Euclidean Error
Entry Depth Error
Radial Angle
Line Angle
Trajectory Length
Implantation Angle
Scalp-Dura Distance
Skull Thickness
Scalp Thickness
Number of Electrodes
Procedure Duration
Anesthesia Duration
BMI
CSF Volume
1.00
0.75
0.50
0.25
0.00
−0.25
−0.50
−0.75
−1.00

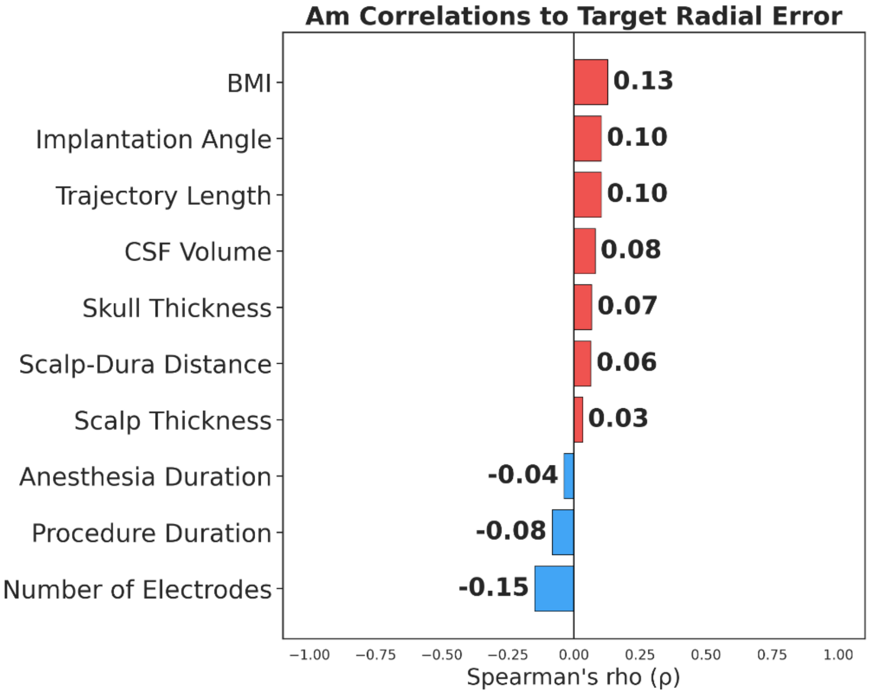
Am Correlations to Target Radial Error
BMI 0.13
Implantation Angle 0.10
Trajectory Length 0.10
CSF Volume 0.08
Skull Thickness 0.07
Scalp-Dura Distance 0.06
Scalp Thickness 0.03
Anesthesia Duration -0.04
Procedure Duration -0.08
Number of Electrodes -0.15
−1.00
−0.75
−0.50
−0.25
0.00
0.25
0.50
0.75
1.00
Spearman's rho (ρ)

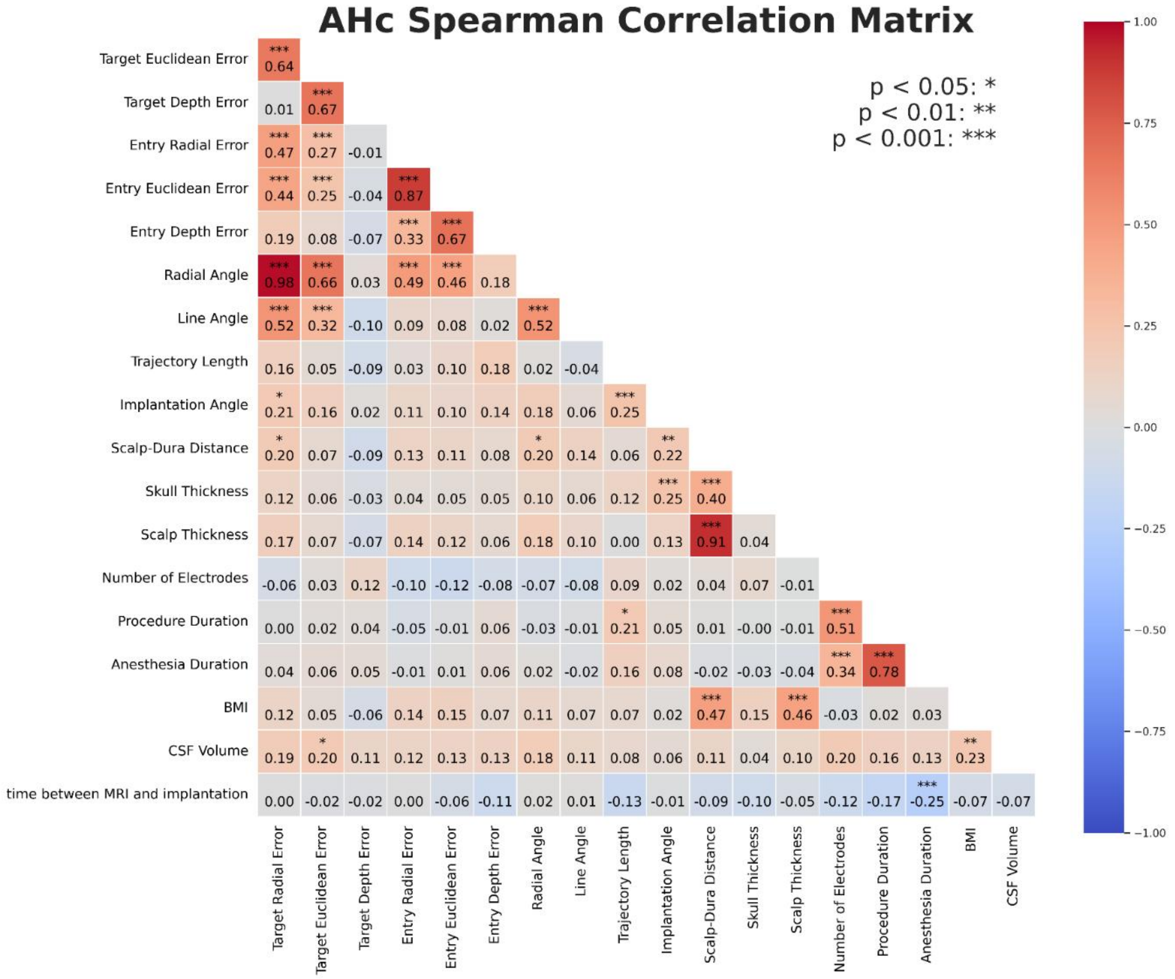
AHc Spearman Correlation Matrix
p < 0.05: *
p < 0.01: **
p < 0.001: ***
Target Euclidean Error
Target Depth Error
Entry Radial Error
Entry Euclidean Error
Entry Depth Error
Radial Angle
Line Angle
Trajectory Length
Implantation Angle
Scalp-Dura Distance
Skull Thickness
Scalp Thickness
Number of Electrodes
Procedure Duration
Anesthesia Duration
BMI
CSF Volume
time between MRI and implantation
Target Radial Error
Target Euclidean Error
Target Depth Error
Entry Radial Error
Entry Euclidean Error
Entry Depth Error
Radial Angle
Line Angle
Trajectory Length
Implantation Angle
Scalp-Dura Distance
Skull Thickness
Scalp Thickness
Number of Electrodes
Procedure Duration
Anesthesia Duration
BMI
CSF Volume
1.00
0.75
0.50
0.25
0.00
−0.25
−0.50
−0.75
−1.00

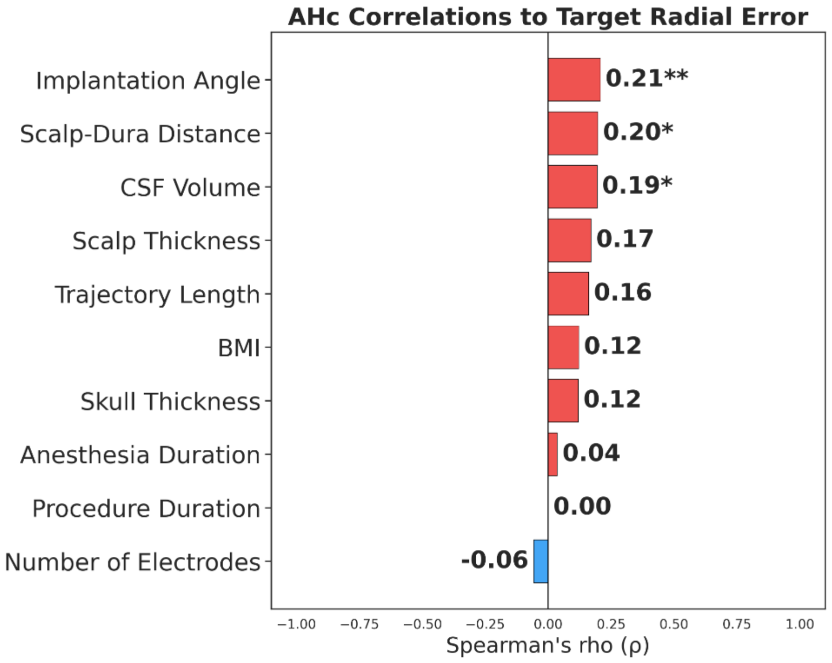
AHc Correlations to Target Radial Error
Implantation Angle
0.21**
Scalp-Dura Distance
0.20*
CSF Volume
0.19*
Scalp Thickness
0.17
Trajectory Length
0.16
BMI
0.12
Skull Thickness
0.12
Anesthesia Duration
0.04
Procedure Duration
0.00
Number of Electrodes
-0.06
−1.00 −0.75 −0.50 −0.25 0.00 0.25 0.50 0.75 1.00
Spearman's rho (ρ)

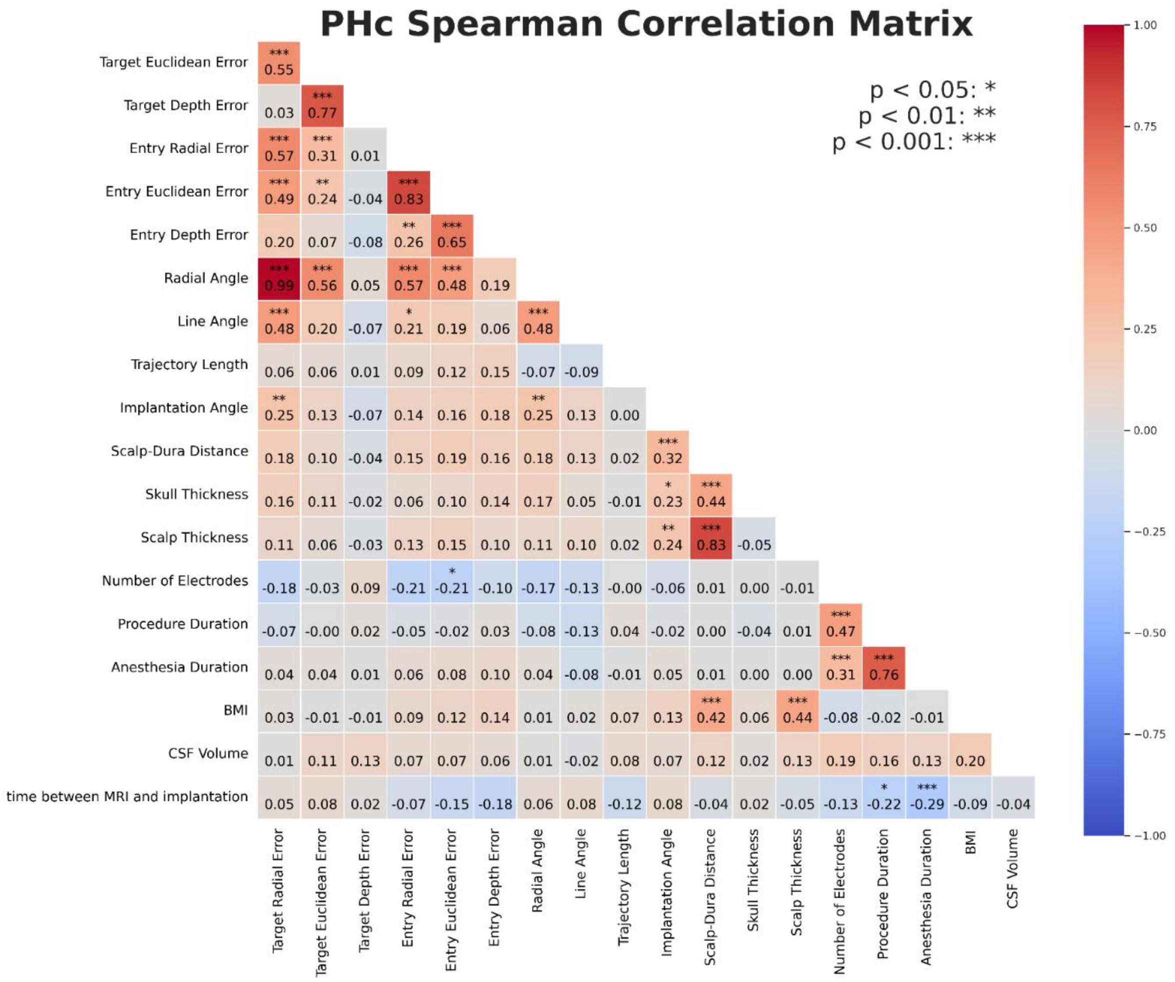
PHc Spearman Correlation Matrix
p < 0.05: *
p < 0.01: **
p < 0.001: ***
Target Euclidean Error
Target Depth Error
Entry Radial Error
Entry Euclidean Error
Entry Depth Error
Radial Angle
Line Angle
Trajectory Length
Implantation Angle
Scalp-Dura Distance
Skull Thickness
Scalp Thickness
Number of Electrodes
Procedure Duration
Anesthesia Duration
BMI
CSF Volume
time between MRI and implantation
Target Radial Error
Target Euclidean Error
Target Depth Error
Entry Radial Error
Entry Euclidean Error
Entry Depth Error
Radial Angle
Line Angle
Trajectory Length
Implantation Angle
Scalp-Dura Distance
Skull Thickness
Scalp Thickness
Number of Electrodes
Procedure Duration
Anesthesia Duration
BMI
CSF Volume
1.00
0.75
0.50
0.25
0.00
−0.25
−0.50
−0.75
−1.00

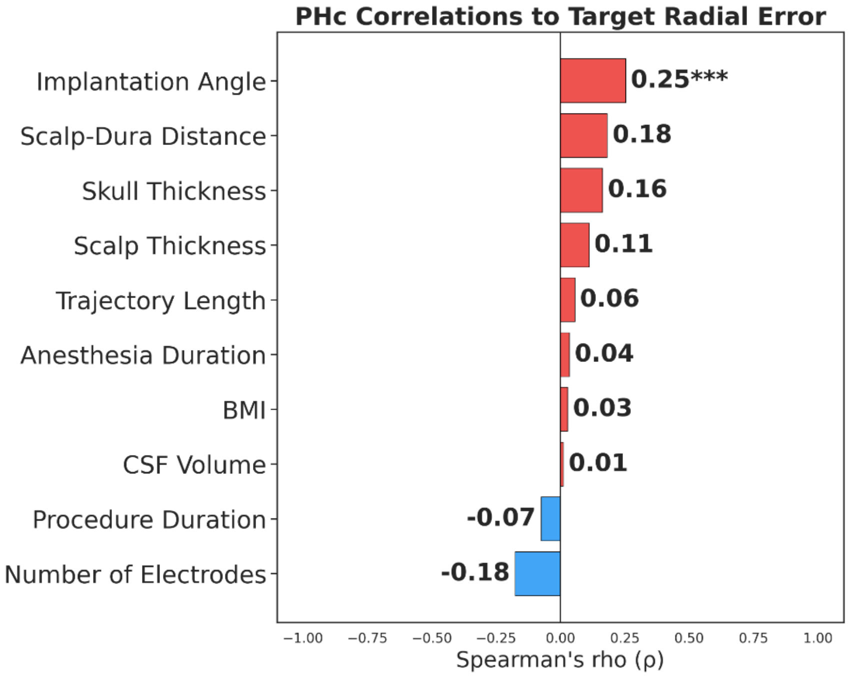
PHc Correlations to Target Radial Error
Implantation Angle
Scalp-Dura Distance
Skull Thickness
Scalp Thickness
Trajectory Length
Anesthesia Duration
BMI
CSF Volume
Procedure Duration
Number of Electrodes
0.25***
0.18
0.16
0.11
0.06
0.04
0.03
0.01
-0.07
-0.18
−1.00
−0.75
−0.50
−0.25
0.00
0.25
0.50
0.75
1.00
Spearman's rho (ρ)

### Appendix

**Naming Convention**

| Direction | |
|---|---|
| A | Anterior |
| C/Ce | Center/Central |
| I | Inferior |
| L | Lateral |
| M | Medial |
| P | Posterior/Post |
| S | Superior |
| Inf | Infra |
| Int | Intra |
| Ln | Line |
| Ma | Margin |
| Me | Mesial |
| Mi | Middle/Mid |
| Or | Orthogonal |
| Par | Para |
| ParC | Paracentral |
| Pr | Pre/Pr |
| Ps | Parasagittal |
| Sa | Sagittal |
| Sp | Supplementary |
| Sup | Supra |
| Sc | Sensory cortex |
| Mc | Motor cortex |
| Te | Temporal |
| Pa | Parietal |
| Oc | Occipital |
| Fr | Frontal |
| Mo | Motor |

| Structure | |
|---|---|
| Cu | Cuneus |
| Am | Amygdala |
| An | Angular |
| Br | Broca |
| Ca | Calcarine |
| Cc | Corpus Callosum |
| Cg | Cingulate |
| Cl | Cleft |
| Cv | Cavernoma |
| Cx | Convex |
| Dy | Dysplasia |
| Ec | Entorhinal cortex |
| Ece | Encephalocele |
| Fa | Face |
| Gy | Gyrus |
| Ha | Hand |
| Hc | Hippocampus |
| Hs | Heschel's |
| Ht | Heterotopia |
| In | Insula |
| Lg | Lingula |
| Lo | lobe/lobule |
| Ls | Lesion |
| O | Orbital |
| Op | Operculum |
| Po | Pole |
| Re | Resection |
| Ro | Rolandic |
| Rs | Rostrum |
| Sl | Splenium |
| SMA | Supplementary motor area |
| Tu | Tuber |
| Unc | Uncus |
| We | Wernicke |

**Metrics and Variables for Surgeon-Labeled Trajectories**

| *Anatomical Target* | *Metric/Variable* | *median (IQR)* | mean ± std |
|---|---|---|---|
| **Anterior Hippocampus (n=337)** | **Euclidean error (mm)** | | |
| | Entry | 1.21 (0.79-1.71) | 1.36 ± 0.85 |
| | Target | 2.36 (1.68-3.12) | 2.48 ± 1.18 |
| | **Radial error (mm)** | | |
| | Entry | 1.01 (0.60-1.42) | 1.13 ± 0.78 |
| | Target | 1.45 (0.89-2.08) | 1.60 ± 0.99 |
| | **Depth error (mm)** | | |
| | Entry | 0.45 (0.19-0.91) | 0.63 ± 0.57 |
| | Target | 1.55 (0.76-2.18) | 1.64 ± 1.16 |
| | **Radial Angle** | 1.83 (1.17-2.67) | 2.05 ± 1.26 |
| | **Line Angle** | 1.09 (0.69-1.61) | 1.34 ± 0.99 |
| | **Implantation Angle** | 18.06 (13.10-22.55) | 18.22 ± 6.97 |
| | **Trajectory Length** | 45.30 (42.90-48.20) | 45.49 ± 4.44 |
| | **Scalp-Dura Distance** | 12.70 (11.10-14.70) | 13.11 ± 2.97 |
| | **Skull Thickness** | 1.76 (1.25-2.75) | 2.10 ± 1.15 |
| | **Scalp Thickness** | 10.66 (9.28-12.20) | 11.01 ± 2.66 |
| **Posterior Hippocampus (n=291)** | **Euclidean error (mm)** | | |
| | Entry | 1.19 (0.72-1.71) | 1.37 ± 0.94 |
| | Target | 2.07 (1.42-2.88) | 2.30 ± 1.61 |
| | **Radial error (mm)** | | |
| | Entry | 0.87 (0.56-1.39) | 1.07 ± 0.82 |
| | Target | 1.18 (0.73-1.69) | 1.32 ± 0.90 |
| | **Depth error (mm)** | | |

| | | | |
|---|---|---|---|
| | Entry | 0.46 (0.15-0.91) | 0.66 ± 0.70 |
| | Target | 1.35 (0.69-2.31) | 1.65 ± 1.62 |
| | **Radial Angle** | 1.45 (0.90-2.16) | 1.68 ± 1.21 |
| | **Line Angle** | 1.01 (0.61-1.60) | 1.22 ± 0.94 |
| | **Implantation Angle** | 10.40 (7.67-14.74) | 11.94 ± 6.92 |
| | **Trajectory Length** | 46.50 (43.90-49.00) | 46.34 ± 5.15 |
| | **Scalp-Dura Distance** | 12.60 (10.80-14.25) | 12.77 ± 2.63 |
| | **Skull Thickness** | 3.01 (2.45-3.96) | 3.24 ± 1.36 |
| | **Scalp Thickness** | 9.21 (7.88-11.00) | 9.53 ± 2.37 |
| **Anterior Insula (n=289)** | **Euclidean error (mm)** | | |
| | Entry | 1.64 (1.08-2.34) | 1.88 ± 1.33 |
| | Target | 2.18 (1.57-2.98) | 2.61 ± 2.96 |
| | **Radial error (mm)** | | |
| | Entry | 1.19 (0.70-1.84) | 1.36 ± 0.93 |
| | Target | 1.57 (1.00-2.26) | 1.80 ± 1.17 |
| | **Depth error (mm)** | | |
| | Entry | 0.79 (0.30-1.48) | 1.08 ± 1.19 |
| | Target | 1.15 (0.51-1.81) | 1.52 ± 2.94 |
| | **Radial Angle** | 1.24 (0.78-1.88) | 1.43 ± 0.94 |
| | **Line Angle** | 0.86 (0.49-1.20) | 0.99 ± 0.76 |
| | **Implantation Angle** | 27.05 (24.06-29.71) | 27.11 ± 4.61 |
| | **Trajectory Length** | 73.10 (69.30-77.40) | 72.68 ± 8.60 |
| | **Scalp-Dura Distance** | 17.00 (15.10-18.80) | 17.25 ± 2.94 |

| | | | |
|---|---|---|---|
| | **Skull Thickness** | 8.31 (6.91-9.65) | 8.38 ± 2.07 |
| | **Scalp Thickness** | 8.62 (7.51-9.86) | 8.87 ± 2.21 |
| **Orbitofrontal (n=271)** | **Euclidean error (mm)** | | |
| | Entry | 1.45 (0.93-2.11) | 1.69 ± 1.14 |
| | Target | 2.46 (1.75-3.26) | 2.63 ± 1.19 |
| | **Radial error (mm)** | | |
| | Entry | 1.20 (0.72-1.72) | 1.27 ± 0.81 |
| | Target | 1.65 (1.10-2.31) | 1.85 ± 1.14 |
| | **Depth error (mm)** | | |
| | Entry | 0.52 (0.23-1.15) | 0.89 ± 1.03 |
| | Target | 1.51 (0.86-2.17) | 1.60 ± 1.03 |
| | **Radial Angle** | 1.88 (1.24-2.73) | 2.14 ± 1.36 |
| | **Line Angle** | 1.11 (0.72-1.52) | 1.28 ± 0.87 |
| | **Implantation Angle** | 28.58 (23.35-33.80) | 28.79 ± 8.02 |
| | **Trajectory Length** | 50.70 (47.50-53.50) | 50.82 ± 5.02 |
| | **Scalp-Dura Distance** | 14.00 (12.30-16.70) | 14.59 ± 3.43 |
| | **Skull Thickness** | 3.72 (2.44-4.90) | 3.97 ± 1.91 |
| | **Scalp Thickness** | 10.25 (8.35-12.52) | 10.62 ± 3.22 |
| **Amygdala (n=259)** | **Euclidean error (mm)** | | |
| | Entry | 1.53 ± 0.91 | 1.33 (0.93-1.94) |
| | Target | 2.52 ± 1.59 | 2.35 (1.61-3.17) |
| | **Radial error (mm)** | | |
| | Entry | 1.21 ± 0.78 | 1.10 (0.69-1.56) |
| | Target | 1.48 ± 0.83 | 1.39 (0.90-1.92) |

| | | | |
|---|---|---|---|
| | **Depth error (mm)** | | |
| | Entry | 0.76 ± 0.72 | 0.60 (0.24-1.06) |
| | Target | 1.82 ± 1.64 | 1.65 (0.94-2.39) |
| | **Radial Angle** | 1.91 ± 1.10 | 1.71 (1.15-2.50) |
| | **Line Angle** | 1.09 ± 0.70 | 0.95 (0.65-1.34) |
| | **Implantation Angle** | 20.09 ± 6.74 | 19.83 (15.91-24.27) |
| | **Trajectory Length** | 45.88 ± 4.27 | 45.90 (43.20-48.70) |
| | **Scalp-Dura Distance** | 15.45 ± 3.80 | 15.10 (12.90-17.02) |
| | **Skull Thickness** | 1.83 ± 1.08 | 1.56 (1.21-2.11) |
| | **Scalp Thickness** | 13.63 ± 3.69 | 12.97 (11.00-15.33) |
| **Posterior Insula (n=238)** | **Euclidean error (mm)** | | |
| | Entry | 1.89 ± 1.40 | 1.64 (1.08-2.40) |
| | Target | 2.72 ± 2.25 | 2.35 (1.58-3.13) |
| | **Radial error (mm)** | | |
| | Entry | 1.25 ± 1.13 | 1.07 (0.60-1.54) |
| | Target | 2.04 ± 1.66 | 1.74 (1.14-2.61) |
| | **Depth error (mm)** | | |
| | Entry | 1.24 ± 1.07 | 1.01 (0.43-1.74) |
| | Target | 1.35 ± 1.92 | 0.91 (0.46-1.78) |
| | **Radial Angle** | 1.29 ± 1.06 | 1.04 (0.72-1.64) |
| | **Line Angle** | 1.05 ± 0.93 | 0.83 (0.55-1.26) |
| | **Implantation Angle** | 22.06 ± 5.69 | 21.98 (18.23-25.67) |
| | **Trajectory Length** | 92.37 ± 12.96 | 94.50 (90.20-98.90) |

| | | | |
|---|---|---|---|
| | **Scalp-Dura Distance** | 18.08 ± 3.39 | 17.75 (15.62-20.00) |
| | **Skull Thickness** | 7.74 ± 2.21 | 7.60 (6.09-9.11) |
| | **Scalp Thickness** | 10.34 ± 2.41 | 9.95 (8.69-11.66) |
| **Posterior Cingulate (n=104)** | **Euclidean error (mm)** | | |
| | Entry | 1.17 ± 0.64 | 1.13 (0.69-1.51) |
| | Target | 2.16 ± 0.82 | 2.17 (1.64-2.66) |
| | **Radial error (mm)** | | |
| | Entry | 0.88 ± 0.48 | 0.79 (0.51-1.19) |
| | Target | 1.20 ± 0.64 | 1.06 (0.66-1.67) |
| | **Depth error (mm)** | | |
| | Entry | 0.60 ± 0.65 | 0.40 (0.14-0.78) |
| | Target | 1.65 ± 0.88 | 1.68 (0.88-2.13) |
| | **Radial Angle** | 1.21 ± 0.67 | 1.10 (0.68-1.71) |
| | **Line Angle** | 0.87 ± 0.50 | 0.75 (0.53-1.22) |
| | **Implantation Angle** | 13.13 ± 5.14 | 13.07 (9.27-16.88) |
| | **Trajectory Length** | 58.45 ± 7.49 | 57.55 (54.55-60.85) |
| | **Scalp-Dura Distance** | 14.10 ± 3.42 | 13.35 (11.86-15.47) |
| | **Skull Thickness** | 4.74 ± 1.75 | 4.53 (3.55-5.86) |
| | **Scalp Thickness** | 9.36 ± 2.49 | 8.96 (7.73-10.47) |
| **Anterior Cingulate (n=98)** | **Euclidean error (mm)** | | |
| | Entry | 1.61 ± 0.91 | 1.35 (1.01-2.04) |
| | Target | 2.42 ± 0.91 | 2.21 (1.76-3.15) |
| | **Radial error (mm)** | | |
| | Entry | 1.26 ± 0.56 | 1.18 (0.86-1.61) |

| | | | |
|---|---|---|---|
| | Target | 1.54 ± 0.83 | 1.35 (0.98-2.08) |
| | **Depth error (mm)** | | |
| | Entry | 0.82 ± 0.91 | 0.56 (0.24-1.01) |
| | Target | 1.69 ± 0.89 | 1.57 (1.13-2.17) |
| | **Radial Angle** | 1.85 ± 1.02 | 1.57 (1.12-2.44) |
| | **Line Angle** | 0.89 ± 0.64 | 0.75 (0.46-1.08) |
| | **Implantation Angle** | 20.33 ± 7.92 | 20.00 (14.93-25.63) |
| | **Trajectory Length** | 50.15 ± 7.89 | 49.15 (45.08-53.60) |
| | **Scalp-Dura Distance** | 14.94 ± 3.56 | 14.40 (12.60-16.77) |
| | **Skull Thickness** | 5.40 ± 2.73 | 4.64 (3.42-6.82) |
| | **Scalp Thickness** | 9.54 ± 3.21 | 8.81 (7.22-11.39) |
